# Drug combinations proposed by machine learning on genes/proteins to improve the efficacy of Tecovirimat in the treatment of Monkeypox: A Systematic Review and Network Meta-analysis

**DOI:** 10.1101/2023.04.23.23289008

**Authors:** Mahnaz Boush, Ali A. Kiaei, Danial Safaei, Sadegh Abadijou, Nader Salari, Masoud Mohammadi

## Abstract

**Background:** The World Health Organization (WHO) describes Monkeypox as a viral zoonosis, or an animal-to-human virus transmission, with symptoms comparable to those of past smallpox patients but clinically less severe. This study’s objective is to assess the results of previous investigations on the best drug combinations for treating Monkeypox.

**Method:** The pharmacological combinations used to treat monkeypox sickness have been researched in two stages for this systematic review and network meta-analysis. To begin with, a certain machine learning technique is used to extract the medication combinations from the researched articles offered on science databases, including Scopus, PubMed, Web of Science (ISI), Science Direct, Embase, and Google Scholar. Second, the tested medicine combinations will have been proven.

**Results:** The results of this study show that the p-value between the proposed drug combination and Monkeypox for scenarios 1 to 5 were 0.108, 0.042, 0.023, 0.018, and 0.015, respectively. Scenario *i* is the combination of the first *i* suggested drugs for treating Monkeypox. This has led to a 720 percent increase in the proposed drug combination’s efficacy in treating Monkeypox.

**Conclusion:** The suggested drug combination decreases the p-value between MonkeyPox and the genes as potential targets for Monkeypox progression, which leads to an improvement in the treatment of Monkeypox. Therefore, using the right combination of drugs is important in improving the community’s health and reducing per capita treatment costs.

## INTRODUCTION

West and Central Africa are home to the uncommon and potentially lethal zoonotic virus known as Monkeypox. It is caused by the orthopoxvirus Monkeypox, which is related to the Variola virus that causes smallpox and the live virus used in orthopoxvirus vaccines and can infect humans. A 2017–2018 outbreak with 118 confirmed cases was discovered in Nigeria after 39 years without the discovery of human disease; sporadic cases still occur. (Rao et al., 2022)

### Rationale

Various drugs have been proposed to block these human genes’ receptors to treat this disease. Among these drugs, the following can be mentioned: Adefovir, bafilomycin A1, dideoxyglucose hydrate, N-carbonyl, N’-carbonyl, diamino, smallpox vaccine, Tecovirimat, Cidofovir, QS21, Clodronic Acid. Among these drugs, some of them have been mentioned in various articles as effective drugs in the treatment of MonkeyPox, such as:

#### Tecovirimat

The Biomedical Advances Research and Development Authority of the U.S. Department of Health and Human Services and SIGA Technologies worked together to develop the orthopoxvirus-specific antiviral medication tecovirimat (TPOXX®). It functions by impeding the activity of the orthopoxvirus VP37 envelope-wrapping protein, which is essential for producing egress-competent enveloped virions, which are required for the virus to spread within the host. Oral tecovirimat was given FDA approval in the USA in July 2018 for treating adult and pediatric patients with human smallpox disease brought on by the variola virus who weigh less than 13 kg. Tecovirimat was approved according to the US FDA’s Animal Rule, which bases marketing approval on a product’s effectiveness in pertinent animal models. (Hoy, 2018)

#### Smallpox vaccine

After being eliminated as a natural disease in the latter decades of the 20th century through a concerted international effort, smallpox has returned as a potential bioterrorist weapon. Smallpox vaccines are now receiving more attention as a result. Despite having a successful clinical history as an efficient smallpox vaccine, live vaccinia virus, an orthopoxvirus closely related to smallpox, is associated with uncommon but serious adverse events. As a result, research into newer-generation smallpox vaccines has recently increased and has improved safety profiles and sustained efficacy. (Artenstein and Grabenstein, 2008)

#### Bafilomycin A1

To measure the activity of the autophagic flux, bafilomycin A1 is frequently used in vitro as an inhibitor of autophagosome-lysosome fusion. Targeting the V-ATPase ATP6V0C/V0 subunit c, a macrolide known as bafilomycin A1 inhibits lysosomal acidification by preventing protons from entering the lysosomal lumen. It is generally agreed that bafilomycin A1 prevents V-ATPase pump activity from preventing autophagosome-lysosome fusion when all of the drug’s properties are considered. As in mammalian cell culture in vitro, we found that bafilomycin A1 inhibits lysosome acidification and autophagosome-lysosome fusion in Drosophila fat body cells. (Mauvezin and Neufeld, 2015)

#### Cidofovir

Cidofovir is a nucleotide analog that prevents human cytomegalovirus (CMV) infection by inhibiting viral DNA polymerase. Cellular enzymes phosphorylate it to make it active. Cidofovir can be given every week during induction therapy and every other week during maintenance therapy due to the prolonged intracellular half-lives of its metabolites. Although it has developed in vitro, viral resistance has not yet been observed in patients receiving cidofovir treatment. Compared to patients with AIDS who delayed treatment, immediate cidofovir therapy prevented the progression of CMV retinitis.

Additionally, after receiving prior treatment, CMV retinitis relapsing was prevented by cidofovir. Probenecid and intravenous saline hydration must be given with each dose of cidofovir to prevent nephrotoxicity. (Lea and Bryson, 1996)

#### Clodronic Acid

A halogenated non-nitrogen-containing bisphosphonate known as clodronic acid is effective against many diseases linked to excessive bone resorption. Contrary to the acute-phase and inflammatory effects of bisphosphonates containing nitrogen, the medication also appears to have analgesic and anti-inflammatory properties. According to theory, it causes osteoclasts to undergo apoptosis, preventing bone resorption. Clodronic acid has been proven successful in maintaining or boosting bone mineral density when given orally, intramuscularly, or intravenously to patients with osteoporosis. Also known to be reduced with drug use is the risk of fractures. (Frediani et al., 2009)

Each of these drugs affects only several human genes associated with MonkeyPox. Therefore, it is important to suggest the correct combination of drugs that can affect more genes than the above. To explore the effective drug combination in the treatment of MonkeyPox, in this study, a native machine learning model has been used.

### Objectives

To provide a holistic yet thorough genes/proteins network meta-analysis on the impact of the drug combination on the treatment of Monkeypox, we decided to perform a systematic review of the prior research in this area. This was motivated by the numerous reported drug treatment consequences for Monkeypox (Figure 1) and the absence of global statistics on the subject. This paper makes an effort to review, systematically examine, and analyze the literature as well as the results that have been reported to better understand the effects of the suggested drug combination on the treatment of Monkeypox.

**Figure 1:**
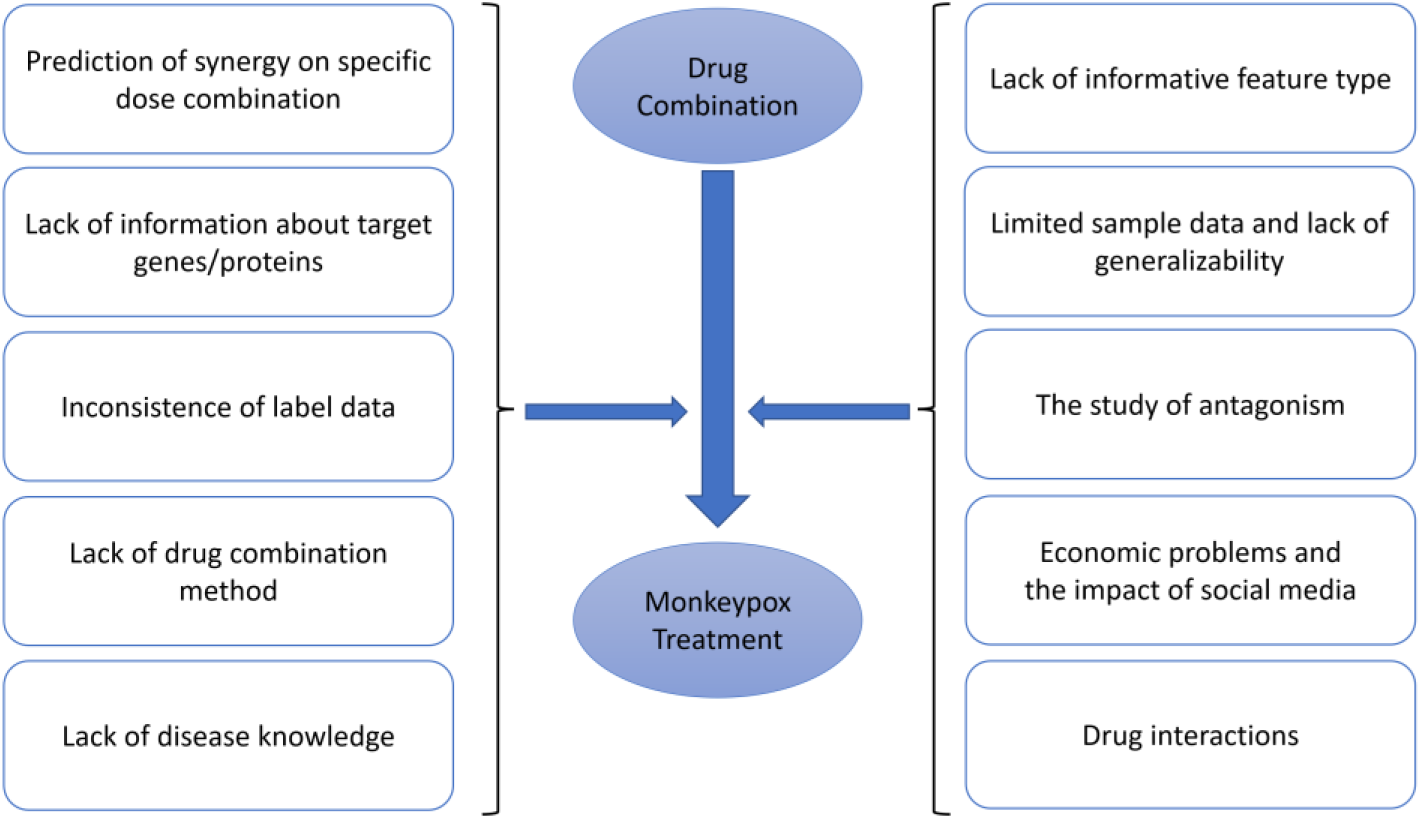
Impacts of suggested drug combinations on monkeypox treatment

## METHOD

### Information sources

In this step, a systematic review is carried out using NLP techniques to locate relevant research in Scopus, PubMed, Web of Science (ISI), Science Direct, Embase, and Google Scholar. Although our native machine learning model provides the medicine combination, as we have already indicated, the essay’s uniqueness is in verifying the suggested drug combination, not in using that model. So, we employed a systematic review to validate our work. The keywords are extracted from the Monkeypox subscription and the outputs of the ML model in the first step. These terms are Monkeypox, Tecovirimat, Smallpox vaccine, bafilomycin A1, Cidofovir, and Clodronic Acid.

### Search strategy

An NLP technique has been hired to search semantically among the Titles / Abstracts of articles by connecting to these databases. Therefore, semantic search is so valuable that it can also consider MeSH terms for keywords to search. For example, the word Monkeypox in semantic search is equivalent to MPV, MPXV, hMPXV, etc. There is no specific time limit for searching related articles.

### Eligibility criteria and Selection process

Selected studies for systematic review include the following: 1) Studies that include one or more of the proposed ML MODEL and Monkeypox drugs. 2) Study-oriented research 3) Full-text research. The systematic review entering criteria are indicated as 1-the term Monkeypox and studies involving one or more of the proposed drugs that ML MODEL explored. 2-Observational research 3-Studies whose full texts are available on science databases. Additionally, studies with the following criteria are eliminated from the systematic review: 1-Unrelated papers, 2-Studies with insufficient data, 3-Multiple sources, 4-Interventional studies, and 5-Studies with ambiguous methods.

### Study selection

In the beginning, redundant studies are removed. A list of the titles of all remaining studies is created to filter the research in a structured manner during the evaluation phase. In the systematic review’s initial phase, screening, the titles, and abstracts of the remaining studies are carefully examined, and the selection criteria exclude several studies. Secondly, the competency assessment, the full text of the remaining studies from the screening phase, is carefully examined according to the criteria, and several unrelated studies are thus eliminated. To avoid the influence of taste on resource selection, research and data extraction are performed by an expert and an NLP Question-Answering (Q.A.) agent completely independent. The expert must write the reason for not selecting the research fully and accurately. On the other hand, the Q.A. agent gives each article a score for the questions asked. Articles with low ratings will be deleted. Questions asked by the Q.A. agent are such as Is *this drug* effective for the treatment of MonkeyPox? The word “*this drug”* is replaced by a drug for each output of the intelligent system, generating an independent question. If there is a disagreement between the expert and the output of the Q.A. agent, the latter expert will review the disputed research. In the present study, 17 cases were selected for the third stage after performing the stated steps.

### Quality evaluation

The final studies, which will have elected to enter into the systematic review, have to assess based on STROBE checklists. The checklists are widely utilized to assess the quality of observational studies. A STROBE checklist contains six scales: title, abstract, introduction, methods, results, and discussion, and also some of the scales have their subscales, resulting in a total of 32 subscales. The majority of the subscales include the following: title, problem statement, study objectives, study type, statistical population, sampling method, sample size, the definition of variables and procedures, data collection technique(s), statistical analysis methods, and findings. As a result, During the quality assessment phase, a score of 32 is the maximum that can be obtained when utilizing the STROBE checklist. A score of 16 has been established as the cutoff point in this study. As a result, articles with a score of 16 or higher are deemed medium or high quality, and papers below 16 were disqualified from the study. Nineteen high or medium-quality papers were included in the systematic review phase after being evaluated for quality using the STROBE checklist. This phase validates the information gathered in the first step. The executive protocol for this paper is based on RAIN (Mohammadi et al., 2021). Numerous research studies have employed the RAIN protocol as a framework for investigating various psychological and emotional phenomena (Kiaei et al., 2022, 2023; Salari et al., 2023, 2022a, 2022b, 2021).

### Study risk of bias assessment

At this stage, the effectiveness of each drug on human genes is calculated based on the p-value. The results will be presented in circular bar charts, radar charts, and Table **3**.

## RESULTS

The effect of the medications mentioned above on the management of Monkeypox was assessed in this study. By PRISMA’s guidelines, the papers with the specified focus were systematically reviewed after being gathered with no time restriction. The reference management software, Zotero, received 350 potential related articles that were found during the initial search. Duplicate studies accounted for 207 of the studies, which were removed. Following the screening stage, 42 articles were discarded. Following a review of the full texts and similar consideration of the inclusion and exclusion criteria, 68 studies were eliminated during the eligibility evaluation phase. At the quality assessment stage, four studies that were deemed to be works of low methodological quality were excluded, and 29 studies made it to the final analysis stage after receiving a grade for each paper based on the STROBE checklist and evaluation of the full text of the articles (please see Figure 2). Additionally, these articles’ specifics and traits are provided in Table 1.

**Figure 2:**
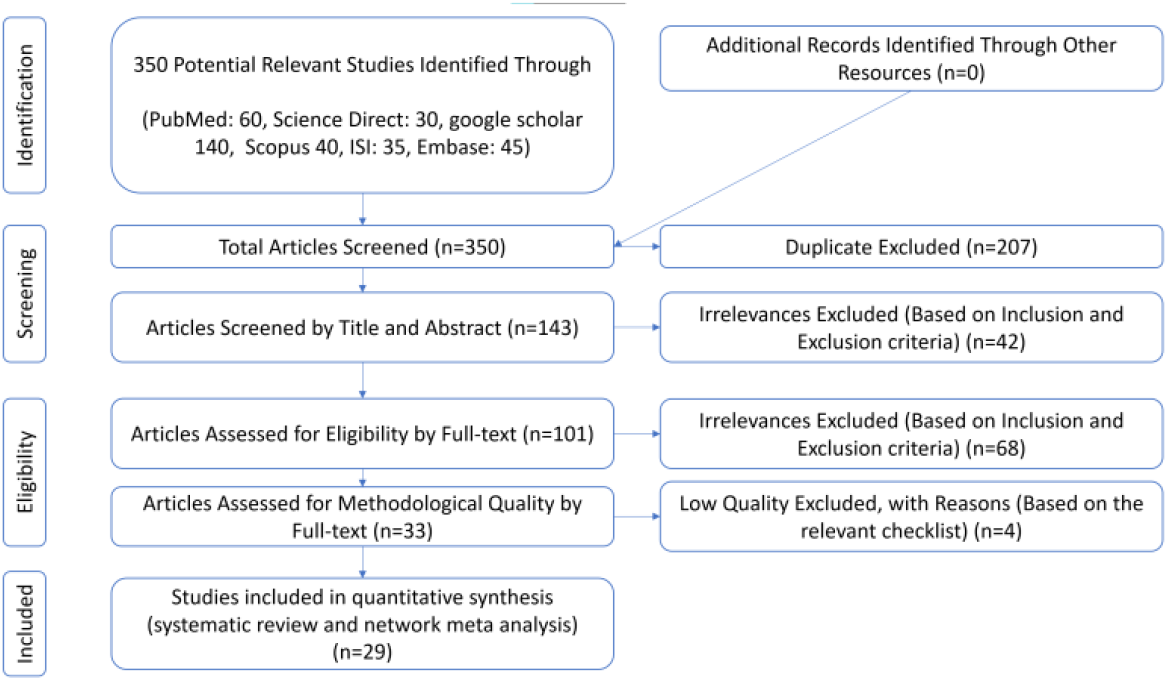
PRISMA flow diagram illustrating the stages of the article examined in this systematic review and network meta-analysis.

**Table 1:**
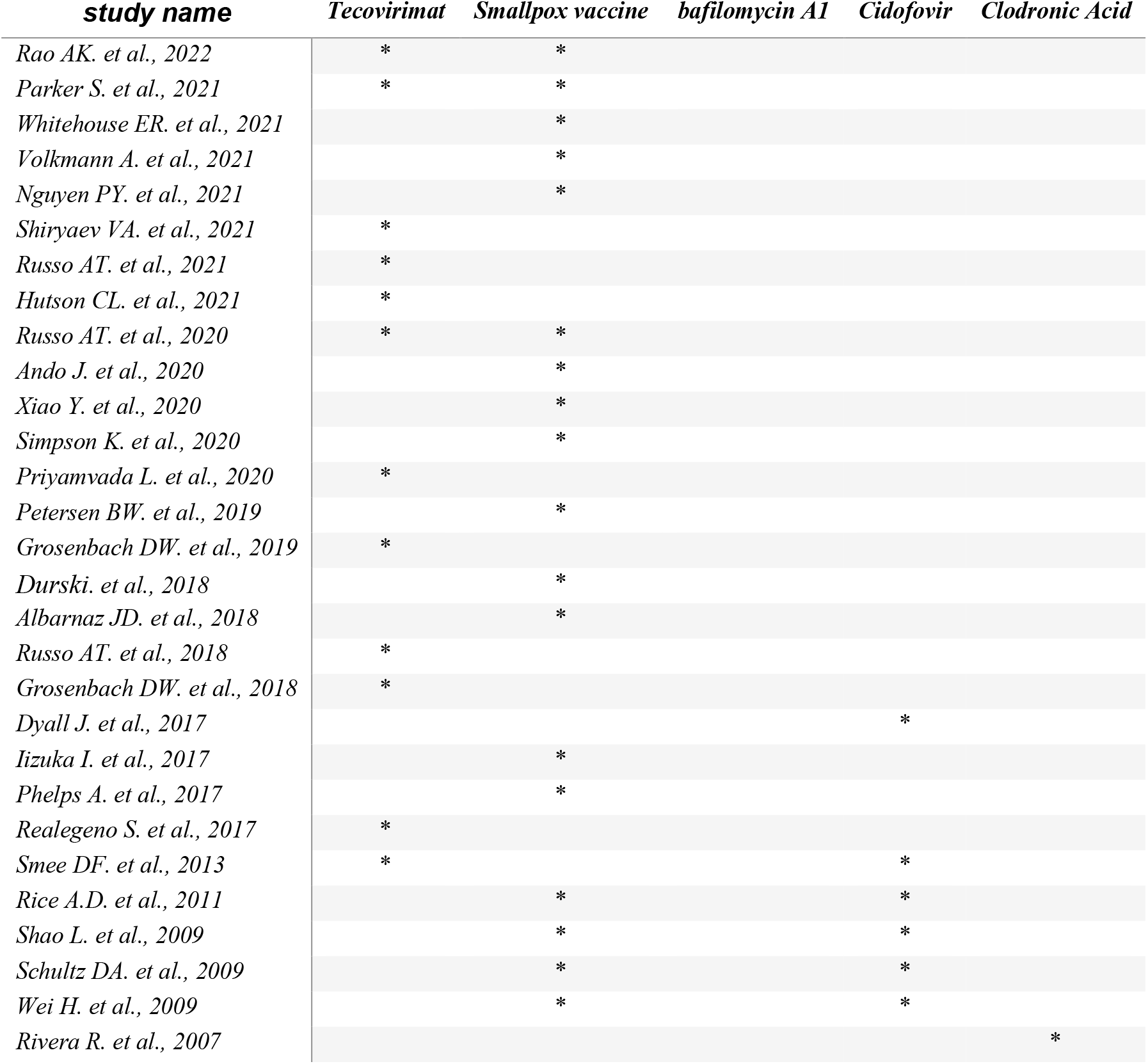
some important research studies for proposed drugs in Monkeypox treatments

### Genes Network Meta-Analysis

Table 2 indicates the p-value of combining above mentioned drugs. For instance, the p-value between Monkeypox and Tecovirimat and Smallpox vaccine combination (Scenario 2) indicated 0.042 and decreased to 0.023 when *bafilomycin A1* was added to the combination (Scenario 3). Moreover, as shown in Table 2, the P-value after applying the fifth scenario has shown that the proposed drug combination had a good effect in treating the disease.

**Table 2:** p-value between scenarios and Monkeypox

| *Scenario* | *Drug Combinations* | *p-value* |
| --- | --- | --- |
| S1 | Tecovirimat | 0.107661605 |
| S2 | S1 + Smallpox vaccine | 0.041676985 |
| S3 | S2 + Bafilomycin A1 | 0.022916327 |
| S4 | S3 + Cidofovir | 0.018140499 |
| S5 | S4 + Clodronic Acid | 0.015230822 |

As shown in Table 3, the p-values between human genes and Monkeypox have been changed by new scenarios. The ‘S0’ column shows the p-value between Monkeypox and the corresponding affected human genes. The “S1” column shows the combined p-value between Monkeypox and the human genes used by the Tecovirimat. The p-values between Monkeypox and many human genes reach 1 in the ‘S5’ column, where scenarios are defined in Table 2.

**Table 3:** p-values between Monkeypox and human genes before/after different scenarios

| Association Name | S0 | S1 | S2 | S3 | S4 | S5 | Association Name | S0 | S1 | S2 | S3 | S4 | S5 |
| --- | --- | --- | --- | --- | --- | --- | --- | --- | --- | --- | --- | --- | --- |
| TWF2 | 6E-04 | 0 | 0 | 0 | 0 | 0 | OCM | 0.005 | 0.01 | 0.97 | 0.97 | 0.97 | 0.97 |
| UBXN2B | 7E-04 | 0.99 | 0.99 | 0.99 | 1 | 1 | CYB5R3 | 0.006 | 0.01 | 0.99 | 1 | 1 | 1 |
| NUP37 | 8E-04 | 0.99 | 0.99 | 0.99 | 0.99 | 1 | MX1 | 0.006 | 0.01 | 0.01 | 0.91 | 0.91 | 0.91 |
| KLRC3 | 0.001 | 0 | 0 | 0 | 0 | 0 | BIK | 0.007 | 0.01 | 0.01 | 0.01 | 0.01 | 0.01 |
| VPS53 | 0.001 | 0.99 | 0.99 | 0.99 | 1 | 1 | DPAGT1 | 0.009 | 0.01 | 0.01 | 0.01 | 0.01 | 0.01 |
| CCNH | 0.001 | 0.99 | 0.99 | 0.99 | 0.99 | 1 | CTNNBIP1 | 0.01 | 0.01 | 0.01 | 0.01 | 0.01 | 0.01 |
| SAMD9L | 0.002 | 0 | 0 | 0 | 0 | 0 | GPT | 0.01 | 0.01 | 0.01 | 0.01 | 0.01 | 0.01 |
| TMED10 | 0.002 | 0 | 0 | 0 | 0 | 0 | EXT1 | 0.013 | 0.01 | 0.01 | 0.01 | 0.01 | 0.96 |
| C8A | 0.002 | 0 | 0 | 0 | 0 | 0 | ERVW-1 | 0.013 | 0.99 | 0.99 | 0.99 | 1 | 1 |
| KLHL2 | 0.002 | 0 | 0 | 0.99 | 1 | 1 | TKT | 0.015 | 0.02 | 0.02 | 0.02 | 0.99 | 0.99 |
| BP4 | 0.002 | 0 | 0 | 0 | 0 | 0 | HLA-A | 0.017 | 0.02 | 0.02 | 0.02 | 0.02 | 0.02 |
| C6 | 0.002 | 0 | 0 | 0 | 0 | 0 | HAP1 | 0.019 | 0.02 | 0.02 | 0.94 | 0.94 | 0.94 |
| GPA33 | 0.002 | 0 | 0.99 | 0.99 | 0.99 | 0.99 | KLRK1 | 0.019 | 0.02 | 0.02 | 0.65 | 0.65 | 0.65 |
| LRRC32 | 0.002 | 0.99 | 0.99 | 0.99 | 1 | 1 | UNG | 0.019 | 0.02 | 0.02 | 0.02 | 0.02 | 0.02 |
| TM9SF2 | 0.003 | 0 | 0 | 0 | 0 | 0 | CD4 | 0.022 | 0.97 | 0.99 | 1 | 1 | 1 |
| SAMD9 | 0.003 | 0 | 0 | 0 | 0 | 0 | MAPKAPK2 | 0.024 | 0.02 | 0.02 | 0.02 | 0.02 | 0.02 |
| VPS51 | 0.003 | 0.99 | 0.99 | 0.99 | 1 | 1 | EIF2AK2 | 0.025 | 0.02 | 0.85 | 0.99 | 1 | 1 |
| CNGB1 | 0.003 | 0.99 | 0.99 | 1 | 1 | 1 | IGKV1-16 | 0.025 | 0.03 | 0.99 | 0.99 | 1 | 1 |
| VPS54 | 0.003 | 0.99 | 1 | 1 | 1 | 1 | FAM49B | 0.026 | 0.03 | 0.99 | 0.99 | 0.99 | 0.99 |
| VPS52 | 0.004 | 0.99 | 0.99 | 0.99 | 1 | 1 | PSMA7 | 0.026 | 0.03 | 0.03 | 0.03 | 0.03 | 0.03 |

While Figure 3(a) indicates the p-values between affected human genes and Monkeypox, Figure 3(b) shows the p-values between them after implementing the fifth scenario. Moreover, Figure 4 shows a radar chart to indicate the efficiency of the drug selected using the drug selection algorithm by demonstrating the p-values between Monkeypox and human genes after consuming selected drugs. Each colored line shows the effectiveness of the corresponding drug in that scenario.

**Figure 3:**
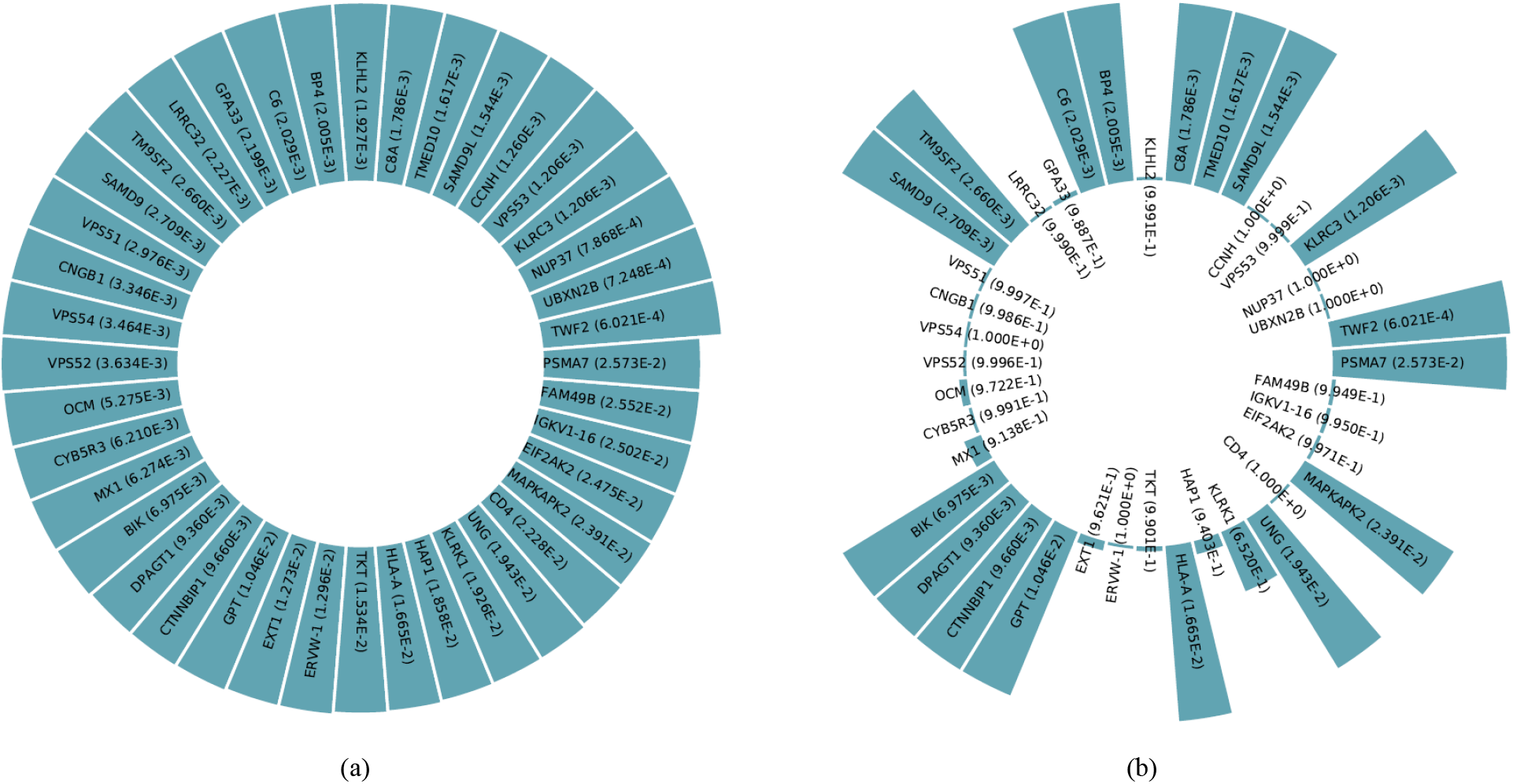
p-values between affected human genes and Monkeypox {(a)before/(b)after} implementing Scenario 5.

**Figure 4:**
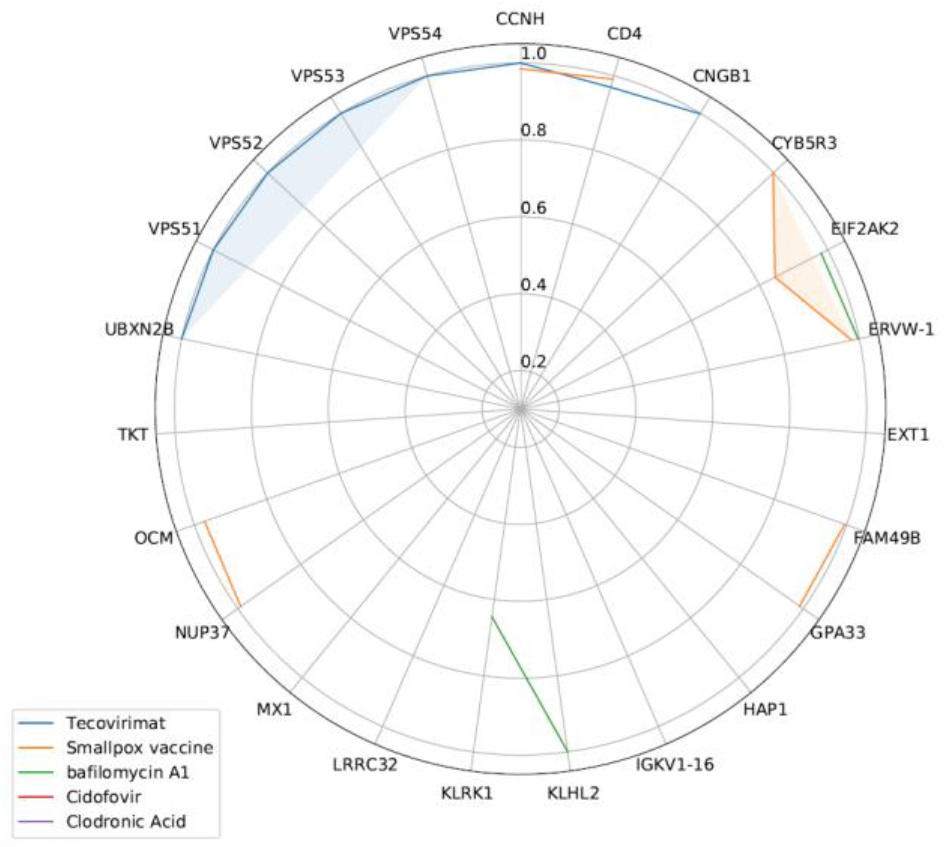
radar chart for p-values between Monkeypox and affected genes after consumption of each drug.

## DISCUSSION

Monkeypox has taken over as the most significant orthopoxvirus for public health since smallpox was eradicated in 1980, and smallpox vaccinations were subsequently discontinued. Primarily affecting central and west Africa, Monkeypox has been spreading into cities and is frequently found close to tropical rainforests. Numerous rodent species and non-human primates serve as hosts for animals.

Due to the unknown nature of this disease and its severe impact on human genes, the use of safe and effective drug combinations for treatment is very important. Proposed drug combinations should be administered with the greatest positive effect on the genes involved. Therefore, it is important to suggest an effective drug combination that can significantly affect the genes involved. For this purpose, in this study, a native machine learning model is used to provide effective drug combinations to treat MonkeyPox disease.

This research included four main steps: extraction of drug combinations using our native machine learning, systematic review, human genes network meta-analysis, and prescription drug information. In the first step, a native machine-learning model proposes drug combinations to treat MonkeyPox disease. In the second step, the proposed drugs of this combination are reviewed during a systematic review to validate the gathered data in the first step. A network meta-analysis analyzes the effectiveness of this proposed drug combination on human genes in the third step. In the fourth step, the prescription drug information of these drug combinations is investigated (such as drug interactions, side effects, drugs, and foods completions, etc.)

The study results show that native machine learning prescribes drugs that cover a wider range of human genes involved in MonkeyPox. Different studies use machine learning in medicine (Kiaei et al., 2019; Kiaei and Khotanlou, 2017). The validity of the proposed drug combinations has been investigated by a systematic review, gene network meta-analysis, and prescription drug information.

### Related genes/proteins

The relevance of the genes identified as potential targets for Monkeypox progression has been validated in many studies, some of which are shown in Table 4. These studies have indicated that MonkeyPox disease has led to the involvement of various genes/proteins. Among the cases with the lowest p-value, the following can be mentioned: TWF2, UBXN2B, NUP37, KLRC3, VPS53, CCNH, SAMD9L, TMED10, C8A, KLHL2, BP4, C6, GPA33, LRRC32, TM9SF2, SAMD9, VPS51, CNGB1, VPS54, VPS52, OCM, CYB5R3, MX1, BIK, DPAGT1, CTNNBIP1, GPT, EXT1, ERVW-1, TKT, HLA-A, HAP1, KLRK1, UNG, CD4, MAPKAPK2, EIF2AK2, GKV1-16, AM49B, SMA7.

**Table 4.** lists studies identifying genes as potential targets for Monkeypox progression.

| Study name | Gene | Study name | Gene | Study name | Gene | Study name | Gene |
| --- | --- | --- | --- | --- | --- | --- | --- |
| *Alkhalil et al., 2009* | TWF2 | *axby, 1982* | BIK | *Shchelkunov et al., 2002* | KLHL2 | *Mitra-Kaushik et al., 2007, p. 4* | CD4 |
| *Shiryayev et al., 2021* | UBXN2B | *Douglas and Dumbell, 1996* | DPAGT1 | *Baxby, 1982* | BP4 | *Goulding et al., 2012* | CD4 |
| *Duraffour et al., 2010* | UBXN2B | *Rajeh et al., 2021* | CTNNBIP1 | *Ibrahim et al., 1998* | C6 | *Silva Gomes et al., 2012* | CD4 |
| *Yang et al., 2005* | UBXN2B | *Douglas and Dumbell, 1996* | GPT | *Golden and Hooper, 2008* | GPA33 | *Sivapalasingam et al., 2007* | CD4 |
| *Shiryayev et al., 2021* | NUP37 | *Luteijn et al., 2019* | EXT1 | *Realegeno et al., 2017* | LRRC32 | *Kennedy et al., 2011* | CD4 |
| *Duraffour et al., 2010* | NUP37 | *Shiryayev et al., 2021* | ERVW-1 | *Levine et al., 2009* | LRRC32 | *Moutafsi et al., 2010* | CD4 |
| *Yang et al., 2005* | NUP37 | *Xu et al., 2011* | ERVW-1 | *Kennedy et al., 2011* | CYB5R3 | *Grosenbach et al., 2010* | CD4 |
| *Fang et al., 2011* | KLRC3 | *Duraffour et al., 2010* | ERVW-1 | *Luteijn et al., 2019* | TM9SF2 | *Hammarlund et al., 2008* | CD4 |
| *Fang et al., 2011* | VPS53 | *Li et al., 2006* | ERVW-1 | *Zhang et al., 2019* | SAMD9 | *Karem et al., 2007* | CD4 |
| *Realegeno et al., 2017* | VPS53 | *Yang et al., 2005* | ERVW-1 | *Realegeno et al., 2017* | VPS51 | *Probst et al., 2013* | CD4 |
| *Shiryayev et al., 2021* | CCNH | *Upton and McFadden, 1986* | TKT | *Realegeno et al., 2017* | CNGB1 | *Gordon et al., 2011* | CD4 |
| *Duraffour et al., 2010* | CCNH | *Esposito and Knight, 1984* | TKT | *Zhang et al., 2019* | CNGB1 | *Edghill-Smith et al., 2005* | CD4 |
| *Yang et al., 2005* | CCNH | *Jing et al., 2005* | HLA-A | *Realegeno et al., 2017* | VPS54 | *Alkhalil et al., 2009* | MAPKAPK2 |
| *Ando et al., 2020, p. 77* | SAMD9L | *Luteijn et al., 2019* | HAP1 | *Realegeno et al., 2017* | VPS52 | *Arndt et al., 2015* | EIF2AK2 |
| *Luteijn et al., 2019* | TMED10 | *Lazeur et al., 2013* | KLRK1 | *Gordon et al., 2011* | OCM | (Fogg et al., 2007) | IGKV1-16 |
| *(Mucker et al., 2018)* | C8A | (Campbell et al., 2007) | KLRK1 | (Lorenzo et al., 2017) | MX1 | (Fogg et al., 2007) | FAM49B |
| *(Totmenin et al., 2002)* | KLHL2 | (Duraffour et al., 2007) | UNG | (Johnston et al., 2012) | MX1 | (Ibrahim et al., 1998) | PSMA7 |

Since our article focuses on the validity of the proposed drug combination (not the machine learning model), we have used a systematic review to verify the results. Many studies use the systematic review to validate a claim of a relationship between two subjects. (Salari et al., 2021) A comprehensive systematic review and meta-analysis show global osteoporosis prevalence among the world’s older adults. (Salari et al., 2022a) Works on systematic review to find the global prevalence of Duchenne and Becker muscular dystrophy.

### Details of the Method

In the first step of this research, a native Machine Learning (ML) model proposed different drug combinations for MonkeyPox. This machine learning model receives p-value information between disease and related biological data, which are human genes, and on the other hand, p-values between the same human genes and effective drugs, as input. And in the output suggests a drug combination after which the p-value between the disease and the same human genes drives close to 1.

Although our native machine learning model suggests the drug combination, the article’s novelty is not in the presentation of that model but in the presentation of the drug combination. Therefore, we have used the systematic review to validate our work. In other words, to validate the results of our native ML model, in the second step, the proposed drugs of the first step are examined in a systematic review. Previous articles reviewed have been extracted from Scopus, Science Direct, Web of Science (ISI), PubMed, Embase, and Google Scholar. In this step, these databases use semantic search methods based on Natural Language Processing (NLP) instead of manual search. The advantage of this is the MeSH search of each keyword. For example, searching for the word Artificial Intelligence in semantic search is equivalent to the following keywords: machine learning, Deep Learning, Transfer Learning, transformer methods, SVM, Reinforcement learning, and other related keywords which may even be unknown to us. It is considered that a wider and more accurate range of articles is explored in a short time.

In the third step, a network meta-analysis examines the proposed synthetic drug combinations on human genes. In this analysis, the effectiveness of each drug on the input biological data that are human genes affected by MonkeyPox is shown. Then, Table 3 examines the p-value between human genes and MonkeyPox before and after the drug combination.

After the proposed drug combinations have been validated in steps two and three, prescription drug information is investigated in step four.

#### Proposed drug combination using our ML model

Many studies have used machine Learning for medicine. For example, (Jafari et al., 2022) developed a comprehensive pipeline for diagnosing and predicting the risk of chronic diseases for a cohort study using deep learning and Shapley values.

In this step, the native ML model receives a weight between Target (Monkeypox) and Associations (Drugs) as input through an Interface Feature, which is biological data (human gene). This weight is calculated by p-value. Figure 5 shows the structure of our native ML model. The p-value between the Target and human genes is first considered in this figure. For example, 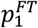 represents the p-value between the human gene F1 (as interface feature) and the Monkeypox (as Target). On the other hand, this model receives the p-value between these genes and different drugs. (For example, 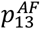 represents the p-value between the first Association and the third Interface human gene.) Using the above biological data as inputs, the ML model estimates the combined p-value between Associations and the Target. (For example, 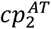 represents the combined p-value between the second Association and Target.)

**Figure 5:**
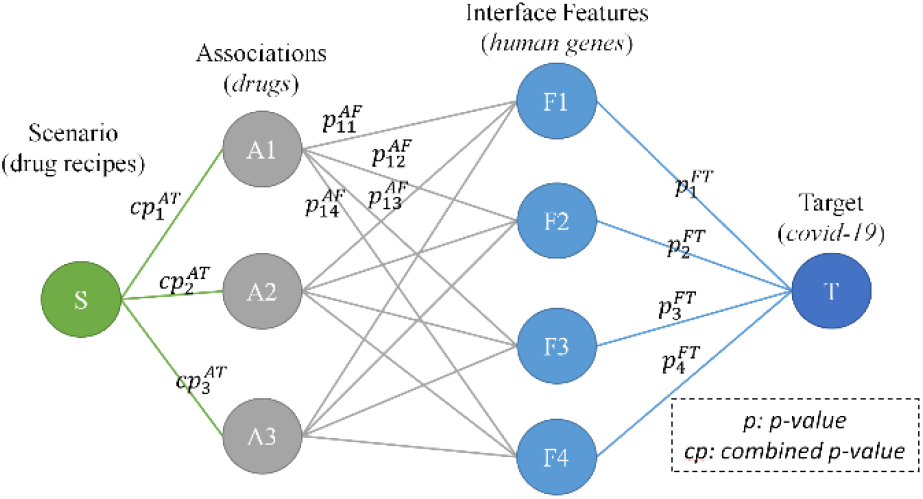
The general structure of the native ML model to suggest an effective drug combination in the treatment of disease using human genes as interface features

By selecting the Association with the lowest combined p-value, the ML model updates all weights based on how much it affects the Interface features. This update is done so that associations have a similar effect on human genes with the selected Association; their p-value will be larger. This process is then repeated until the ML model achieves its stop condition.

The ML model has selected drugs whose p-values with Monkeypox are small. The drug selection algorithm (as stated before, we can’t reveal the algorithm because it has some license issues) determines the p-values of each drug concerning Monkeypox. The drug with the lowest combined p-value is then chosen. After selecting the first drug, the drug selection algorithm updates weights between Monkeypox and affected human genes according to the p-values between those genes and the selected drug. So, all the combined p-values between these drugs and Monkeypox are updated. The drug selection algorithm iterates finding the drug with the lowest combined p-value and updating weights. In the end, the drug selection algorithm proposed scenarios, where each scenario contains a drug recipe, as is the aim of the drug selection algorithm for making associations cooperation.

#### Systematic Review

Although our native machine learning model offers the drug combination, the innovation of the essay is in the presentation of the drug combination rather than the model. So, to validate our study, we used a systematic review. We have employed a systemic review to validate our work in this important step. In this subsection, we have focused on some articles in Table 1 that confirm subgroups of the proposed drug combination.

According to Rao AK. et al., the patients who had Monkeypox underwent a 32-day hospital stay that included tecovirimat treatment due to the severity of the illness. To provide cross-protection against Monkeypox, it was also suggested that the patient get vaccinated against smallpox. (Rao et al., 2022)

The variola virus, the cause of smallpox, has been eradicated, according to Parker S. et al., thanks to a successful vaccination program. The mousepox-causing ectromelia virus has also been used in research, demonstrating that IL-4 recombinant viruses are significantly more virulent than wild-type viruses, even in mice given vaccines and/or antiviral medications. It is crucial to develop antiviral treatments for these viruses for the reasons mentioned above in case they accidentally or purposefully spread. The U.S. stockpile currently contains two antivirals (tecovirimat and brincidofovir) that can treat smallpox in an emergency.(Parker et al., 2021)

According to Whitehouse ER. et al., the incidence of Monkeypox was lower in patients who were assumed to have received smallpox vaccination than in patients who were assumed to have not received the vaccination. (Whitehouse et al., 2021) Volkmann A. et al. state that MVA-BN, which received smallpox vaccine approval in 2013 in Europe and Canada, will also receive monkeypox vaccine approval in the U.S. in 2019.(Volkmann et al., 2021)

The significance of smallpox vaccination was stressed by Nguyen PY et al. A statistical model was created to demonstrate how the growing number of unvaccinated people makes households, rather than just young children, more vulnerable to Monkeypox, raising the possibility of human-to-human transmission. This demonstrates the value of smallpox immunization.(Nguyen et al., 2021) The p37 protein, a tecovirimat target, is one of the potential targets of poxviruses like Monkeypox, according to Shiryaev VA. et al.(Shiryaev et al., 2021)

A significant development in the readiness for biosecurity is the acceptance of tecovirimat as a smallpox treatment. A further improvement in health security would come from the inclusion of tecovirimat in the CDC smallpox response plan, the creation of pediatric liquid and intravenous formulations, and the endorsement of post-exposure prophylaxis. When used against emerging orthopoxvirus diseases like Monkeypox, adverse reactions to vaccinations, and side effects of vaccinia oncolytic virus therapy, tecovirimat exhibits broad anti-orthopoxvirus efficacy both in vitro and in vivo. (Russo et al., 2021)

According to Russo AT. et al., the approval of tecovirimat for the treatment of smallpox represents a significant improvement in biosecurity readiness. The addition of tecovirimat to the CDC’s smallpox response plan, the development of pediatric liquid and intravenous formulations, and getting approval for post-exposure prophylaxis would positively affect health security. Tecovirimat exhibits broad anti-orthopoxvirus efficacy both in vitro and in vivo when used to treat emerging orthopoxvirus diseases like Monkeypox, unfavorable vaccination reactions, and side effects of vaccinia oncolytic virus therapy. (Hutson et al., 2021)

In studies by Russo AT. et al., monkeys simultaneously received tecovirimat and the live attenuated smallpox vaccine ACAM2000TM. The vaccine’s effectiveness in defending against a risky monkeypox virus (MPXV) challenge was assessed along with immune reactions. Despite the use of tecovirimat, primary and anamnestic humoral immune responses were comparable in the two studies. The third study revealed a decline in humoral immunity brought on by the vaccine. Of the 13 animals that had received the vaccine or been given tecovirimat, 12 completed the lethal MPXV challenge. In animals treated with tecovirimat, the illness’ symptoms were more pronounced. This suggests that the immunogenicity of ACAM2000 may change if TPOXX and it are both administered simultaneously. The results of these studies may aid in understanding how TPOXX and the vaccine are used during a smallpox outbreak. (Russo et al., 2020)

Peripheral blood mononuclear cells from 13 healthy volunteers who had previously received the smallpox vaccine (Dryvax) were stimulated and expanded using overlapping E3L peptides in a method by Ando J. et al. to investigate the function, specificity, and antiviral activity of the cells. As a result, E3L-specific T cells killed both peptide-loaded target cells and vaccinia-infected cells. (Ando et al., 2020).

Xiao Y. et al. investigated vaccine formulations with additional significant protein targets, such as A33 and B5 (extracellular virus components) and another protein on the mature virus (A27), which were both adjuvanted with aluminum hydroxide (A.H.) with and without CpG-oligonucleotide. They discovered that CpG, which induced IgG2a-antibody responses, was present in the formulations of the A33 vaccine that were the most effective. (Xiao et al., 2020) According to Simpson K. et al., the experts agreed that the best disease control methods were required, including the potential use of recently developed antivirals and vaccination with new generation non-replicating smallpox vaccines. (Simpson et al., 2020)

According to Priyamvada L. et al., antiviral drugs that are effective as postexposure treatments for variola virus (the cause of smallpox) or other orthopoxviruses are essential in the event of an exposure to or outbreak of orthopoxviruses.

The only treatment for smallpox approved by the U.S. Food and Drug Administration, Tecovirimat (ST-246), functions by limiting the production of extracellular viruses (E.V.), which prevents the virus from spreading from cell to cell and over long distances.(Priyamvada et al., 2020)

The goal of a study described by Petersen BW. et al. was to evaluate the effectiveness, immunogenicity, and safety of a third-generation smallpox vaccine, IMVAMUNE®, in medical professionals who were at risk of contracting the monkeypox virus (MPXV).(Petersen et al., 2019)

Due to the threat posed by variola viruses like Monkeypox, Grosenbach DW. et al. emphasized the necessity of antivirals like tecovirimat.(Grosenbach and Hruby, 2019)

Albarnaz JD. et al. and Durski. et al. emphasize the importance of the smallpox vaccine in eradicating smallpox.(Albarnaz et al., 2018; Durski et al., 2018)

According to Russo AT. et al., tecovirimat therapy is effective smallpox antiviral therapeutic candidate because it increases survival when administered up to 8 days after a lethal aerosol Monkeypox virus challenge and protects against the disease’s clinical manifestations when administered earlier than five days after the challenge. (Russo et al., 2018)

According to Grosenbach DW. Tecovirimat is being marketed as a smallpox treatment following the FDA Animal Rule based on its efficacy in two animal models and human pharmacokinetic and safety data.(Grosenbach et al., 2018) Dyall J. et al. emphasize the importance of cidofovir in treating Monkeypox. (Dyall et al., 2017)

The vaccination of people with LC16m8, a highly attenuated smallpox vaccine, may result in long-term protection against MPXV infection, according to Iizuka I. et al.(Iizuka et al., 2016)

Smallpox vaccination produces an immune response and guards against serious illness and death, according to Phelps A. et al.(Phelps et al., 2017)

Realegeno S. et al. emphasize using antiviral therapy for treating Monkeypox.(Realegeno et al., 2017)

According to Smee DF. et al., Antiviral agents are being sought to treat smallpox (live-attenuated vaccinia virus) vaccination-related complications or unintentional infections brought on by contact with vaccinated people. They are also being sought as preventative measures against the possible deliberate release of the monkeypox and smallpox viruses. Two substances that are of great interest are cidofovir and tecovirimat. (Smee, 2013)

Cidofovir can be used to treat monkeypox virus infections, according to Rice A.D. et al. Additionally, it can treat the side effects of smallpox vaccination. (Rice et al., 2011)

Infections caused by the monkeypox virus can be treated with cidofovir, according to Shao et al. Furthermore, the negative effects of receiving a smallpox vaccination can be treated with it. (Shao et al., 2009)

Schultz DA proposed a dormouse model. et al. and validated for testing both therapeutic (cidofovir) and prophylactic (Dryvax vaccine) test articles against intranasal challenges with the monkeypox virus.(Schultz et al., 2009)

Wei H. et al. investigated the effects of a single dose of cidofovir and Dryvax co-administration in reducing the adverse effects of vaccination while significantly impairing the immune responses and immunity to Monkeypox induced by the vaccine. (Wei et al., 2009)

According to research by Rivera R. et al., The severity of infection in mice is made worse by liposomal clodronate depletion of alveolar macrophages, leading to increased replication and systemic spread of vaccinia.(Rivera et al., 2007)

### Prescription drug information

Some prescription drug information is employed to investigate drug interactions, drug and foods completions, side effects, and serious conditions concerns

Several reputable websites, such as Medscape^7^, WebMD^8^, Drugs^9^, and Drugbank,^10^ have been used to investigate drug interactions. In all of these databases, the drugs were tested in pairs.

The drugs were examined in pairs among the Internet databases for identifying drug interactions, such as Medscape, WebMD, Drugs, and Drugbank. According to the information provided by these databases, we found two pair interactions: Tecovirimat-smallpox vaccine and Cidofovir-Clodronic Acid.

#### Drug interactions

According to Medscape, Tecovirimat and the smallpox vaccine should be used cautiously/monitored. There have been no human studies on vaccine-drug interactions. According to some animal studies, administering tecovirimat and the smallpox vaccine may reduce the immune response. The effect of this interaction on vaccine efficacy in the clinical setting is unknown.

Drugbank claims that Combining Cidofovir and Clodronic acid can increase the risk or severity of nephrotoxicity and hypocalcemia. The mild, asymptomatic hypocalcemia that bisphosphonates present on their own (Do et al., 2012). Nephrotoxic substances may enhance the hypocalcemic effects of bisphosphonates. Both substances can cause hypocalcemia by various mechanisms, and these effects are cumulative. Although the nephrotoxic potential of bisphosphonates is not well understood, renal damage may worsen when combined with other nephrotoxic substances.

## CONCLUSION

This study proposed a combination of Tecovirimat, Smallpox vaccine, bafilomycin A1, Cidofovir, and Clodronic Acid to treat Monkeypox. The p-value between Monkeypox and the human genes associated with this combination has reached 0.015, which indicates that this drug combination is very effective. The effectiveness and safety of other antiviral treatments still have controversy and require more high-quality clinical trials.

## Data Availability

Not applicable

## Abbreviations

STROBE: Strengthening the Reporting of Observational Studies in Epidemiology;
PRISMA: Preferred Reporting Items for Systematic Reviews and Meta-Analysis
RAIN: Systematic Review and Artificial Intelligence Network Meta-Analysis

## Acknowledgments

This research is performed by Student researchers in Bioinformatics, Computational Biology, Machine Learning, and Medicine.

## Authors’ contributions

N.S. contributed to the design, AAK and M.B. native machine learning algorithm, MM, AAK, MB, and D.S. bio-statistical analysis; S.A. participated in most of the study steps. S.A. and A.F. used machine learning in exploring drug combinations. All authors have read and approved the content of the manuscript.

## Availability of data and materials

Datasets are available through the corresponding author upon reasonable request.

## Ethics approval and consent to participate

Not applicable.

## Consent for publication

Not applicable.

## Footnotes

7 https://reference.medscape.com/drug-interactionchecker

8 https://www.webmd.com/interaction-checker/default.htm

9 https://www.drugs.com/drug_interactions.html

10 https://go.drugbank.com/drug-interaction-checker

