## Supplementary material for "Drug combinations proposed by machine learning on genes/proteins to improve the efficacy of Tecovirimat in the treatment of Monkeypox: A Systematic Review and Network Meta-analysis": Assoc.v32_supplementary_material_2.pdf

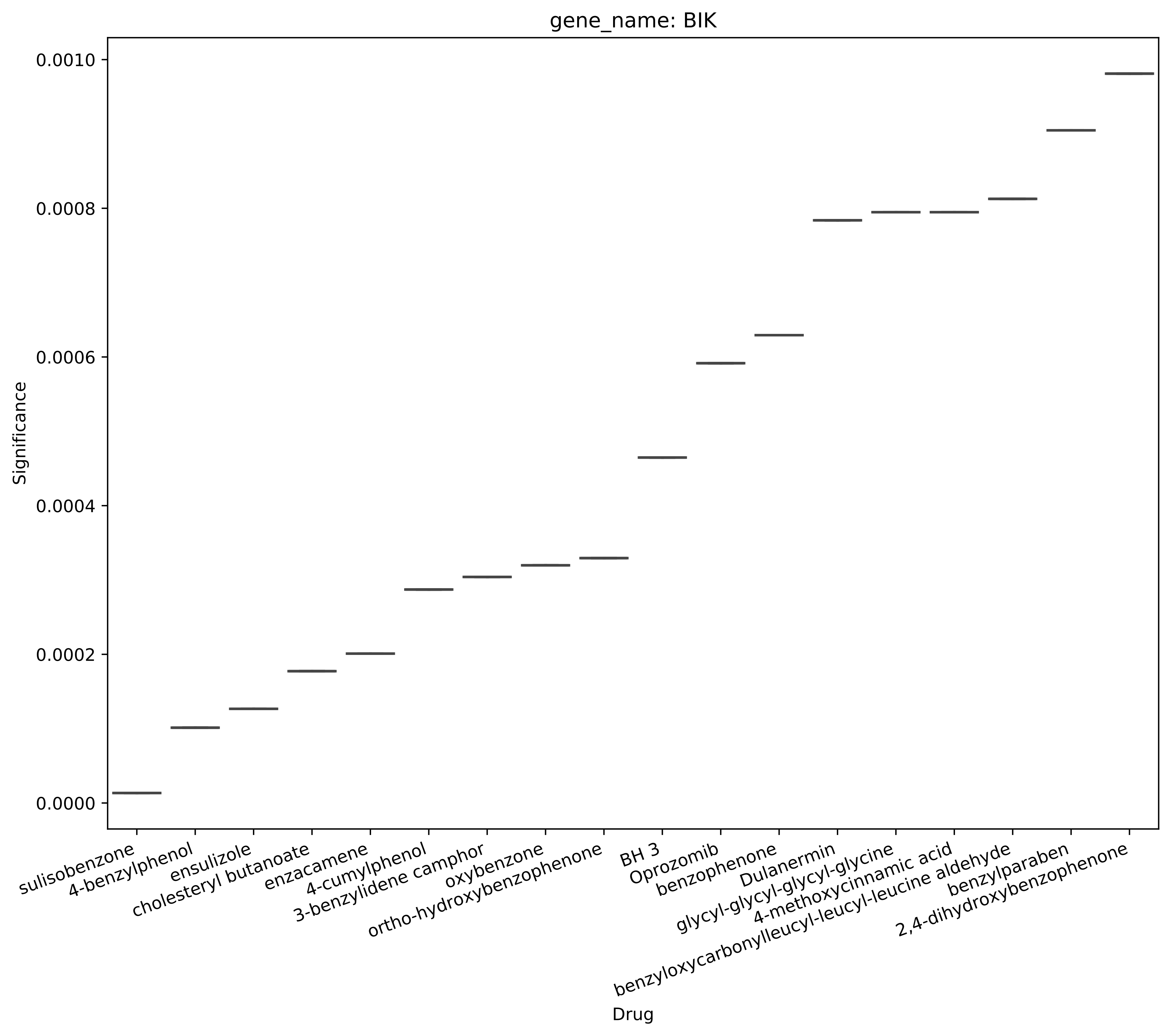
gene_name: BIK
Significance
0.0010
0.0008
0.0006
0.0004
0.0002
0.0000
Drug
sulisobenzone
4-benzylphenol
ensulizole
cholesteryl butanoate
enzacamene
4-cumylphenol
3-benzylidene camphor
oxybenzone
ortho-hydroxybenzophenone
BH 3
Oprozomib
benzophenone
Dulanermin
glycyl-glycyl-glycyl-glycine
4-methoxycinnamic acid
benzyloxycarbonylleucyl-leucyl-leucine aldehyde
benzylparaben
2,4-dihydroxybenzophenone

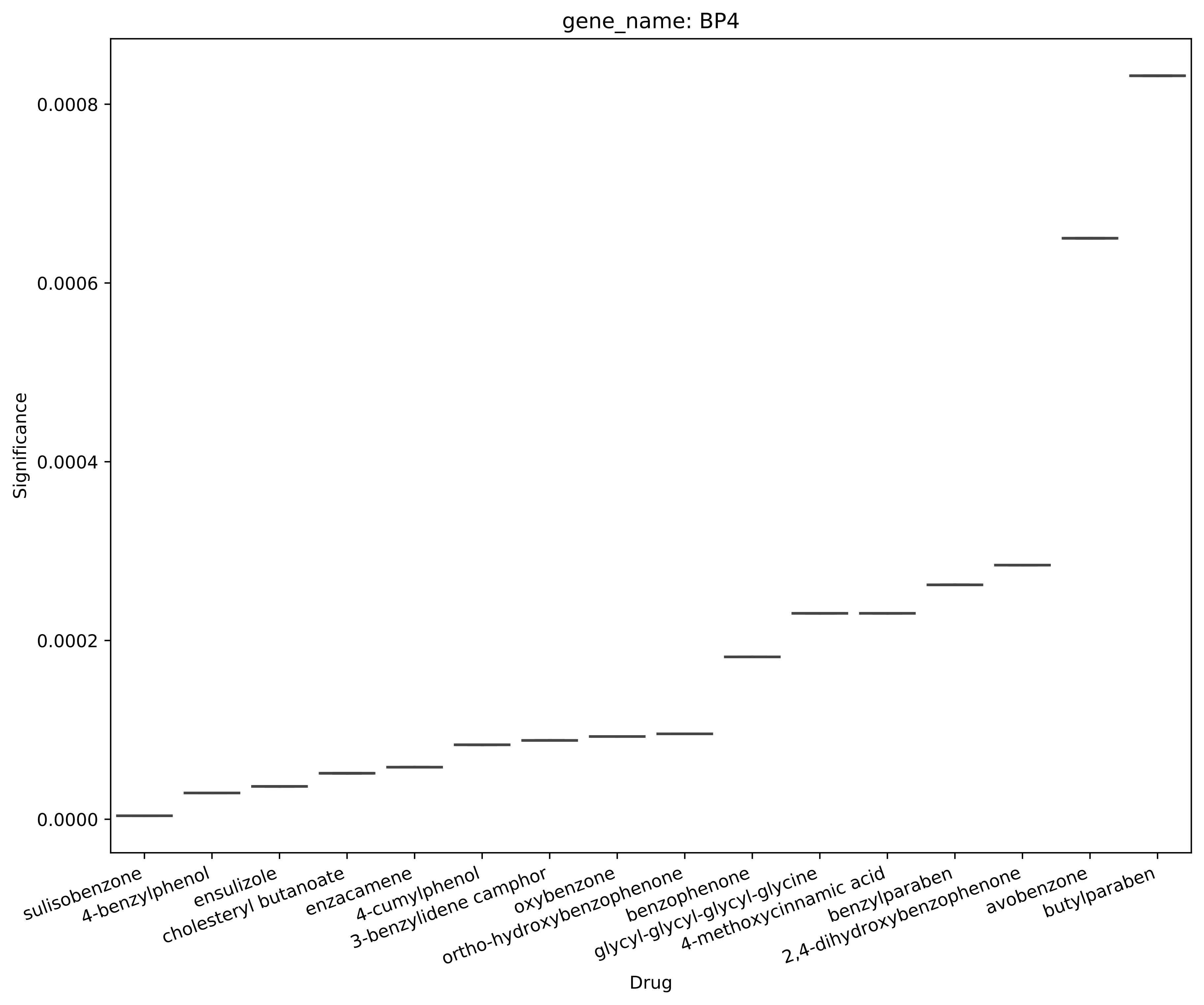
gene_name: BP4
Significance
0.0000
0.0002
0.0004
0.0006
0.0008
Drug
sulisobenzone
4-benzylphenol
ensulizole
cholesteryl butanoate
enzacamene
4-cumylphenol
3-benzylidene camphor
oxybenzone
ortho-hydroxybenzophenone
benzophenone
glycyl-glycyl-glycyl-glycine
4-methoxycinnamic acid
benzylparaben
2,4-dihydroxybenzophenone
avobenzone
butylparaben

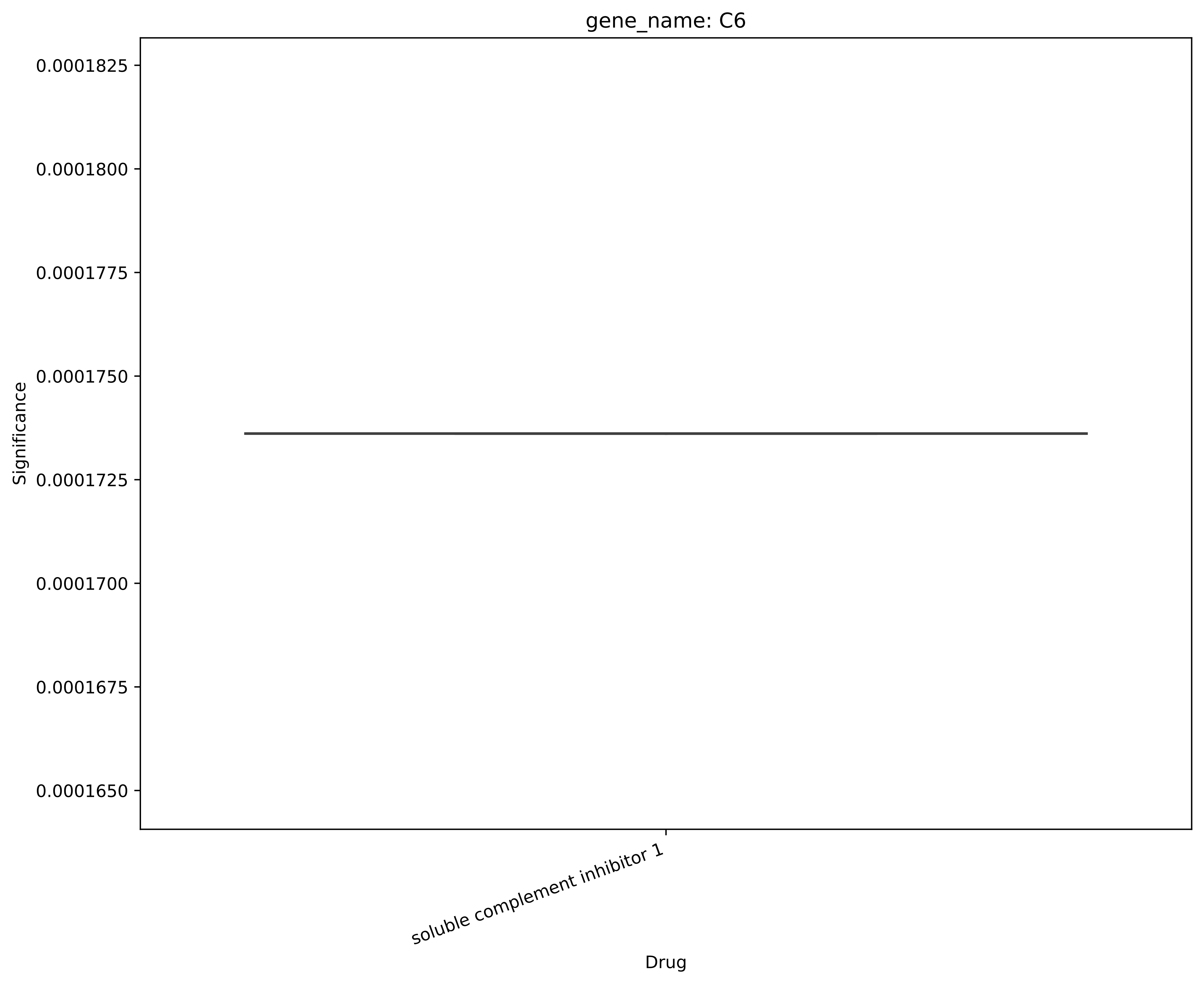
gene_name: C6
Significance
0.0001825
0.0001800
0.0001775
0.0001750
0.0001725
0.0001700
0.0001675
0.0001650
soluble complement inhibitor 1
Drug

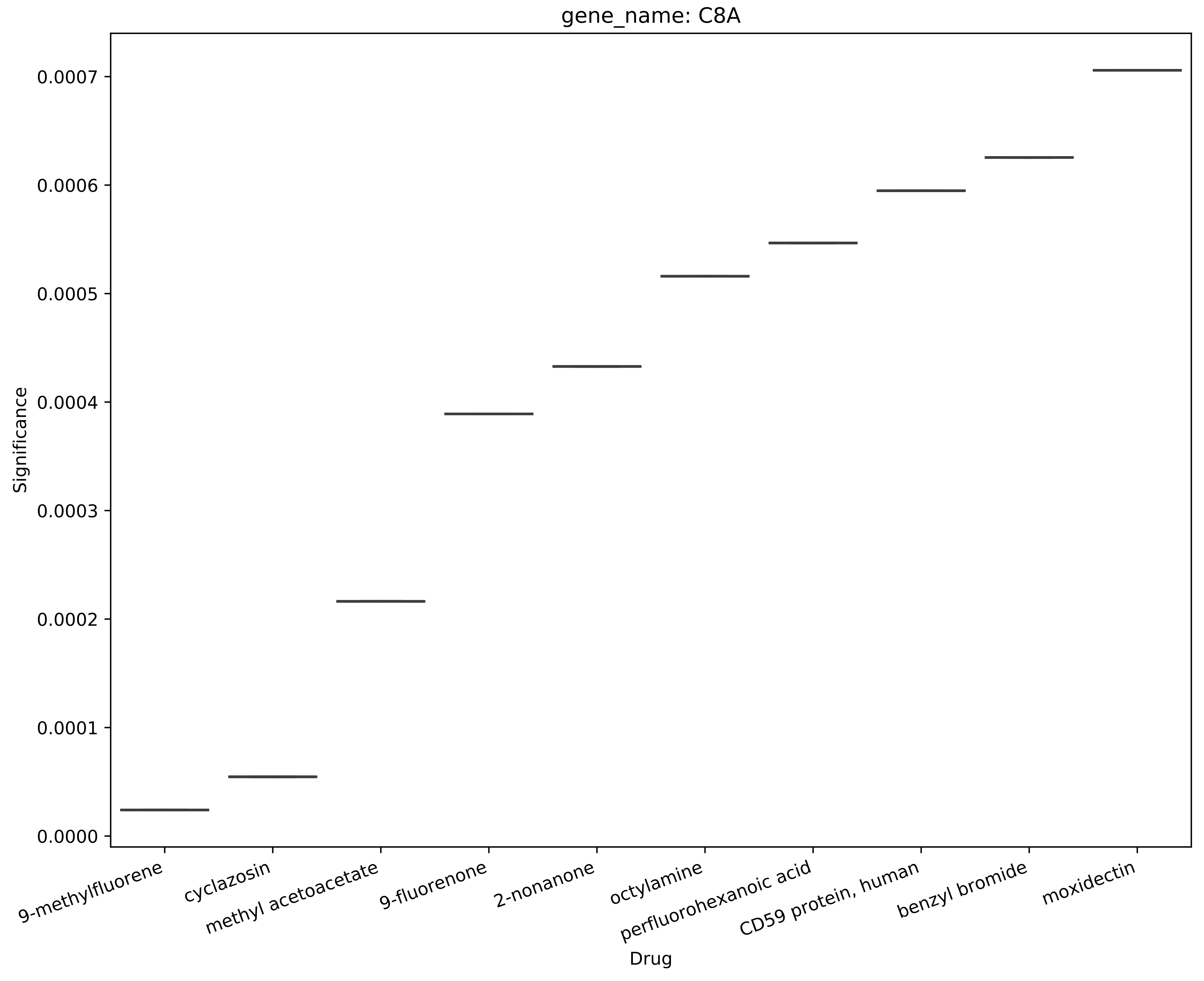
gene_name: C8A
Significance
0.0000
0.0001
0.0002
0.0003
0.0004
0.0005
0.0006
0.0007
9-methylfluorene
cyclazosin
methyl acetoacetate
9-fluorenone
2-nonanone
octylamine
perfluorohexanoic acid
CD59 protein, human
benzyl bromide
moxidectin
Drug

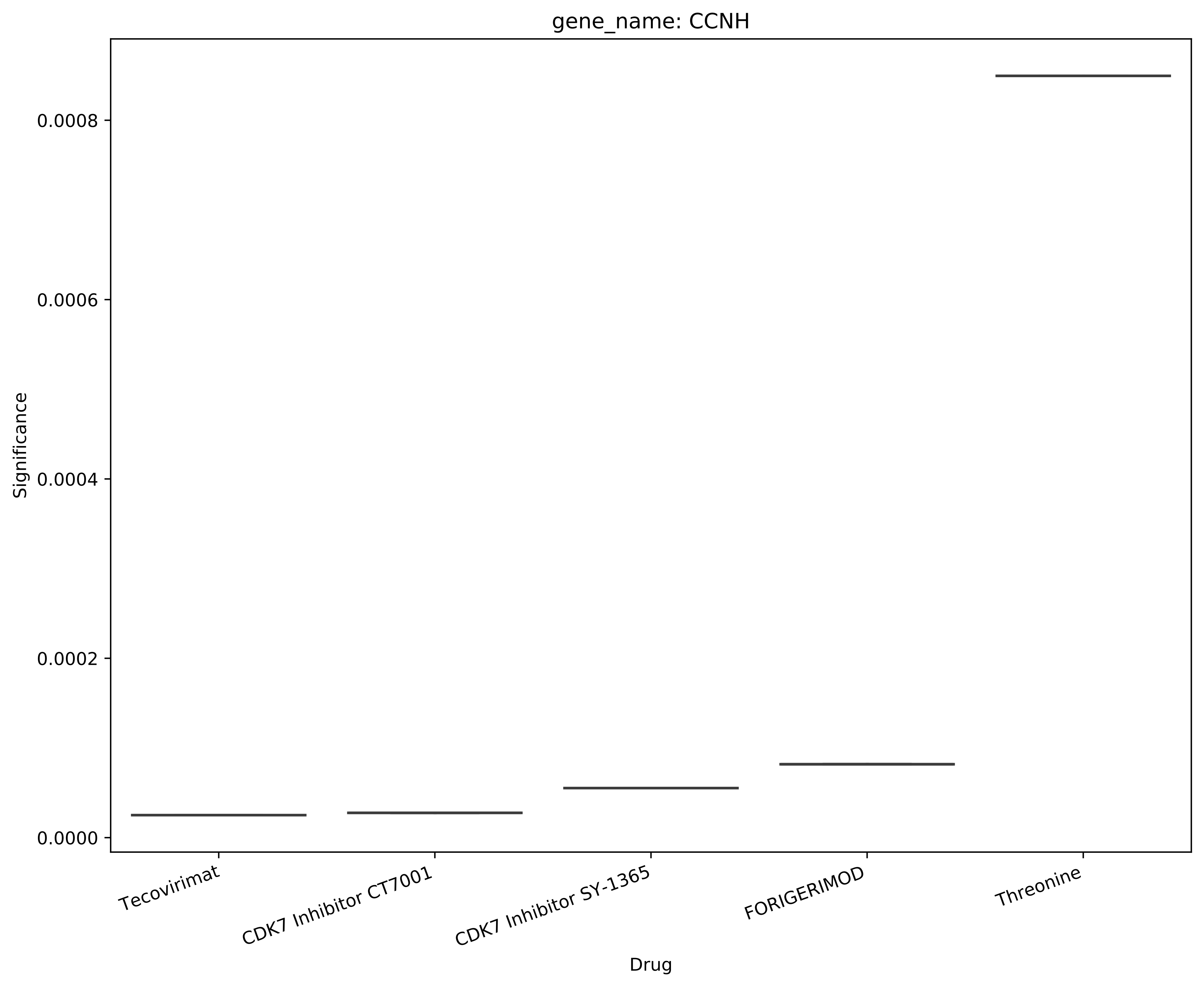
gene_name: CCNH
Significance
0.0008
0.0006
0.0004
0.0002
0.0000
Tecovirimat
CDK7 Inhibitor CT7001
CDK7 Inhibitor SY-1365
FORIGERIMOD
Threonine
Drug

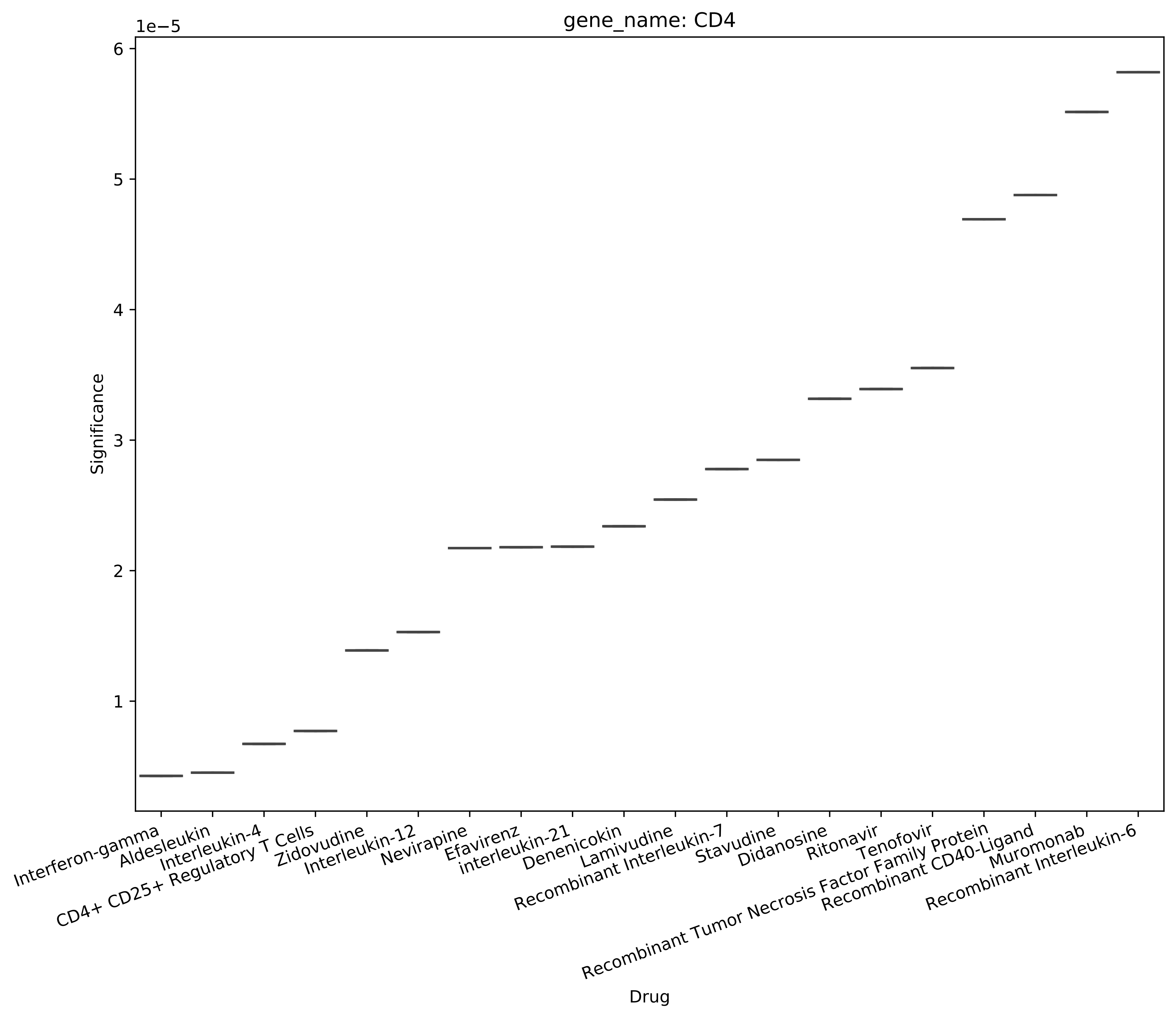

gene_name: CD4
1e−5
6
5
4
3
2
1
Significance
Interferon-gamma
Aldesleukin
Interleukin-4
CD4+ CD25+ Regulatory T Cells
Zidovudine
Interleukin-12
Nevirapine
Efavirenz
interleukin-21
Denenicokin
Lamivudine
Recombinant Interleukin-7
Stavudine
Didanosine
Ritonavir
Tenofovir
Recombinant Tumor Necrosis Factor Family Protein
Recombinant CD40-Ligand
Muromonab
Recombinant Interleukin-6
Drug

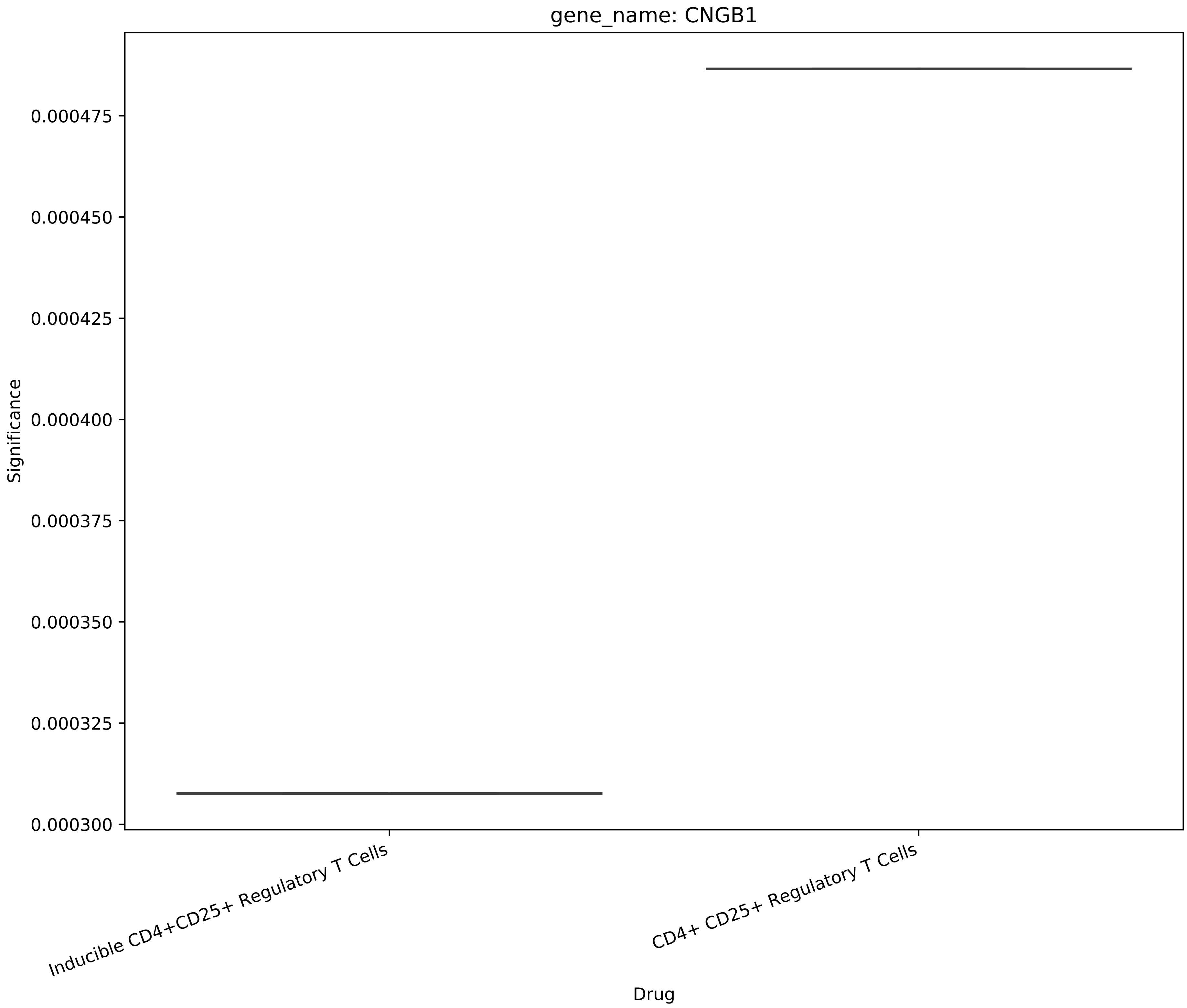
gene_name: CNGB1
Significance
0.000475
0.000450
0.000425
0.000400
0.000375
0.000350
0.000325
0.000300
Inducible CD4+CD25+ Regulatory T Cells
CD4+ CD25+ Regulatory T Cells
Drug

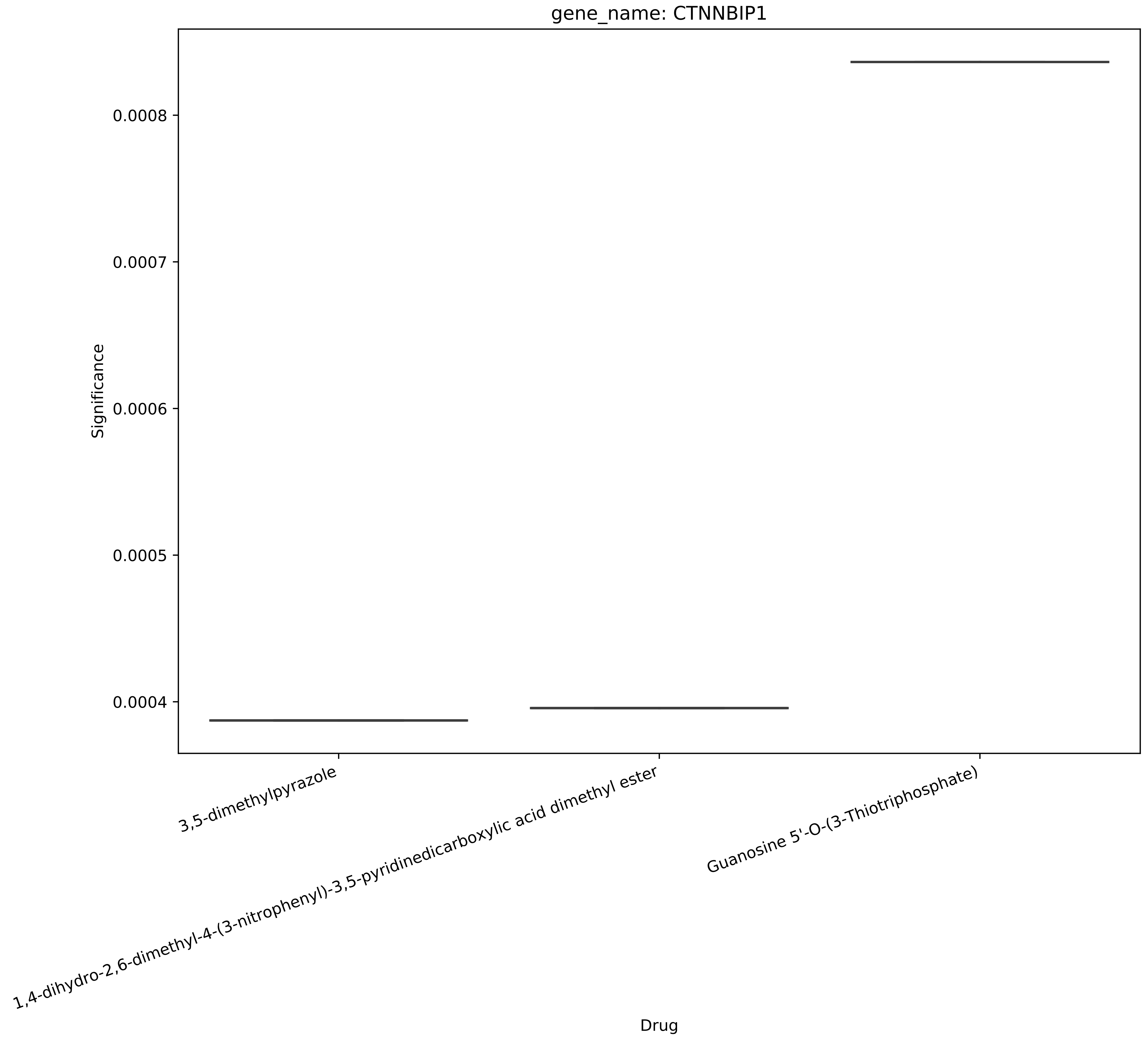
gene_name: CTNNBIP1
Significance
0.0008
0.0007
0.0006
0.0005
0.0004
3,5-dimethylpyrazole
1,4-dihydro-2,6-dimethyl-4-(3-nitrophenyl)-3,5-pyridinedicarboxylic acid dimethyl ester
Guanosine 5'-O-(3-Thiotriphosphate)
Drug

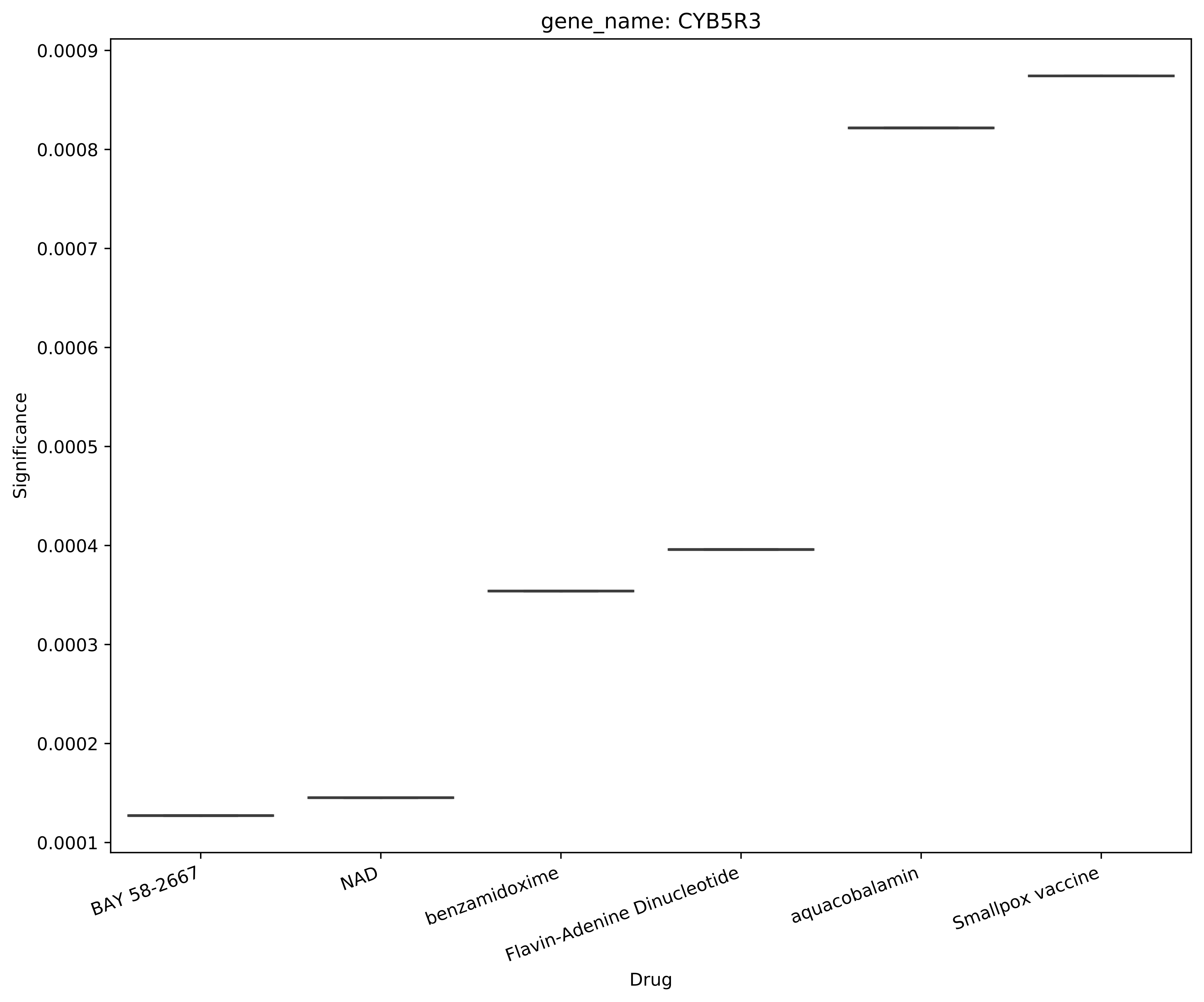
gene_name: CYB5R3
Significance
0.0009
0.0008
0.0007
0.0006
0.0005
0.0004
0.0003
0.0002
0.0001
BAY 58-2667
NAD
benzamidoxime
Flavin-Adenine Dinucleotide
aquacobalamin
Smallpox vaccine
Drug

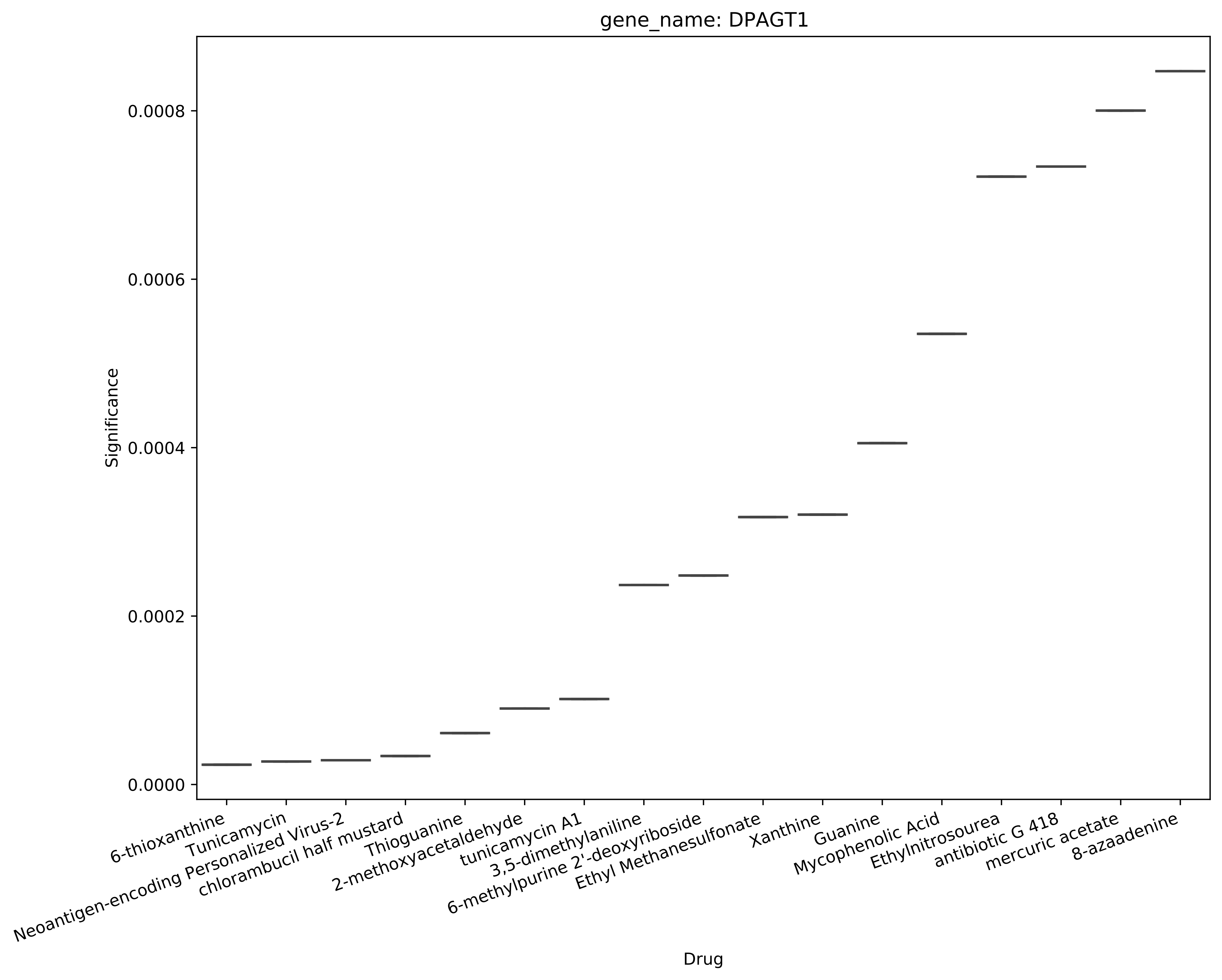

gene_name: DPAGT1
Significance
0.0000
0.0002
0.0004
0.0006
0.0008
Drug
6-thioxanthine
Tunicamycin
Neoantigen-encoding Personalized Virus-2
chlorambucil half mustard
Thioguanine
2-methoxyacetaldehyde
tunicamycin A1
3,5-dimethylaniline
6-methylpurine 2'-deoxyriboside
Ethyl Methanesulfonate
Xanthine
Guanine
Mycophenolic Acid
Ethylnitrosourea
antibiotic G 418
mercuric acetate
8-azaadenine

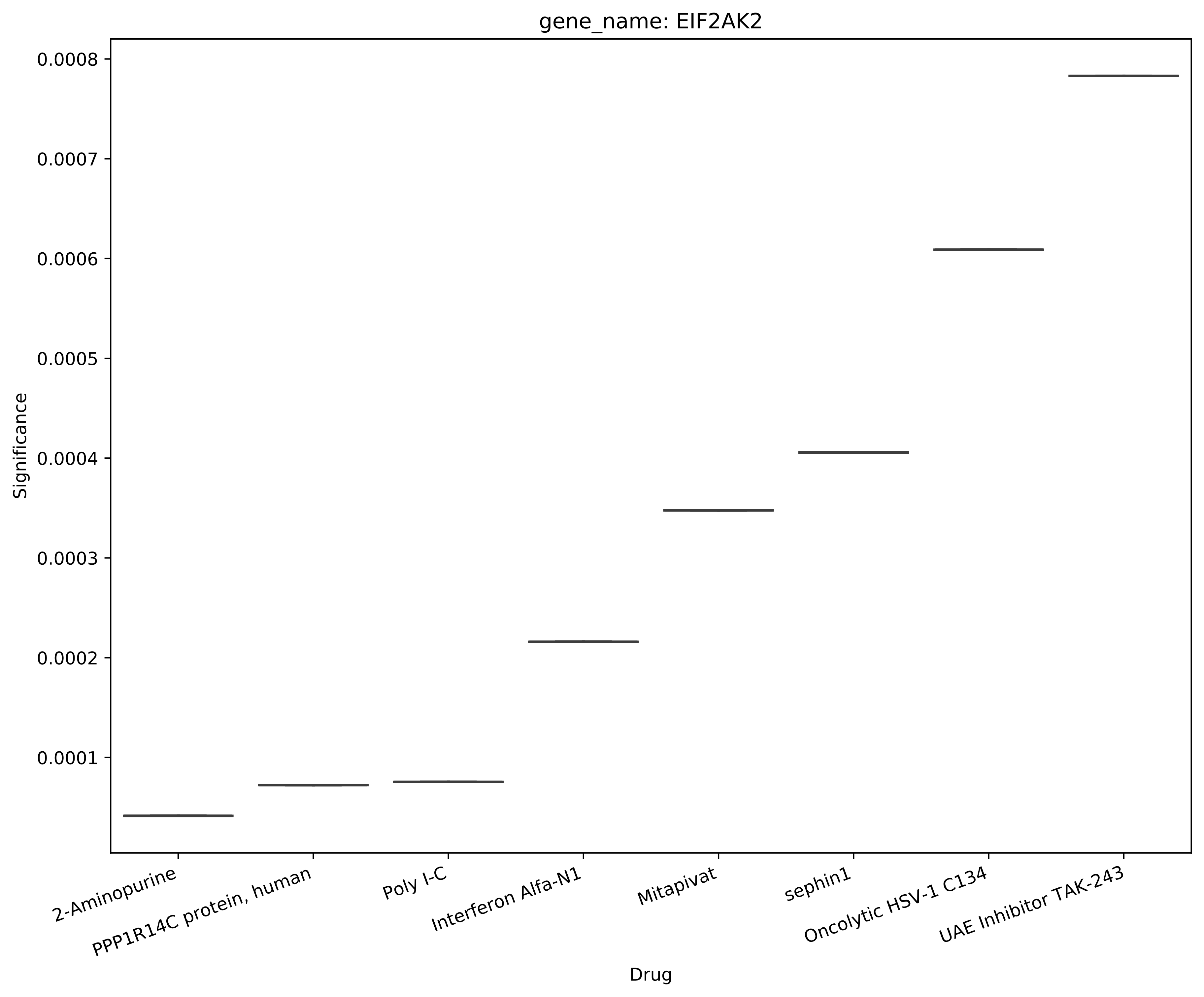

gene_name: EIF2AK2
Significance
0.0001
0.0002
0.0003
0.0004
0.0005
0.0006
0.0007
0.0008
2-Aminopurine
PPP1R14C protein, human
Poly I-C
Interferon Alfa-N1
Mitapivat
sephin1
Oncolytic HSV-1 C134
UAE Inhibitor TAK-243
Drug

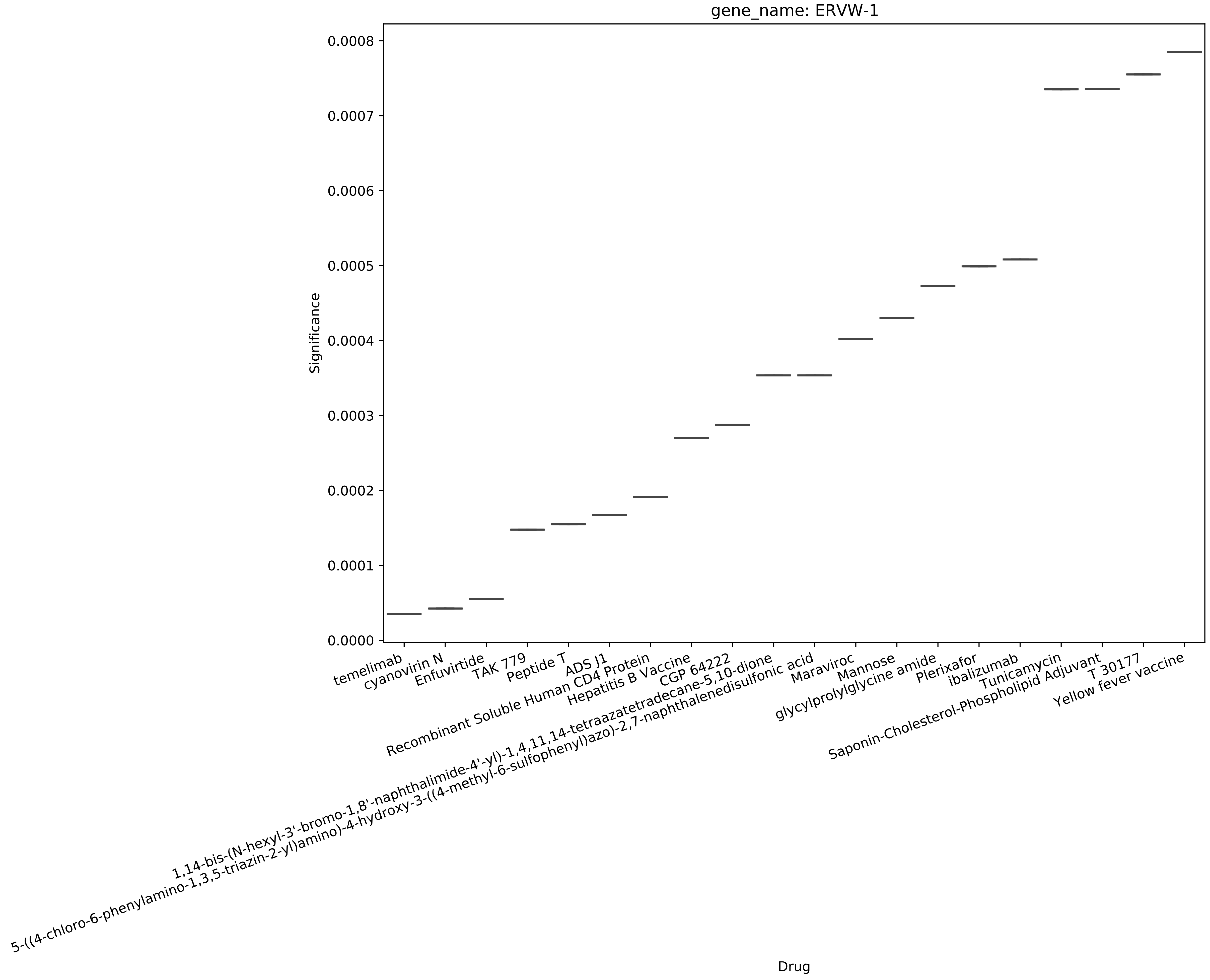
gene_name: ERVW-1
Significance
0.0000
0.0001
0.0002
0.0003
0.0004
0.0005
0.0006
0.0007
0.0008
Drug
temelimab
cyanovirin N
Enfuvirtide
TAK 779
Peptide T
ADS J1
Recombinant Soluble Human CD4 Protein
Hepatitis B Vaccine
CGP 64222
1,14-bis-(N-hexyl-3'-bromo-1,8'-naphthalimide-4'-yl)-1,4,11,14-tetraazatetradecane-5,10-dione
5-((4-chloro-6-phenylamino-1,3,5-triazin-2-yl)amino)-4-hydroxy-3-((4-methyl-6-sulfophenyl)azo)-2,7-naphthalenedisulfonic acid
Maraviroc
Mannose
glycylprolylglycine amide
Plerixafor
ibalizumab
Tunicamycin
Saponin-Cholesterol-Phospholipid Adjuvant
T 30177
Yellow fever vaccine

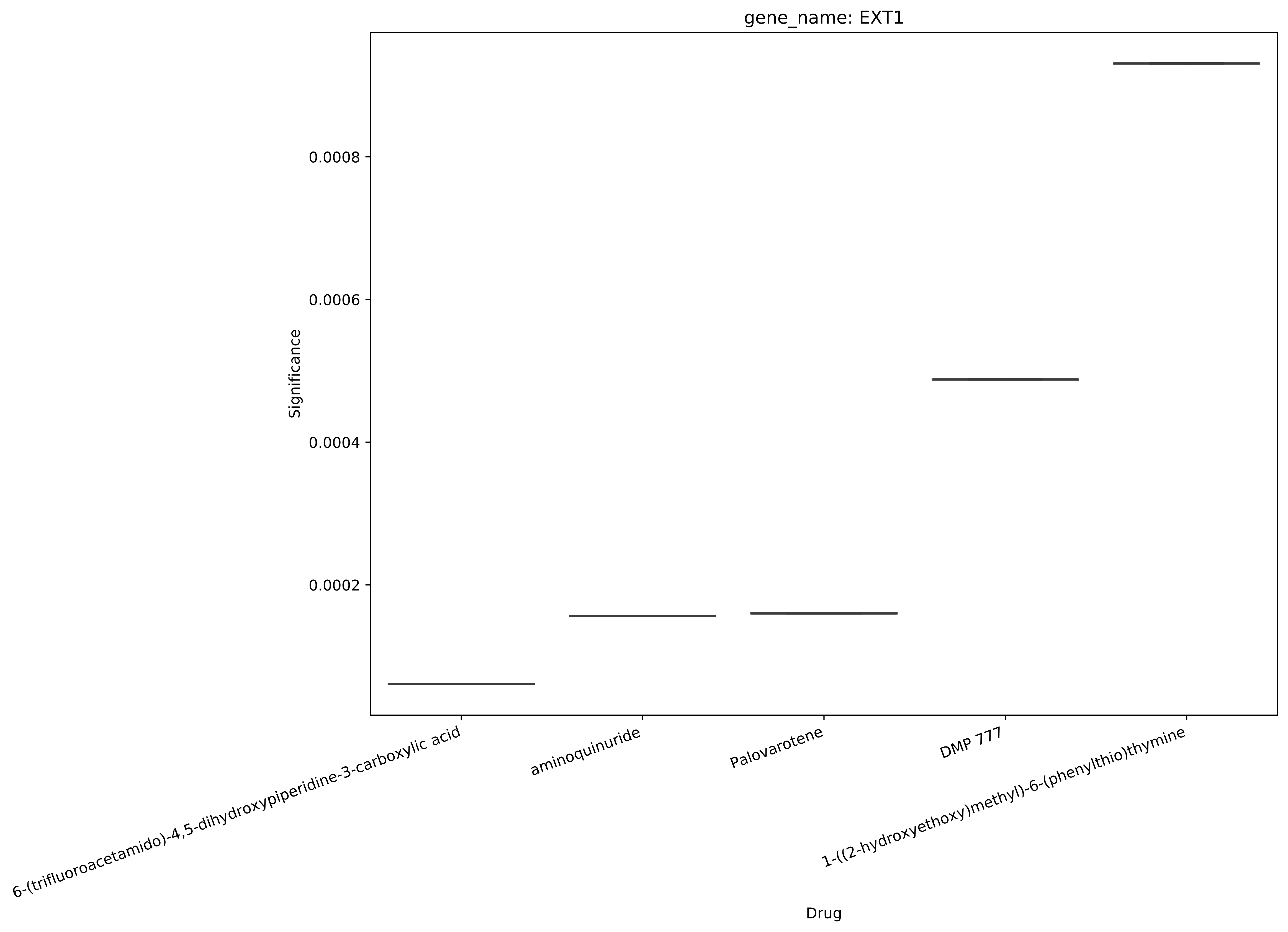
gene_name: EXT1
Significance
0.0002
0.0004
0.0006
0.0008
Drug
6-(trifluoroacetamido)-4,5-dihydroxypiperidine-3-carboxylic acid
aminoquinuride
Palovarotene
DMP 777
1-((2-hydroxyethoxy)methyl)-6-(phenylthio)thymine

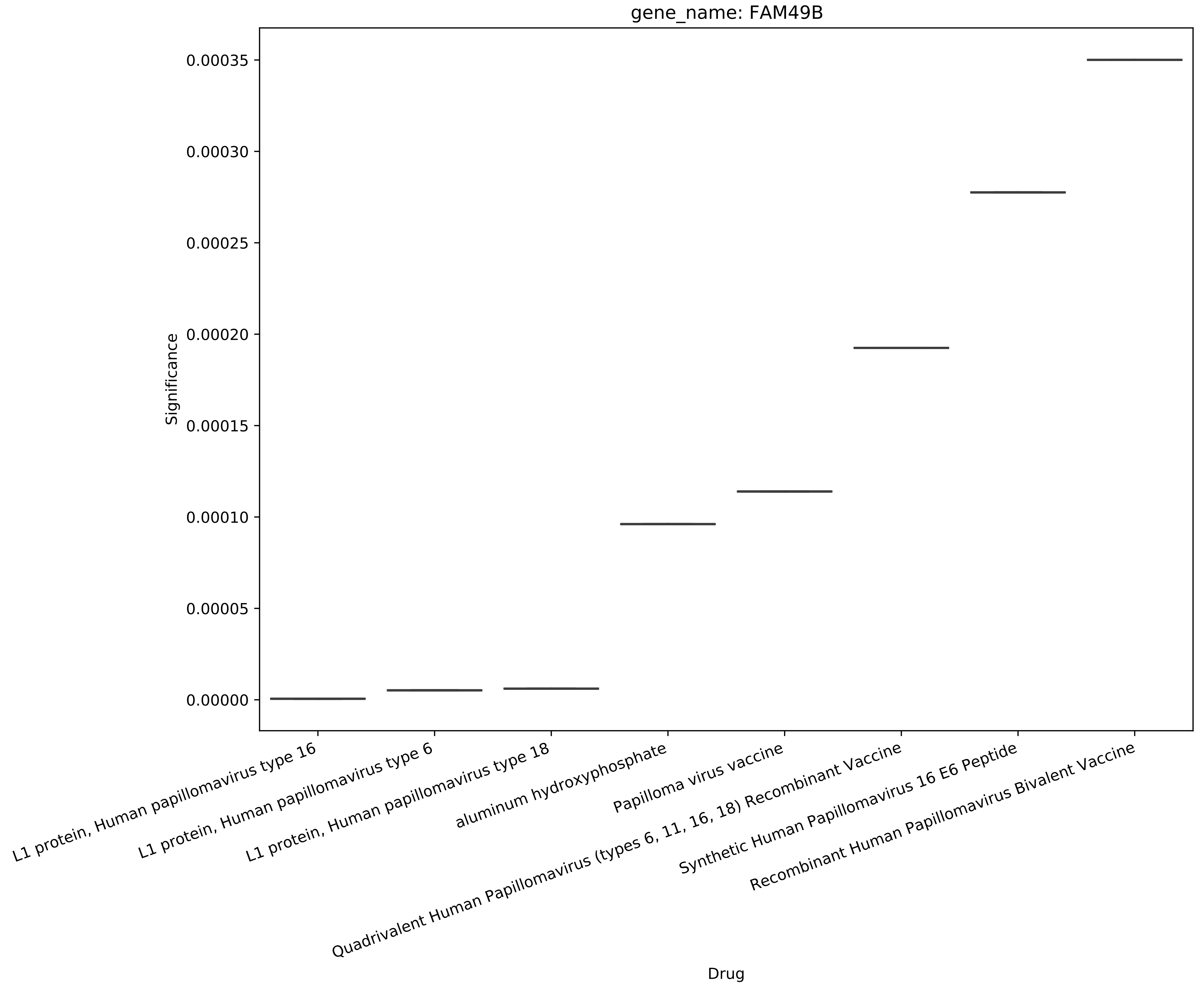
gene_name: FAM49B
Significance
0.00035
0.00030
0.00025
0.00020
0.00015
0.00010
0.00005
0.00000
L1 protein, Human papillomavirus type 16
L1 protein, Human papillomavirus type 6
L1 protein, Human papillomavirus type 18
aluminum hydroxyphosphate
Papilloma virus vaccine
Quadrivalent Human Papillomavirus (types 6, 11, 16, 18) Recombinant Vaccine
Synthetic Human Papillomavirus 16 E6 Peptide
Recombinant Human Papillomavirus Bivalent Vaccine
Drug

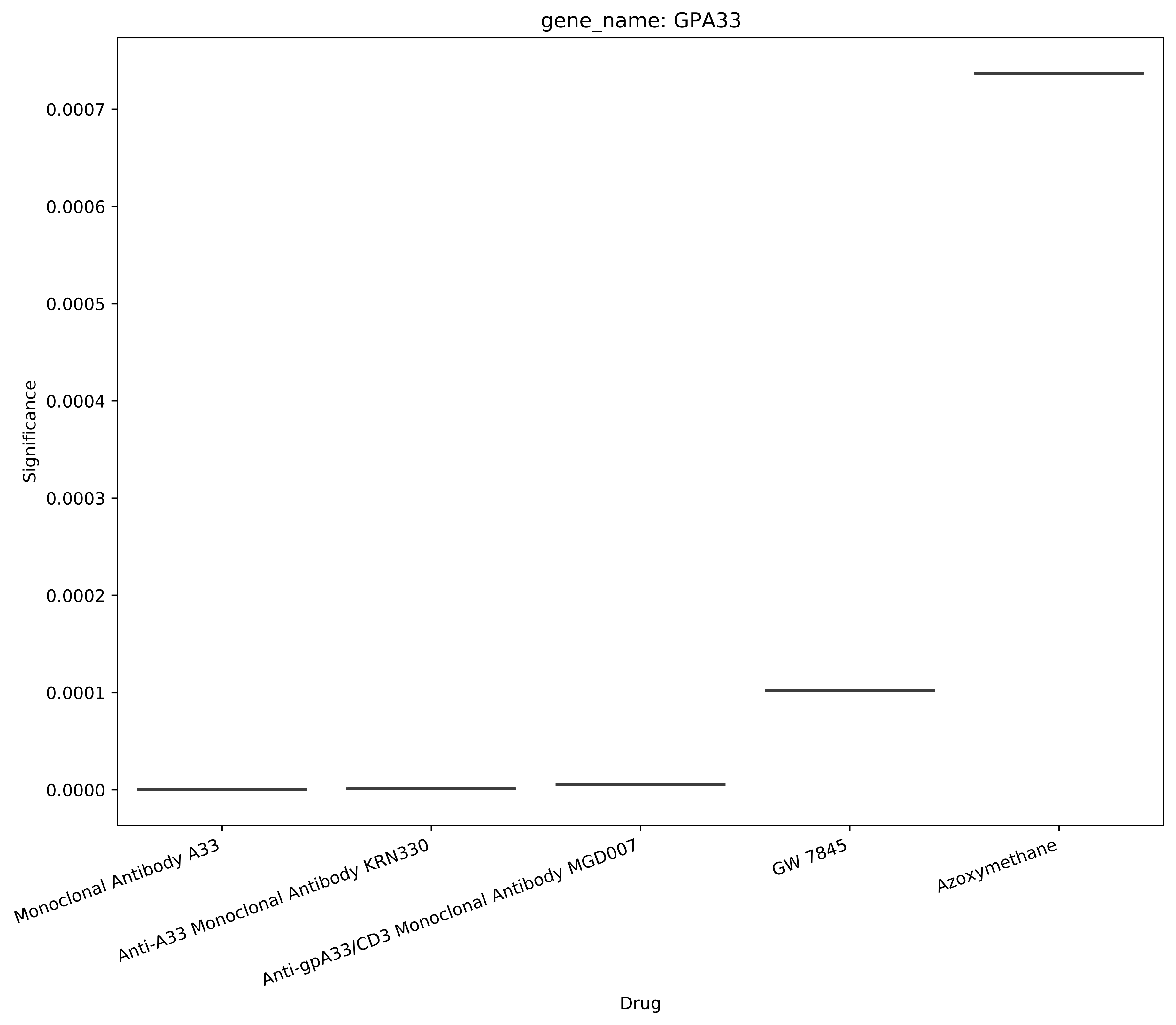
gene_name: GPA33
Significance
0.0007
0.0006
0.0005
0.0004
0.0003
0.0002
0.0001
0.0000
Monoclonal Antibody A33
Anti-A33 Monoclonal Antibody KRN330
Anti-gpA33/CD3 Monoclonal Antibody MGD007
GW 7845
Azoxymethane
Drug

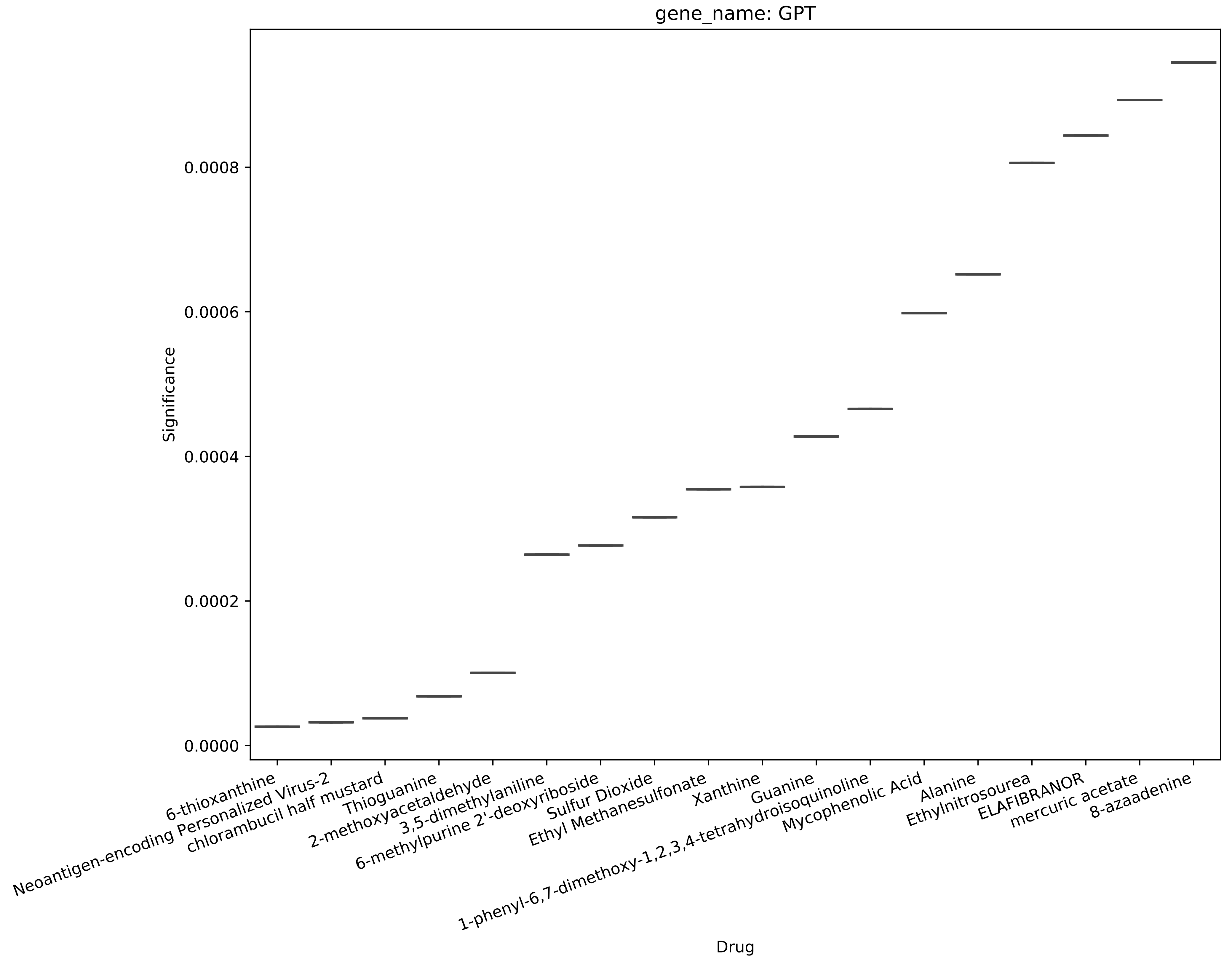

gene_name: GPT
Significance
0.0000
0.0002
0.0004
0.0006
0.0008
Drug
6-thioxanthine
Neoantigen-encoding Personalized Virus-2
chlorambucil half mustard
Thioguanine
2-methoxyacetaldehyde
3,5-dimethylaniline
6-methylpurine 2'-deoxyriboside
Sulfur Dioxide
Ethyl Methanesulfonate
Xanthine
1-phenyl-6,7-dimethoxy-1,2,3,4-tetrahydroisoquinoline
Guanine
Mycophenolic Acid
Alanine
Ethylnitrosourea
ELAFIBRANOR
mercuric acetate
8-azaadenine

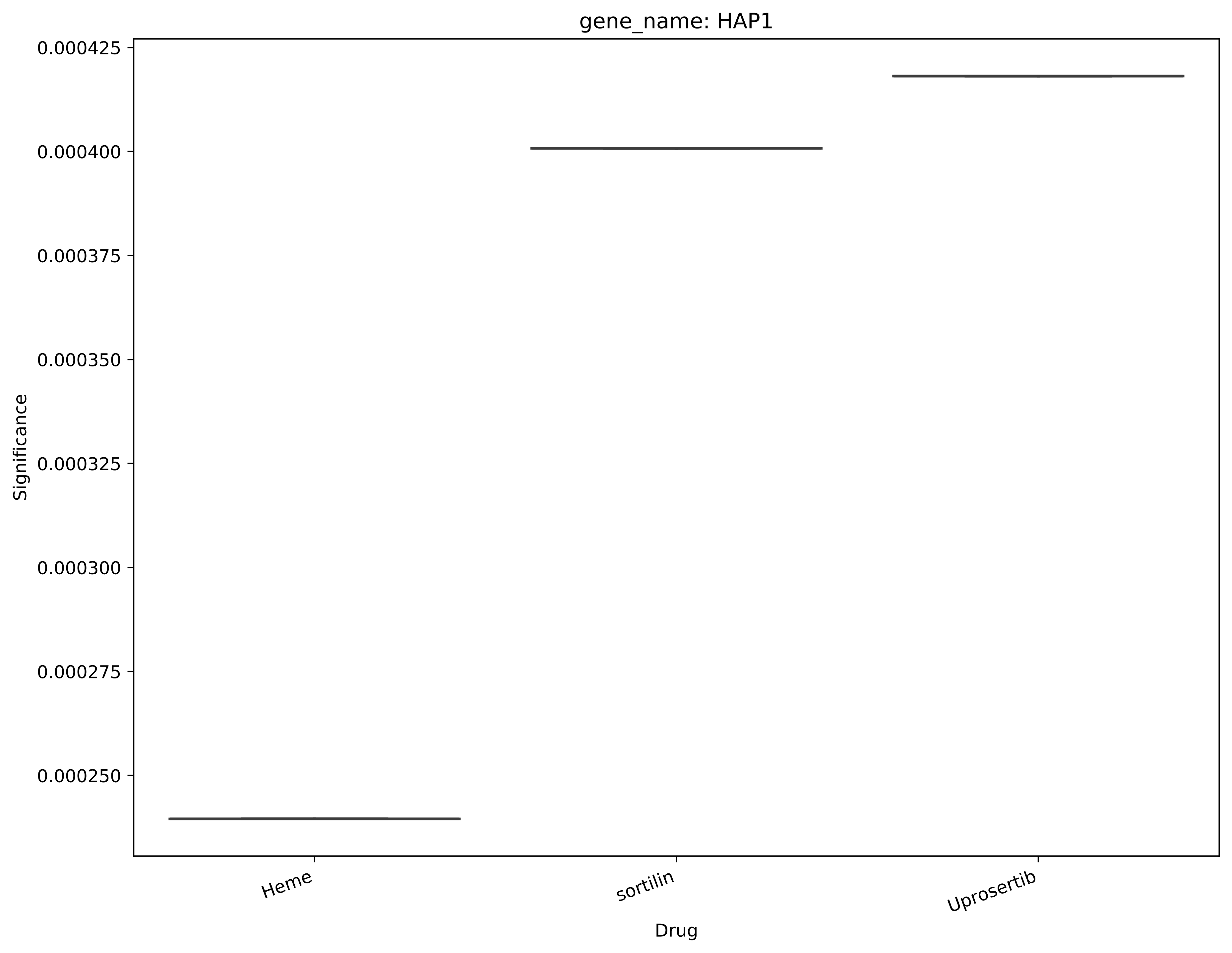

gene_name: HAP1
0.000425
0.000400
0.000375
0.000350
0.000325
0.000300
0.000275
0.000250
Significance
Heme
sortilin
Uprosertib
Drug

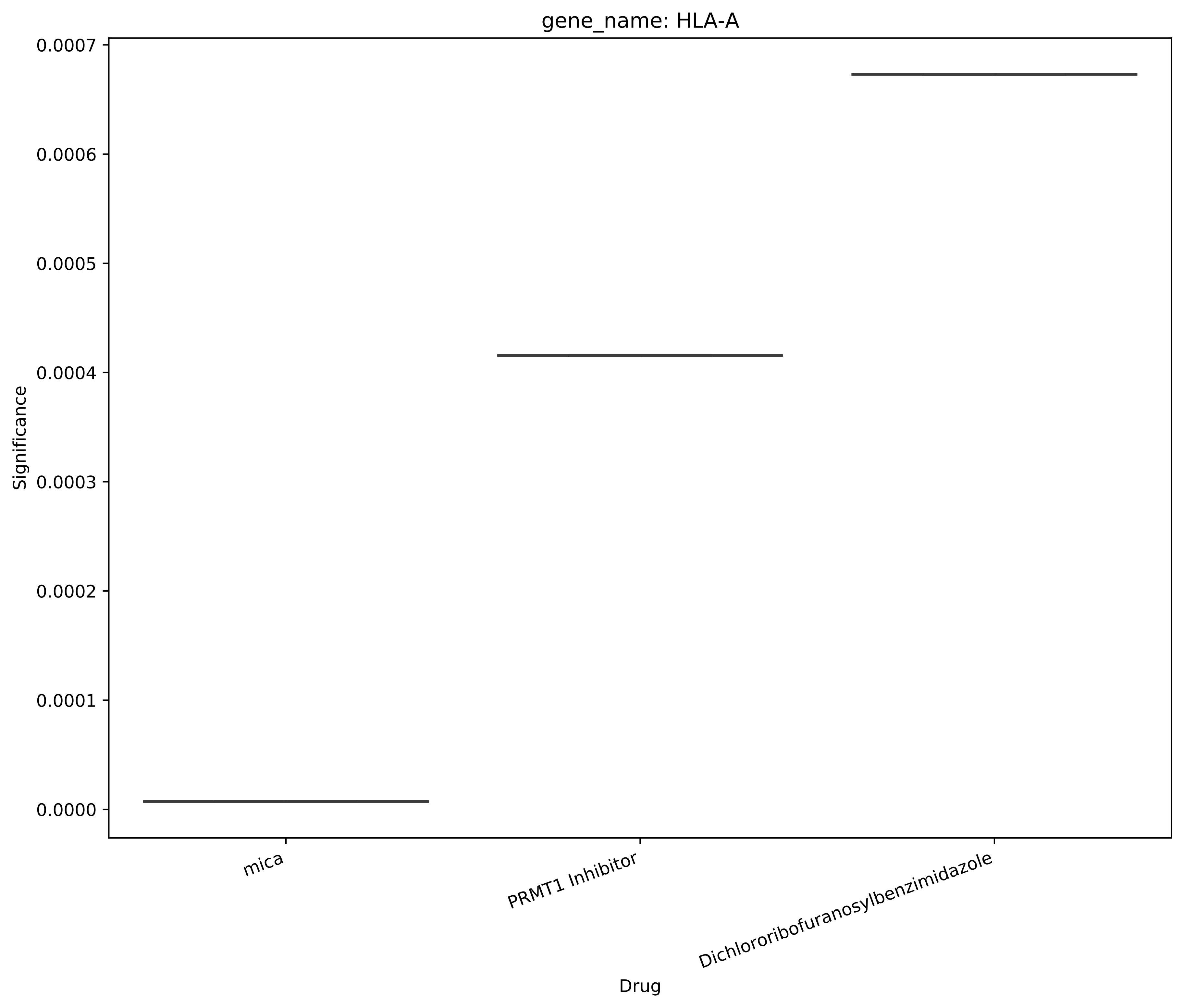
gene_name: HLA-A
Significance
0.0007
0.0006
0.0005
0.0004
0.0003
0.0002
0.0001
0.0000
mica
PRMT1 Inhibitor
Dichlororibofuranosylbenzimidazole
Drug

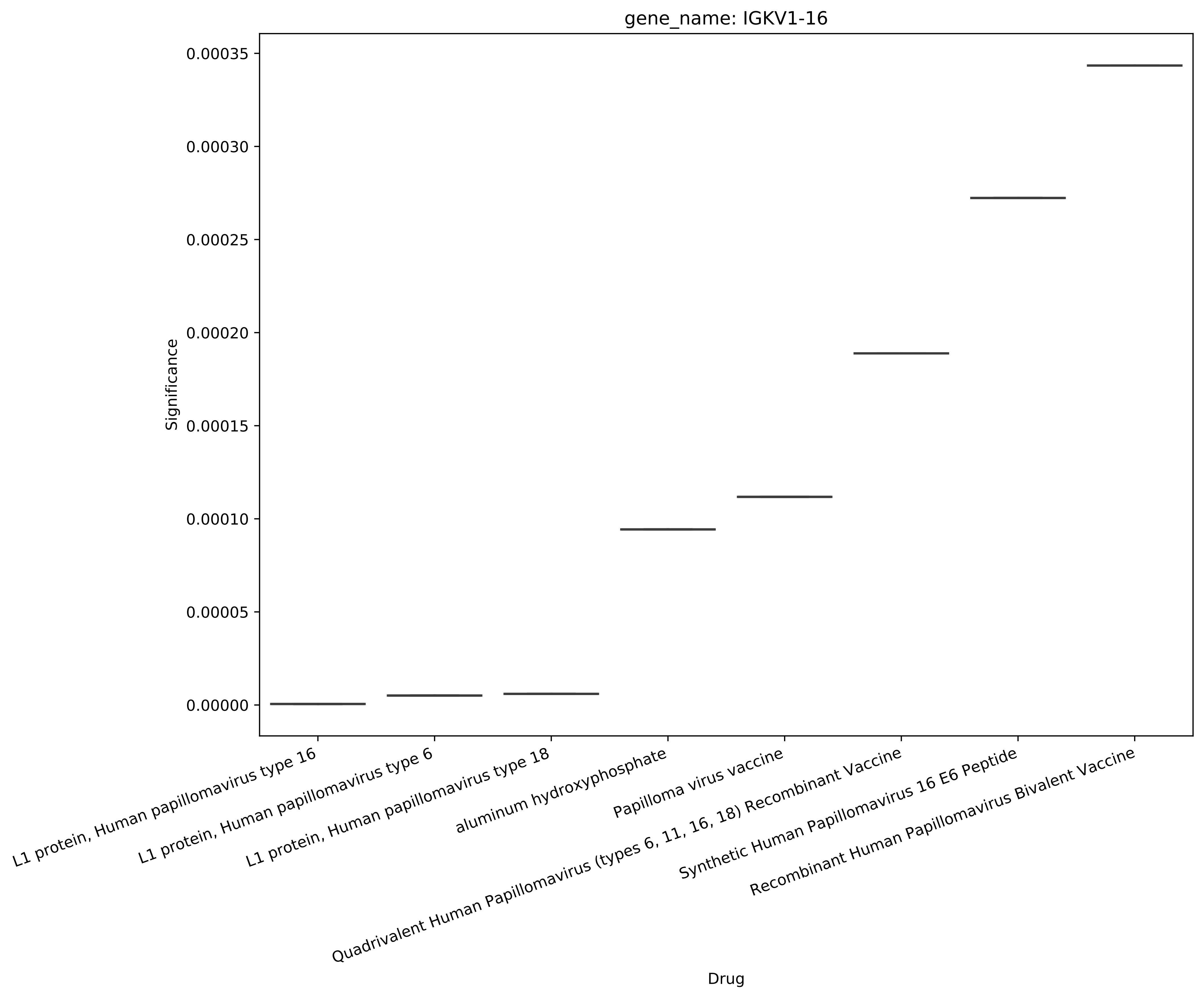

gene_name: IGKV1-16
Significance
0.00035
0.00030
0.00025
0.00020
0.00015
0.00010
0.00005
0.00000
L1 protein, Human papillomavirus type 16
L1 protein, Human papillomavirus type 6
L1 protein, Human papillomavirus type 18
aluminum hydroxyphosphate
Papilloma virus vaccine
Quadrivalent Human Papillomavirus (types 6, 11, 16, 18) Recombinant Vaccine
Synthetic Human Papillomavirus 16 E6 Peptide
Recombinant Human Papillomavirus Bivalent Vaccine
Drug

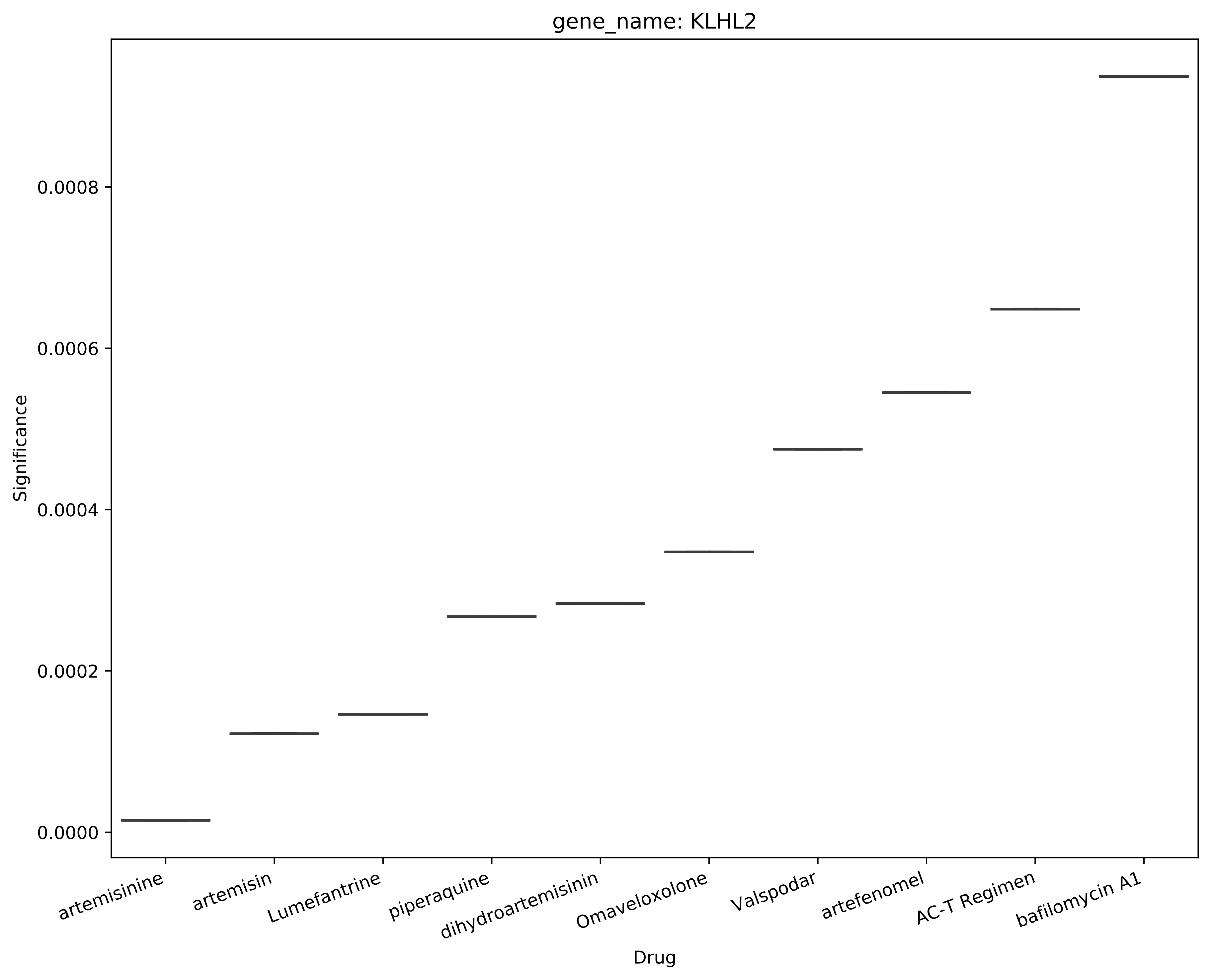
gene_name: KLHL2
Significance
0.0000
0.0002
0.0004
0.0006
0.0008
artemisinine
artemisin
Lumefantrine
piperaquine
dihydroartemisinin
Omaveloxolone
Valspodar
artefenomel
AC-T Regimen
bafilomycin A1
Drug

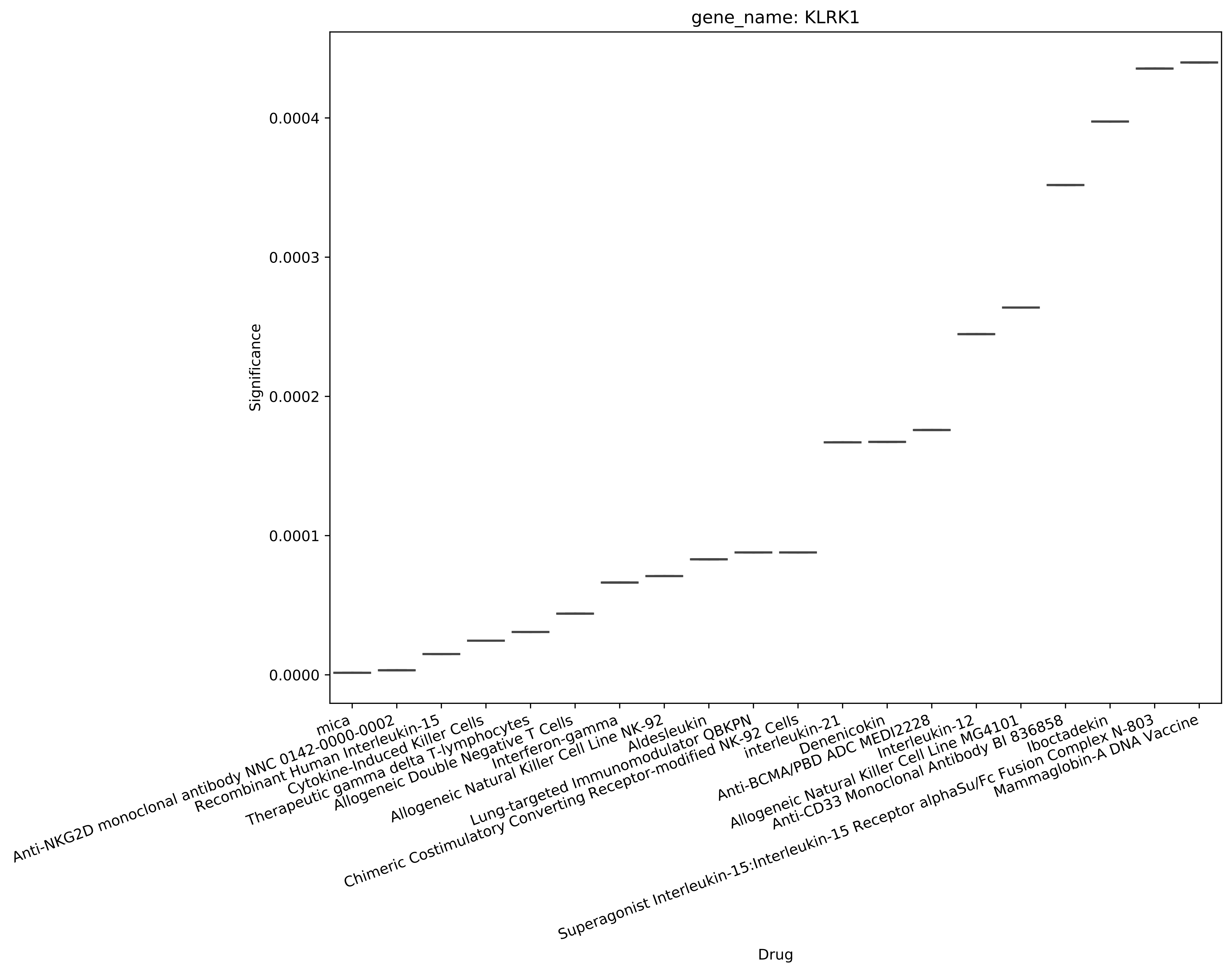
gene_name: KLRK1
Significance
0.0000
0.0001
0.0002
0.0003
0.0004
Drug
mica
Anti-NKG2D monoclonal antibody NNC 0142-0000-0002
Recombinant Human Interleukin-15
Cytokine-Induced Killer Cells
Therapeutic gamma delta T-lymphocytes
Allogeneic Double Negative T Cells
Interferon-gamma
Allogeneic Natural Killer Cell Line NK-92
Aldesleukin
Lung-targeted Immunomodulator QBKPN
Chimeric Costimulatory Converting Receptor-modified NK-92 Cells
interleukin-21
Denenicokin
Anti-BCMA/PBD ADC MEDI2228
Interleukin-12
Allogeneic Natural Killer Cell Line MG4101
Anti-CD33 Monoclonal Antibody BI 836858
Iboctadekin
Superagonist Interleukin-15:Interleukin-15 Receptor alphaSu/Fc Fusion Complex N-803
Mammaglobin-A DNA Vaccine

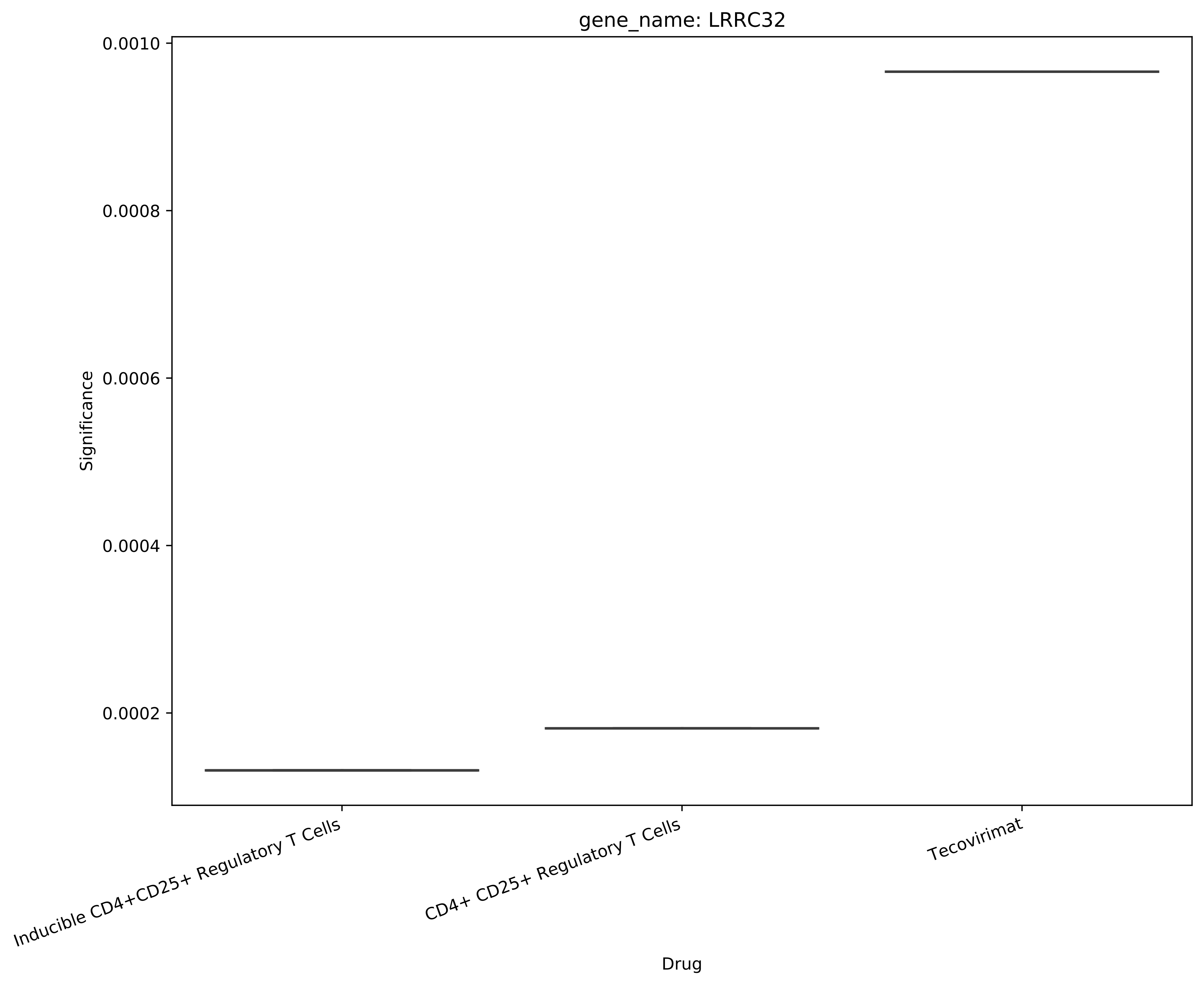

gene_name: LRRC32
Significance
0.0010
0.0008
0.0006
0.0004
0.0002
Inducible CD4+CD25+ Regulatory T Cells
CD4+ CD25+ Regulatory T Cells
Tecovirimat
Drug

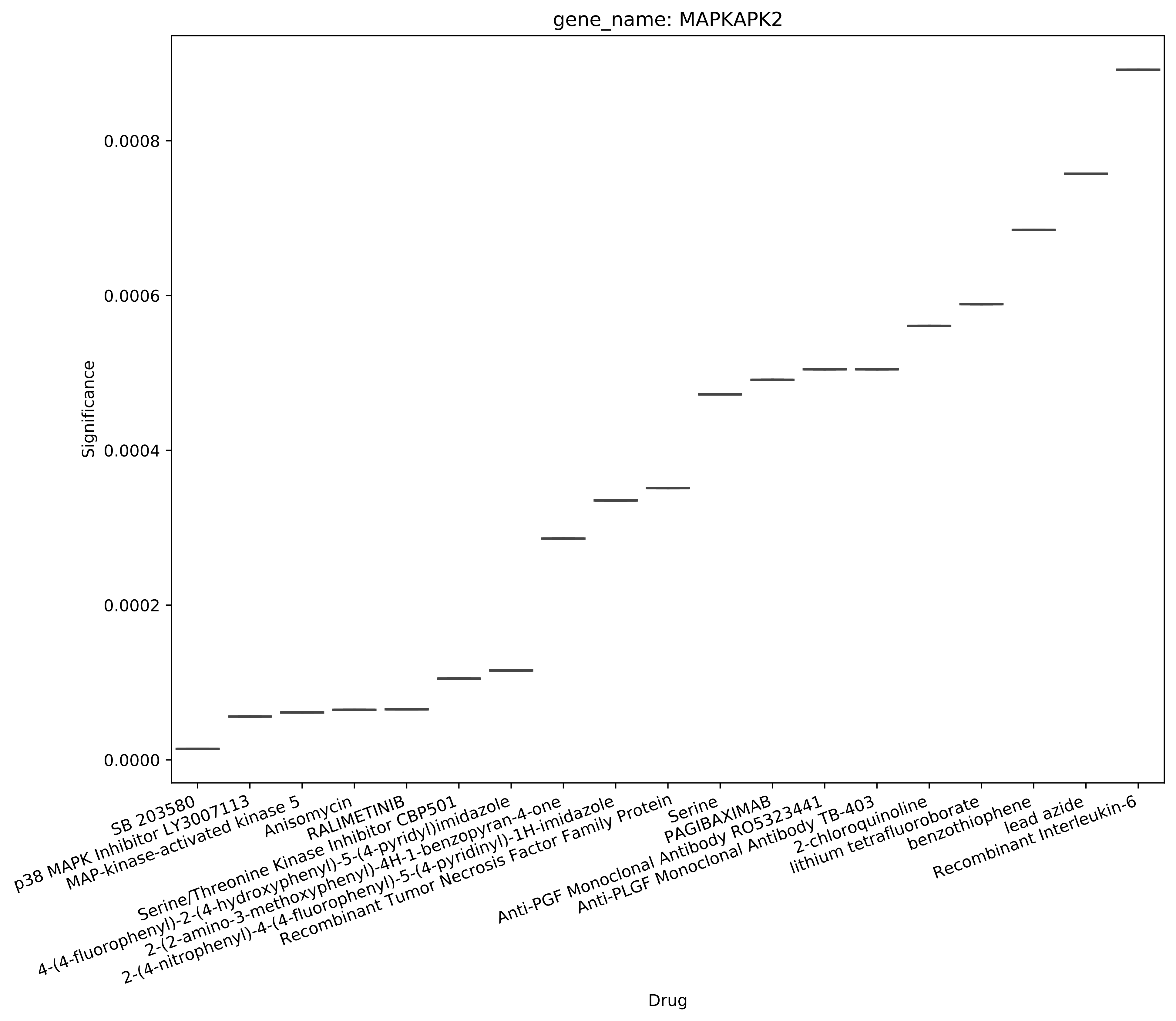
gene_name: MAPKAPK2
Significance
0.0000
0.0002
0.0004
0.0006
0.0008
Drug
SB 203580
p38 MAPK Inhibitor LY3007113
MAP-kinase-activated kinase 5
Anisomycin
RALIMETINIB
Serine/Threonine Kinase Inhibitor CBP501
4-(4-fluorophenyl)-2-(4-hydroxyphenyl)-5-(4-pyridyl)imidazole
2-(2-amino-3-methoxyphenyl)-4H-1-benzopyran-4-one
2-(4-nitrophenyl)-4-(4-fluorophenyl)-5-(4-pyridinyl)-1H-imidazole
Recombinant Tumor Necrosis Factor Family Protein
Serine
PAGIBAXIMAB
Anti-PGF Monoclonal Antibody RO5323441
Anti-PLGF Monoclonal Antibody TB-403
2-chloroquinoline
lithium tetrafluoroborate
benzothiophene
lead azide
Recombinant Interleukin-6

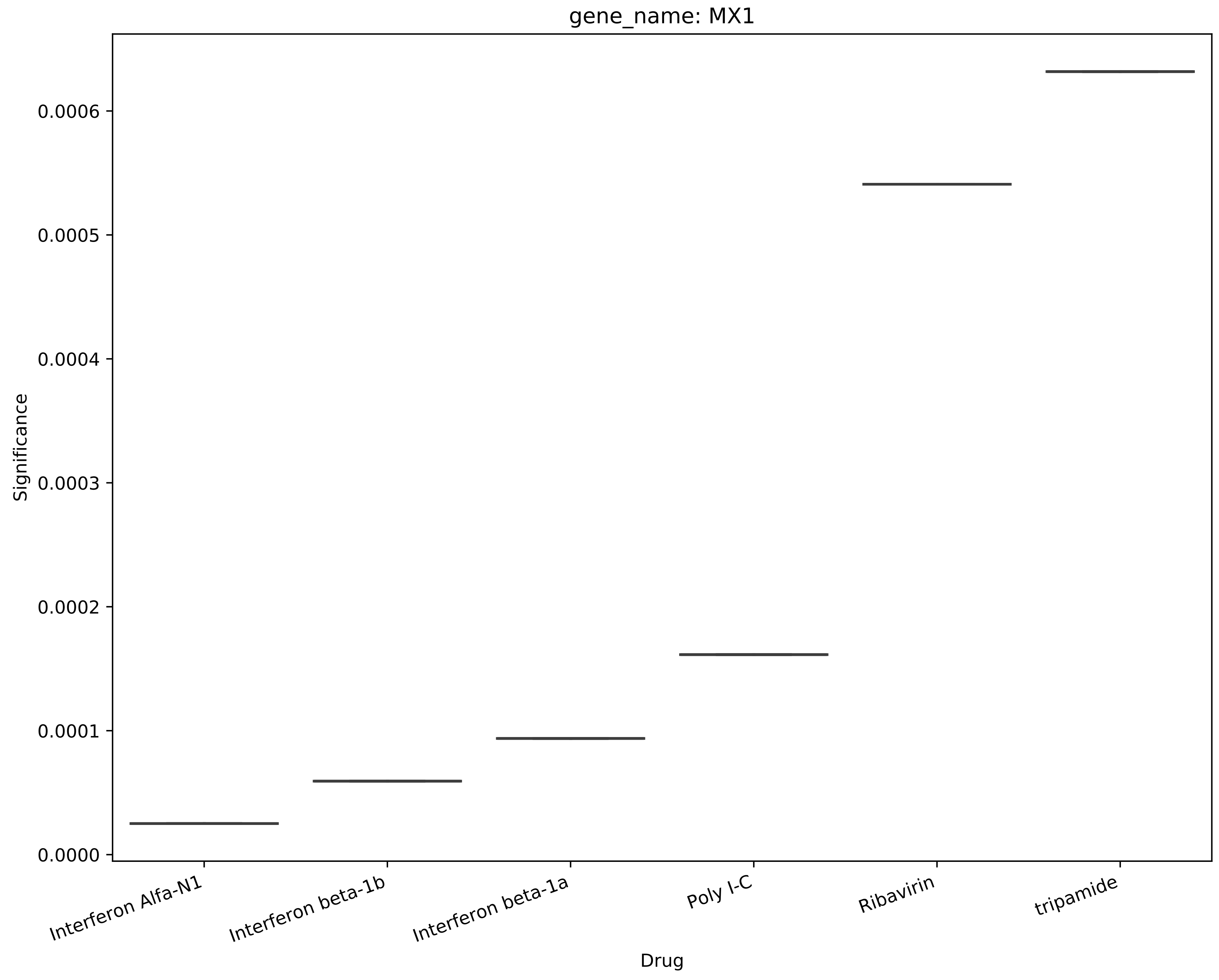
gene_name: MX1
Significance
0.0000
0.0001
0.0002
0.0003
0.0004
0.0005
0.0006
Interferon Alfa-N1
Interferon beta-1b
Interferon beta-1a
Poly I-C
Ribavirin
tripamide
Drug

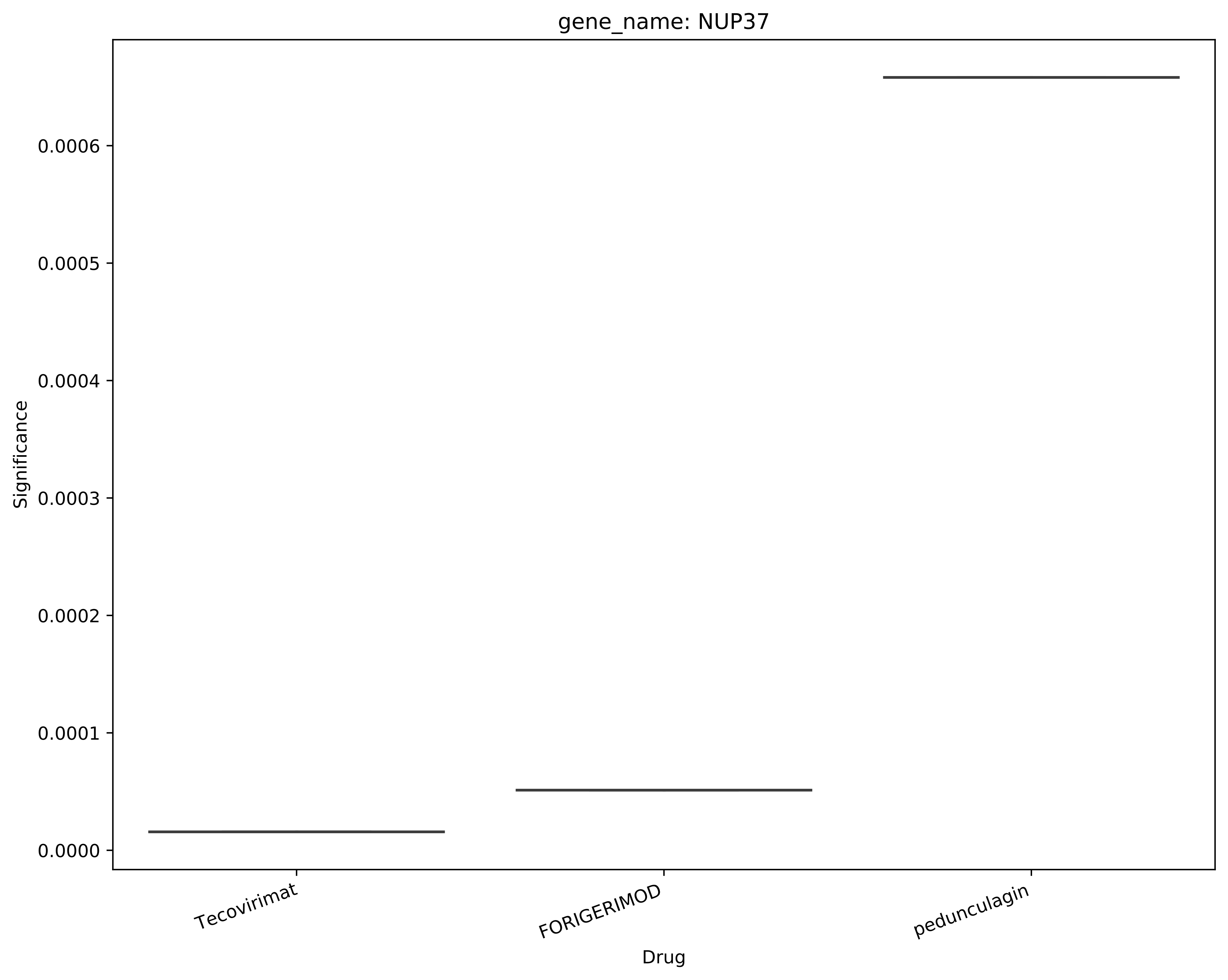
gene_name: NUP37
Significance
0.0006
0.0005
0.0004
0.0003
0.0002
0.0001
0.0000
Tecovirimat
FORIGERIMOD
pedunculagin
Drug

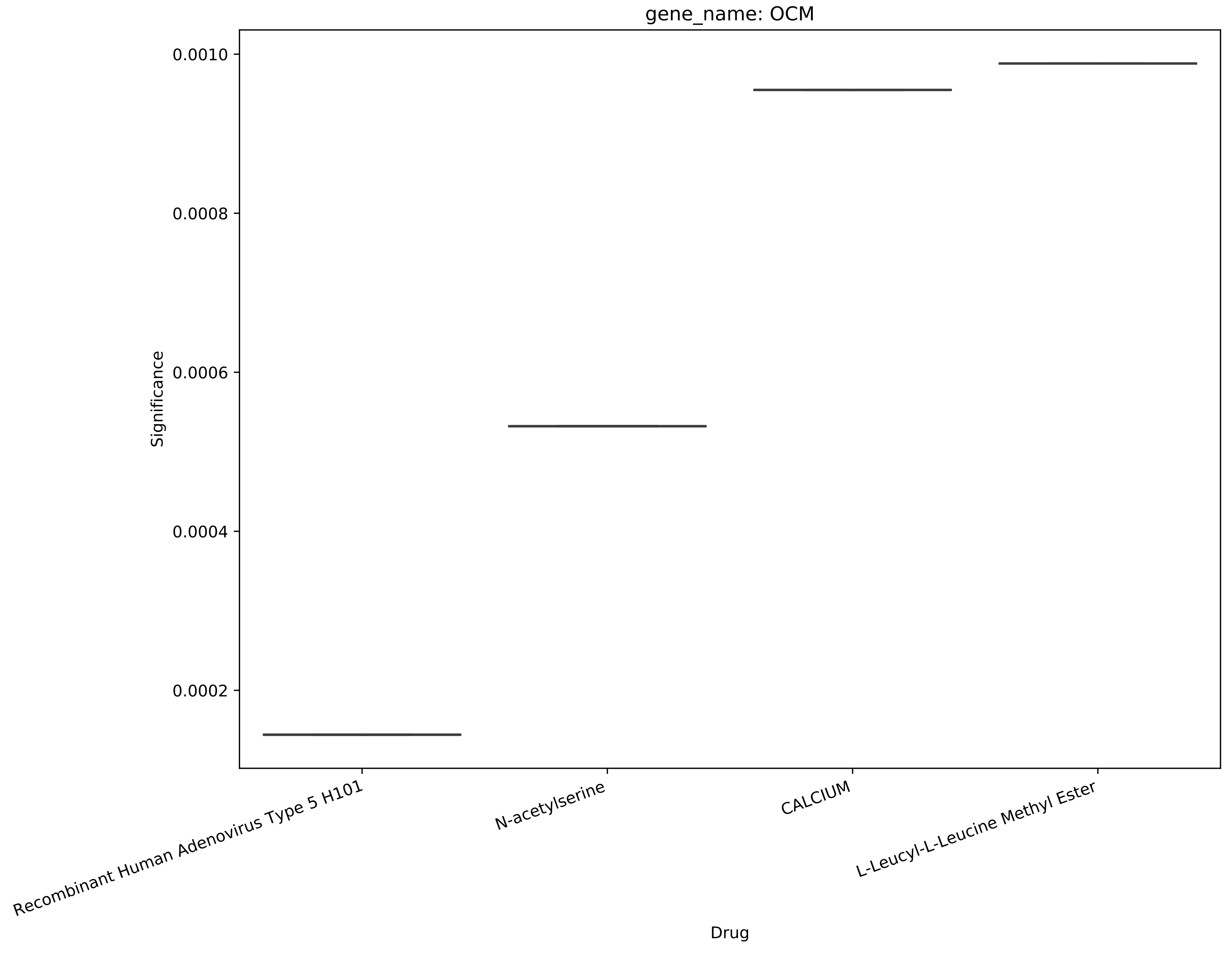
gene_name: OCM
Significance
0.0010
0.0008
0.0006
0.0004
0.0002
Recombinant Human Adenovirus Type 5 H101
N-acetylserine
CALCIUM
L-Leucyl-L-Leucine Methyl Ester
Drug

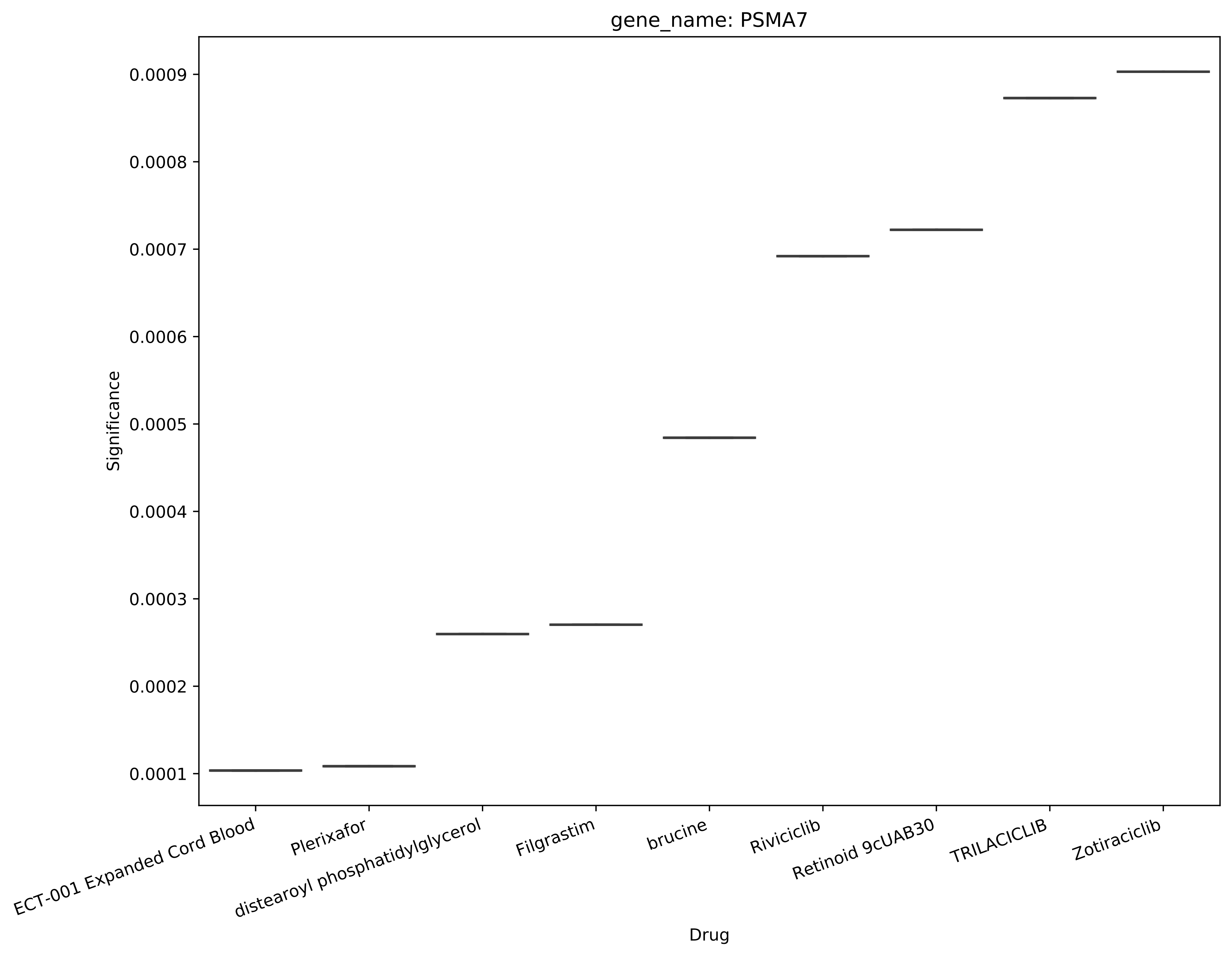
gene_name: PSMA7
Significance
0.0001
0.0002
0.0003
0.0004
0.0005
0.0006
0.0007
0.0008
0.0009
ECT-001 Expanded Cord Blood
Plerixafor
distearoyl phosphatidylglycerol
Filgrastim
brucine
Riviciclib
Retinoid 9cUAB30
TRILACICLIB
Zotiraciclib
Drug

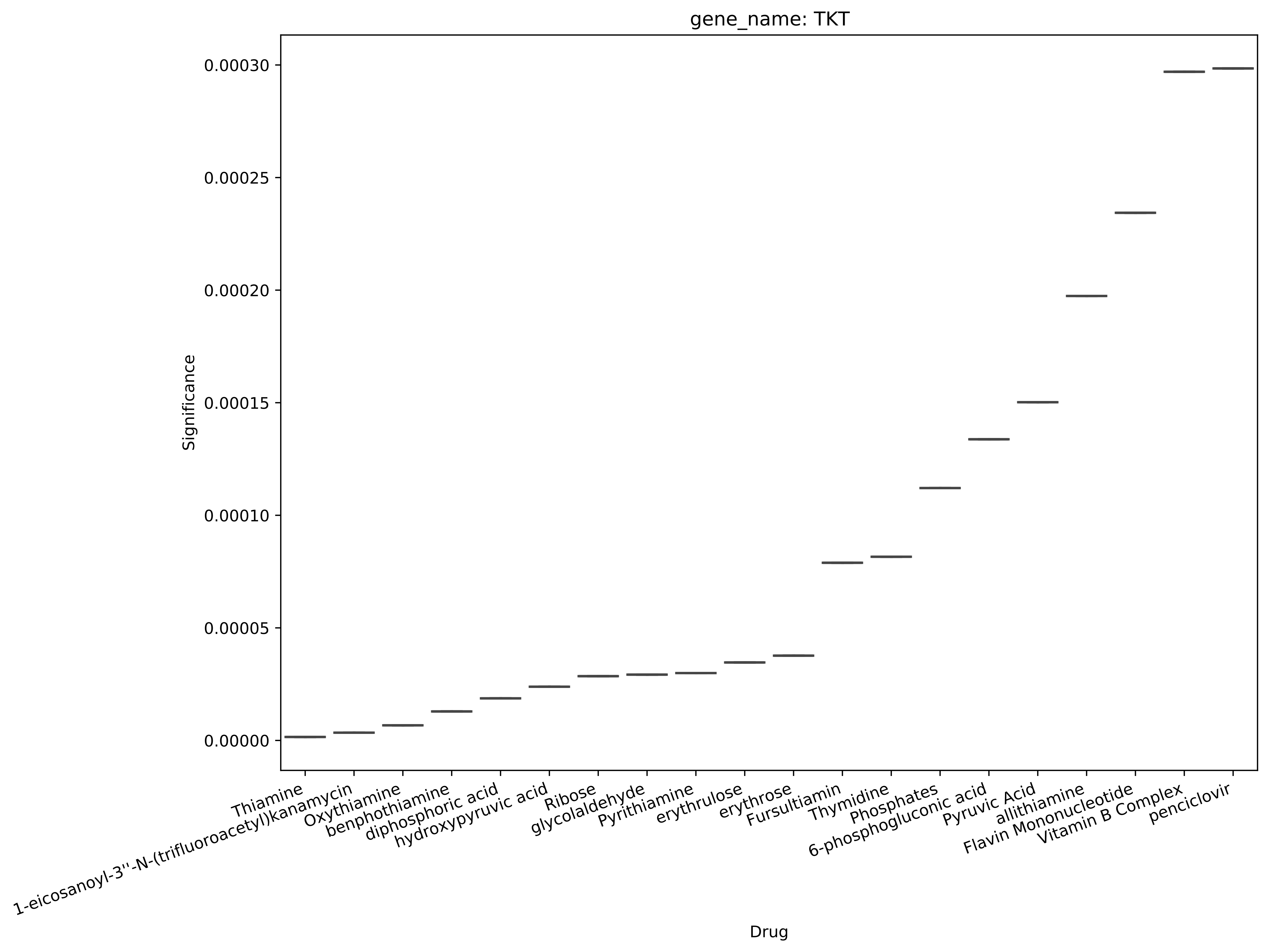

gene_name: TKT
Significance
0.00030
0.00025
0.00020
0.00015
0.00010
0.00005
0.00000
Thiamine
1-eicosanoyl-3''-N-(trifluoroacetyl)kanamycin
Oxythiamine
benphothiamine
diphosphoric acid
hydroxypyruvic acid
Ribose
glycolaldehyde
Pyrithiamine
erythrulose
erythrose
Fursultiamin
Thymidine
Phosphates
6-phosphogluconic acid
Pyruvic Acid
allithiamine
Flavin Mononucleotide
Vitamin B Complex
penciclovir
Drug

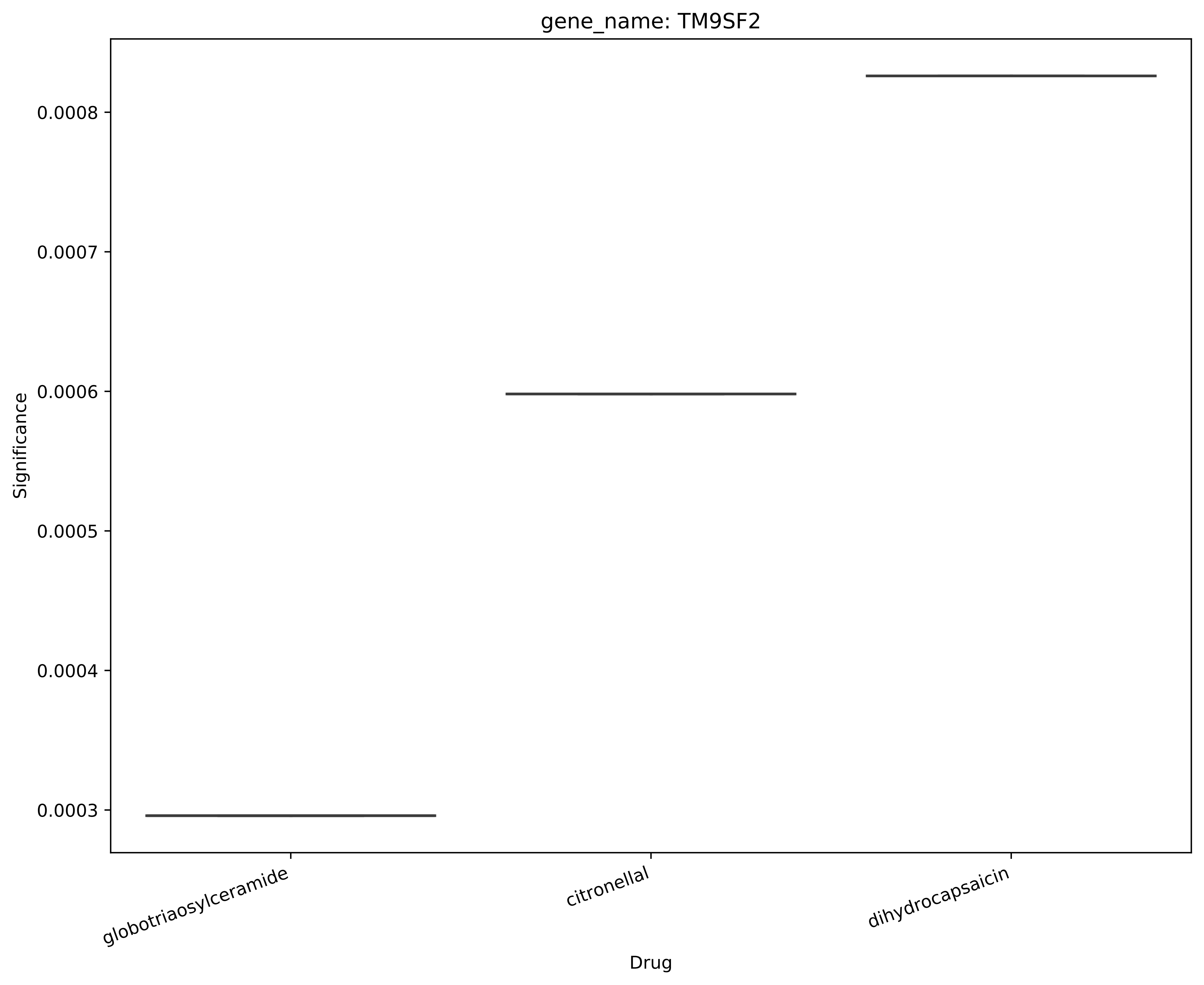

gene_name: TM9SF2
Significance
0.0008
0.0007
0.0006
0.0005
0.0004
0.0003
globotriaosylceramide
citronellal
dihydrocapsaicin
Drug

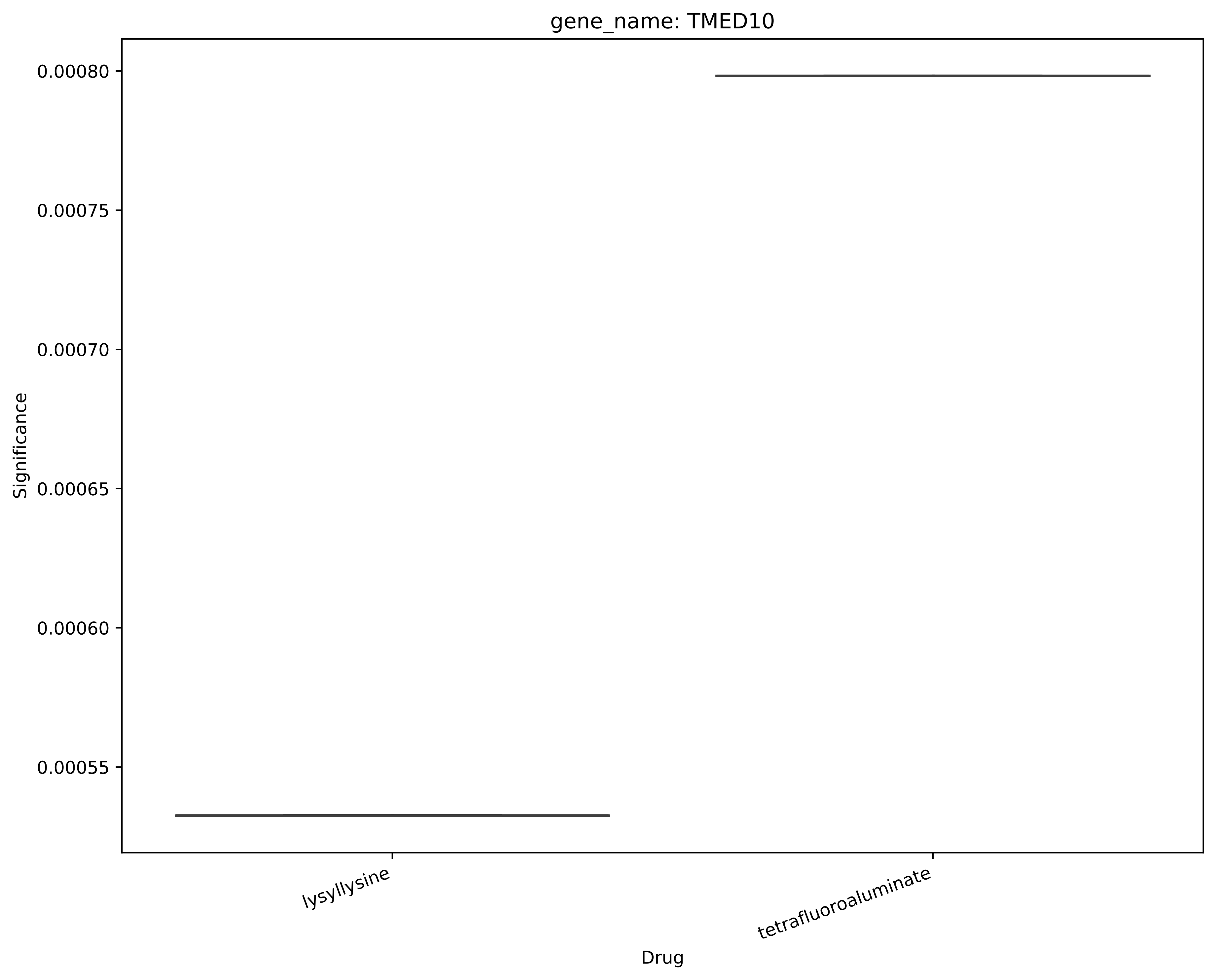

gene_name: TMED10
0.00080
0.00075
0.00070
0.00065
0.00060
0.00055
Significance
lysyllysine
tetrafluoroaluminate
Drug

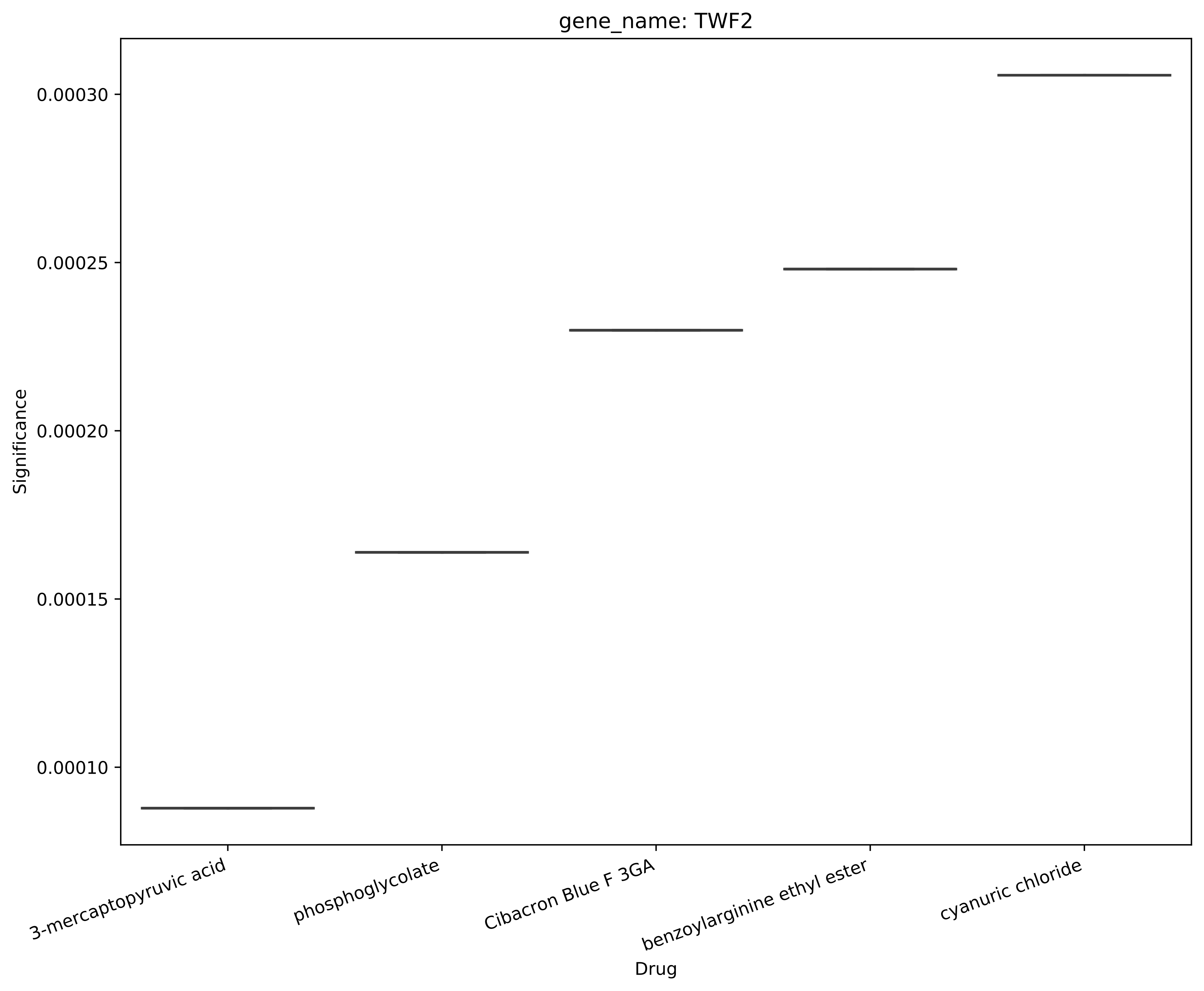
gene_name: TWF2
Significance
0.00030
0.00025
0.00020
0.00015
0.00010
3-mercaptopyruvic acid
phosphoglycolate
Cibacron Blue F 3GA
benzoylarginine ethyl ester
cyanuric chloride
Drug

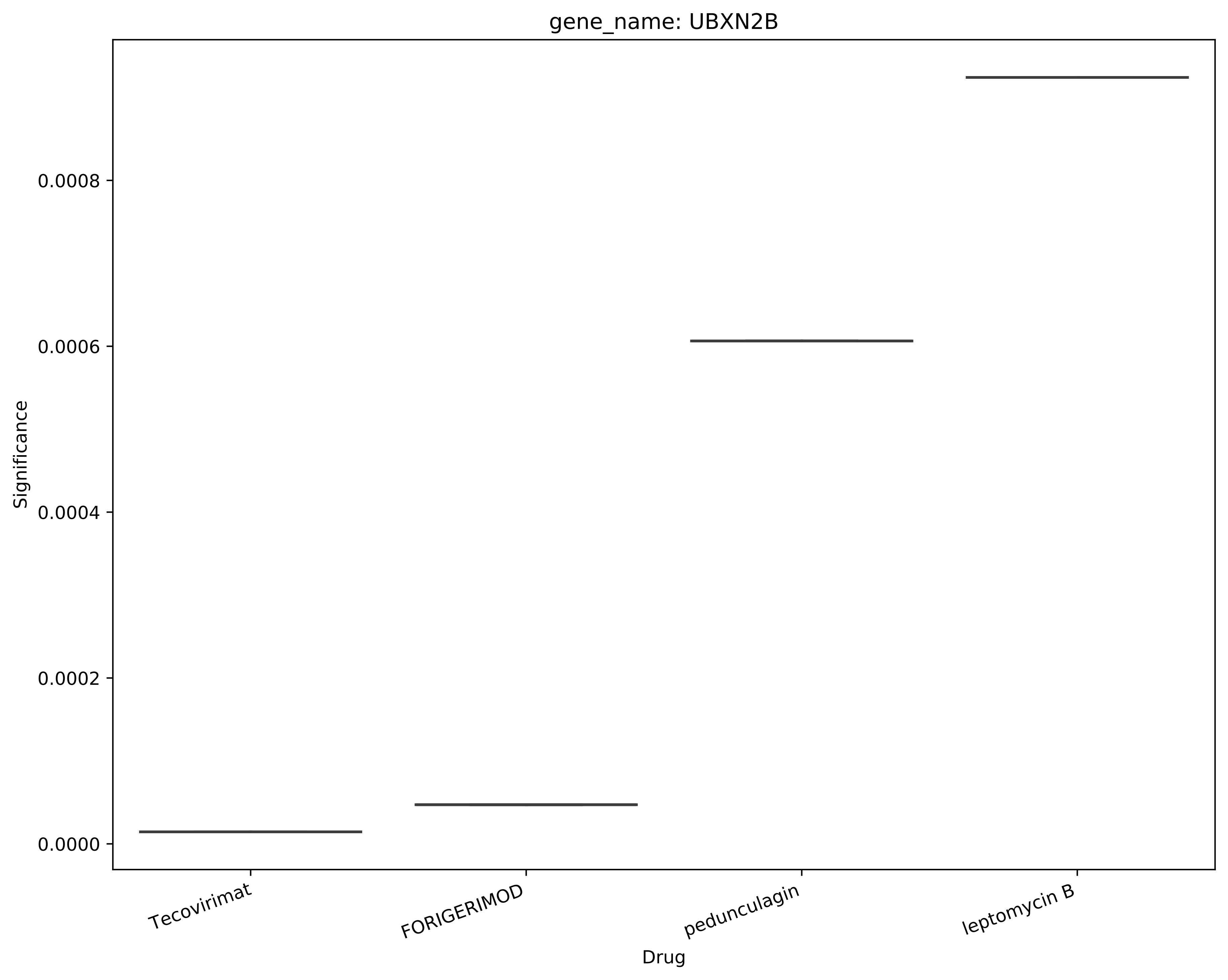
gene_name: UBXN2B
Significance
0.0000
0.0002
0.0004
0.0006
0.0008
Tecovirimat
FORIGERIMOD
pedunculagin
leptomycin B
Drug

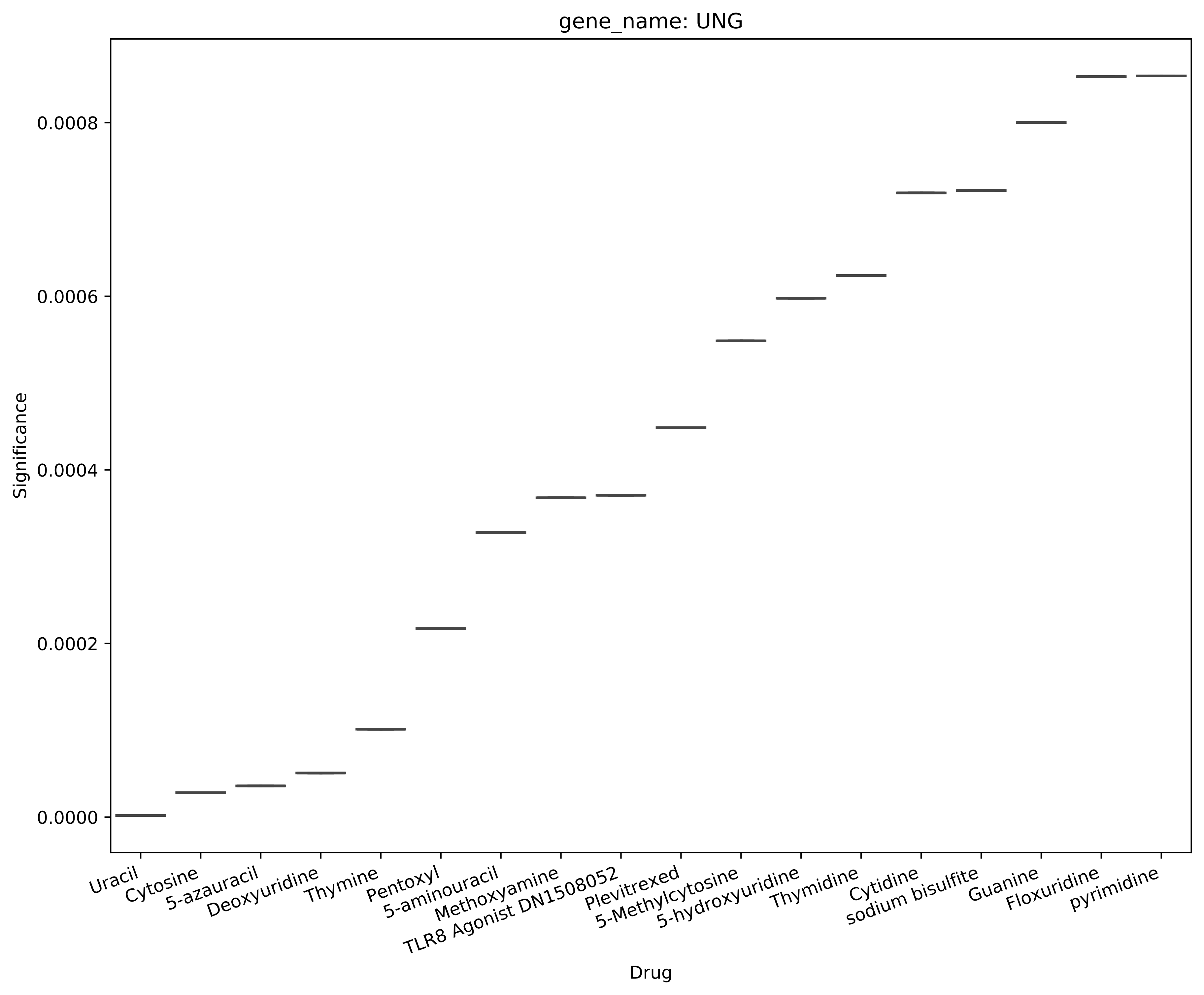
gene_name: UNG
Significance
0.0000
0.0002
0.0004
0.0006
0.0008
Uracil
Cytosine
5-azauracil
Deoxyuridine
Thymine
Pentoxyl
5-aminouracil
Methoxyamine
TLR8 Agonist DN1508052
Plevitrexed
5-Methylcytosine
5-hydroxyuridine
Thymidine
Cytidine
sodium bisulfite
Guanine
Floxuridine
pyrimidine
Drug

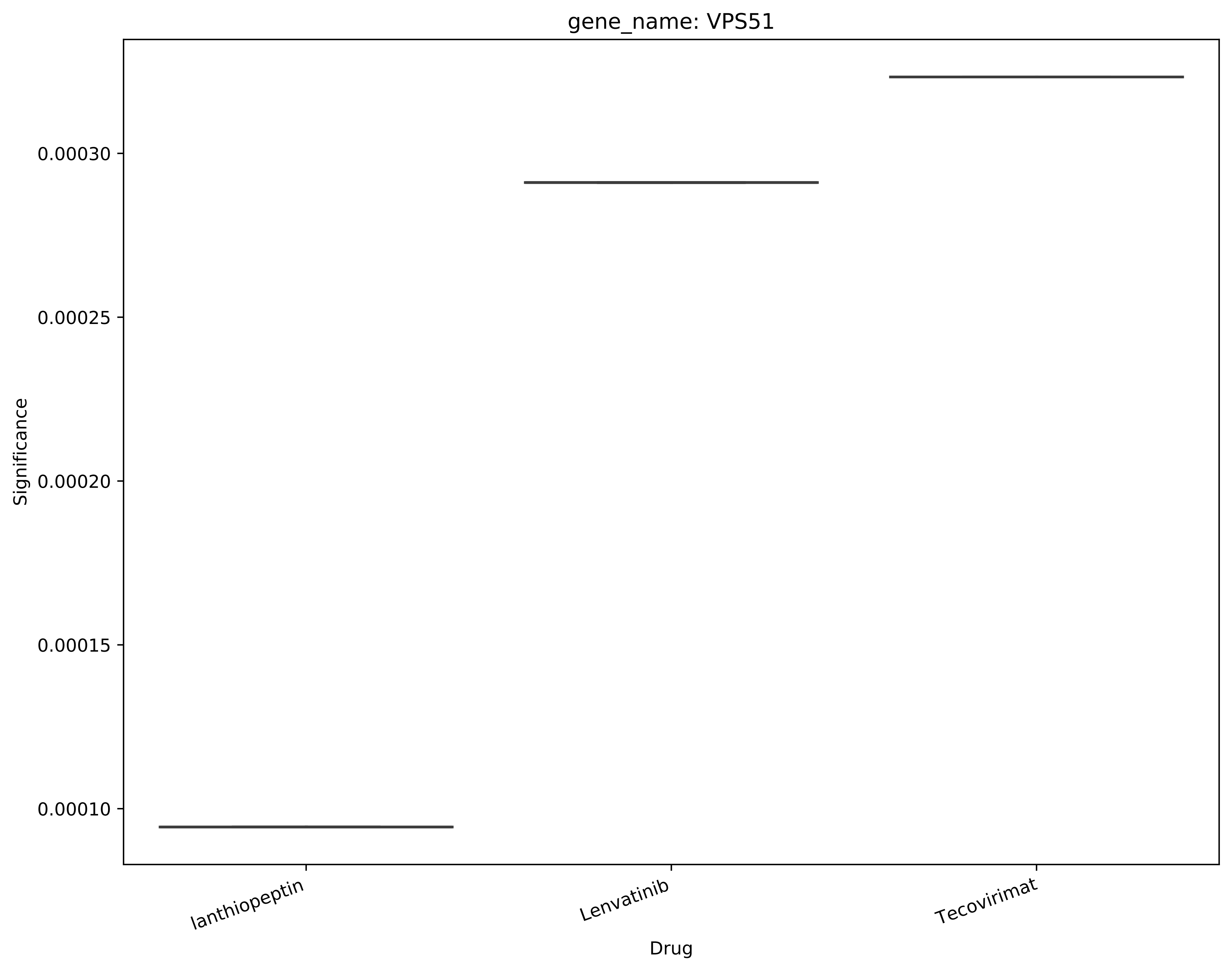

gene_name: VPS51
Significance
0.00030
0.00025
0.00020
0.00015
0.00010
lanthiopeptin
Lenvatinib
Tecovirimat
Drug

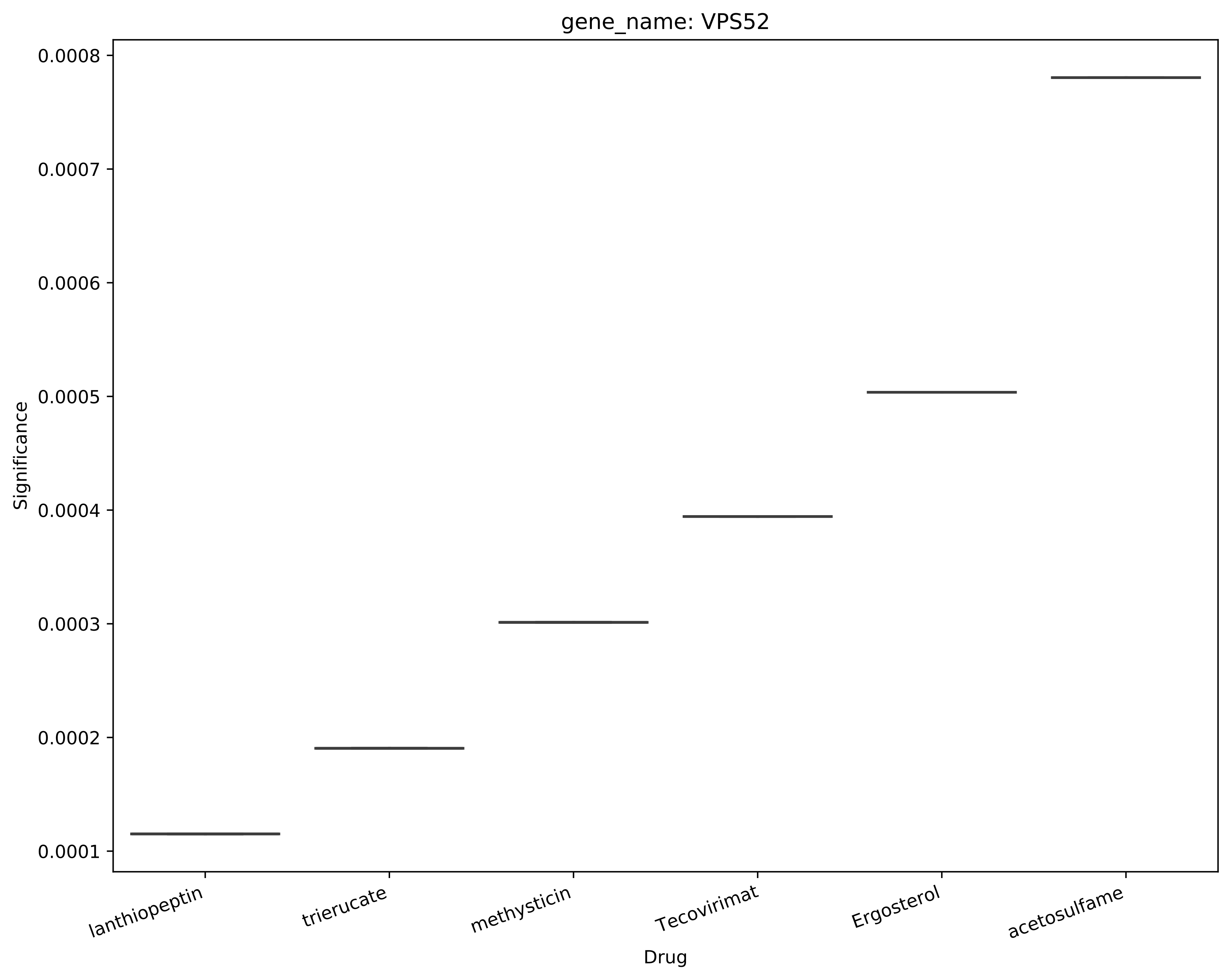
gene_name: VPS52
Significance
0.0008
0.0007
0.0006
0.0005
0.0004
0.0003
0.0002
0.0001
lanthiopeptin
trierucate
methysticin
Tecovirimat
Ergosterol
acetosulfame
Drug

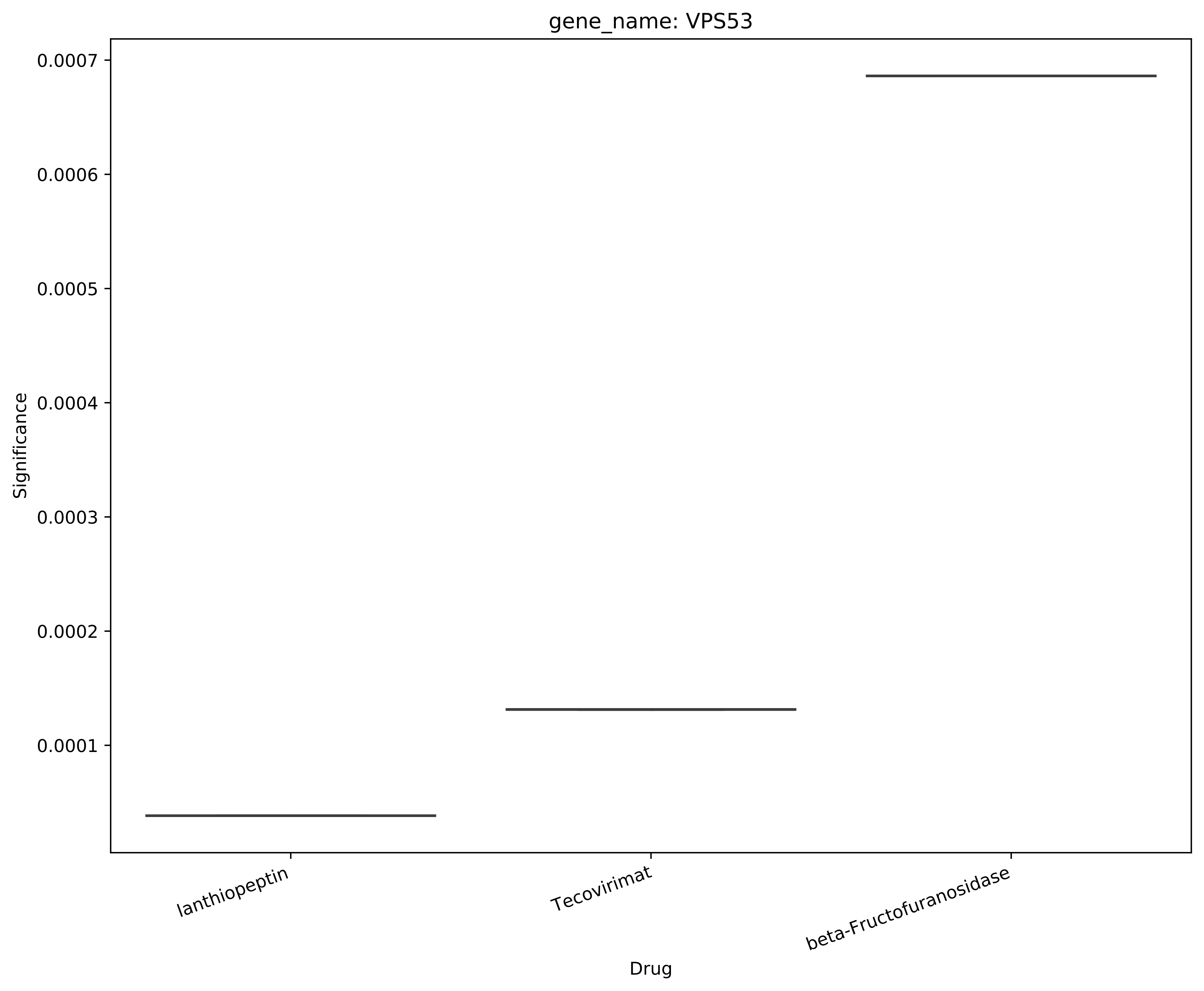
gene_name: VPS53
Significance
0.0007
0.0006
0.0005
0.0004
0.0003
0.0002
0.0001
lanthiopeptin
Tecovirimat
beta-Fructofuranosidase
Drug

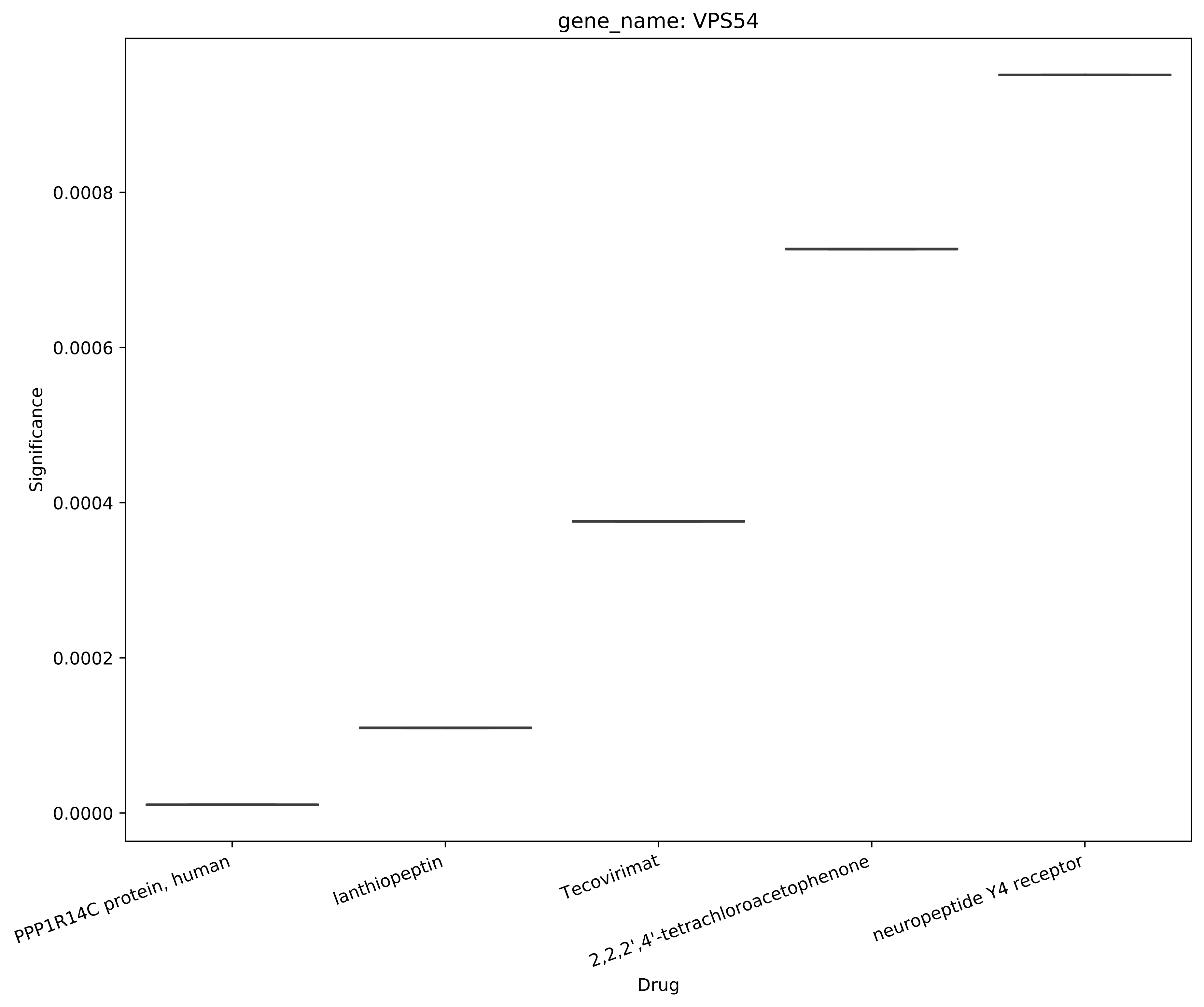
gene_name: VPS54
Significance
0.0008
0.0006
0.0004
0.0002
0.0000
PPP1R14C protein, human
lanthiopeptin
Tecovirimat
2,2,2',4'-tetrachloroacetophenone
neuropeptide Y4 receptor
Drug
