## Supplementary figures and images for "Drug combinations proposed by machine learning on genes/proteins to improve the efficacy of Tecovirimat in the treatment of Monkeypox: A Systematic Review and Network Meta-analysis"

### Assoc.v32_supplementary_material_1.pdf

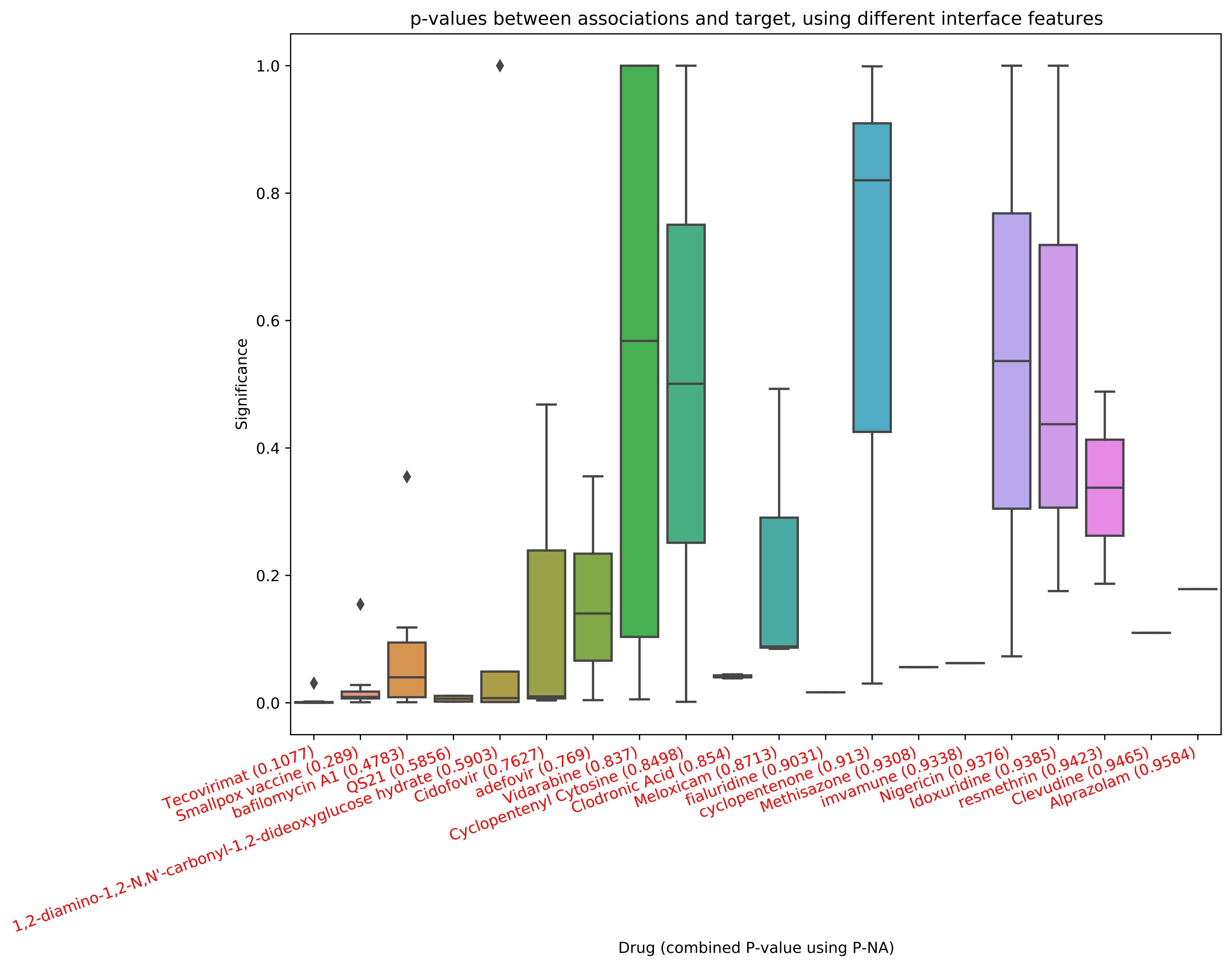

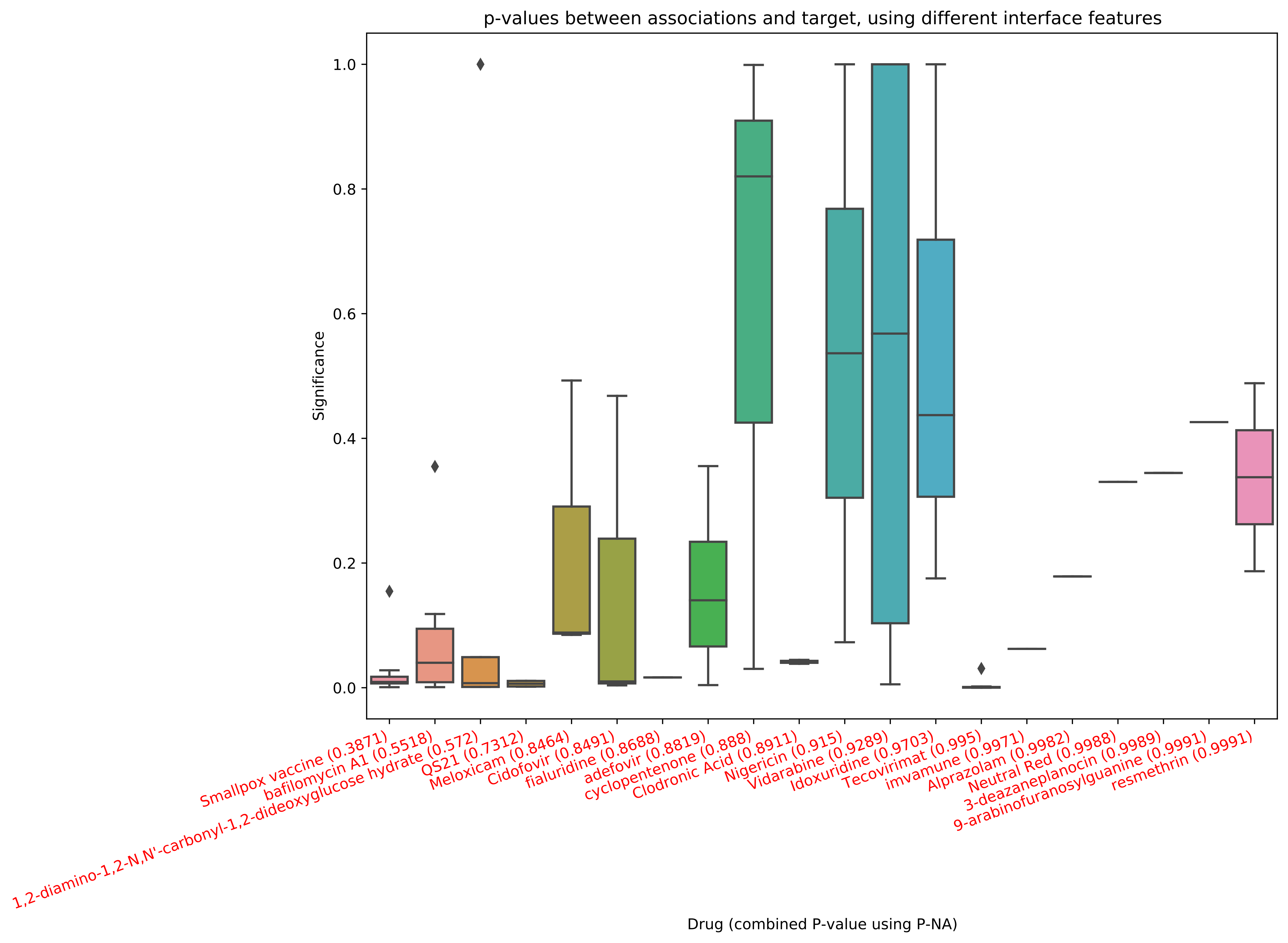

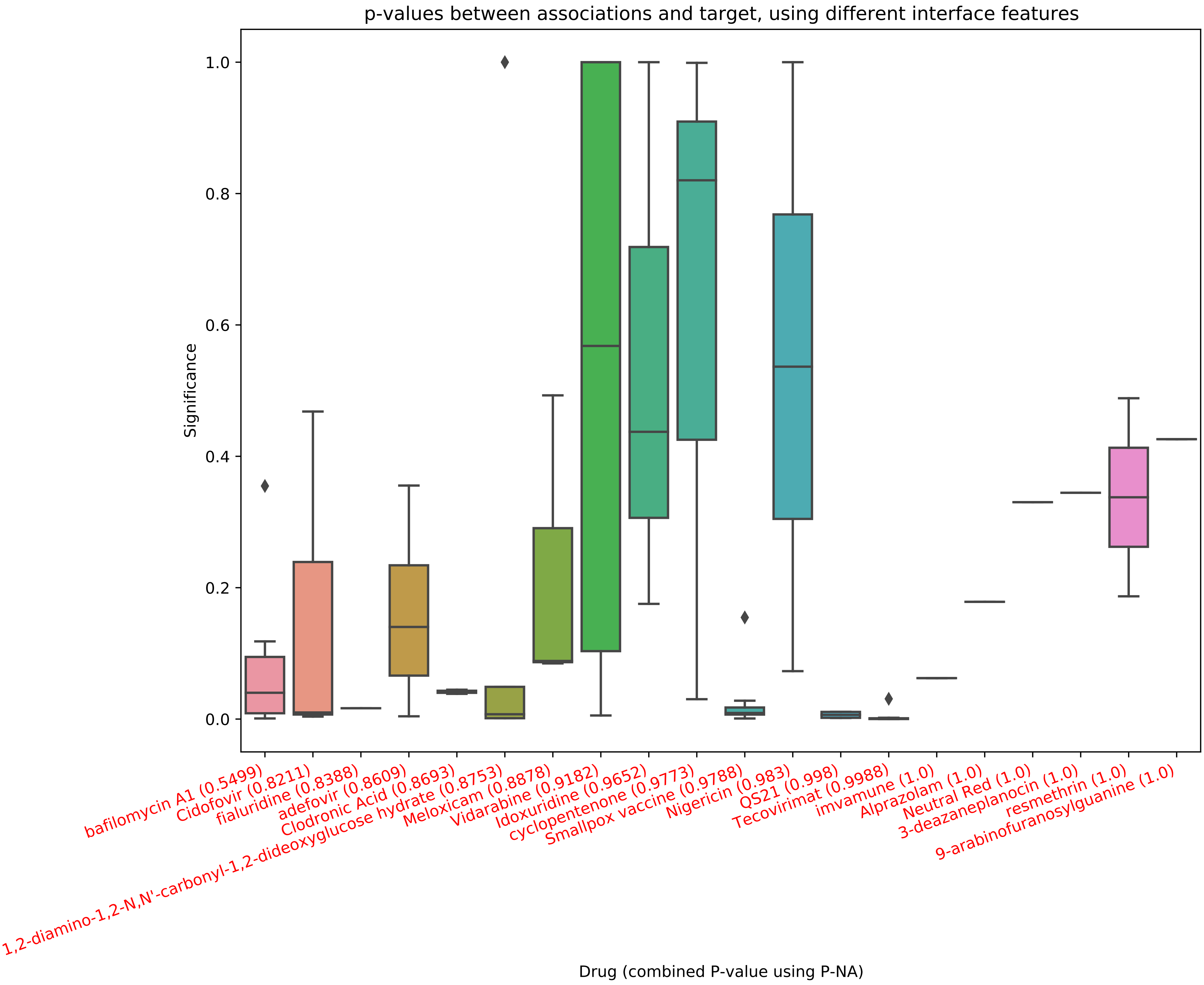

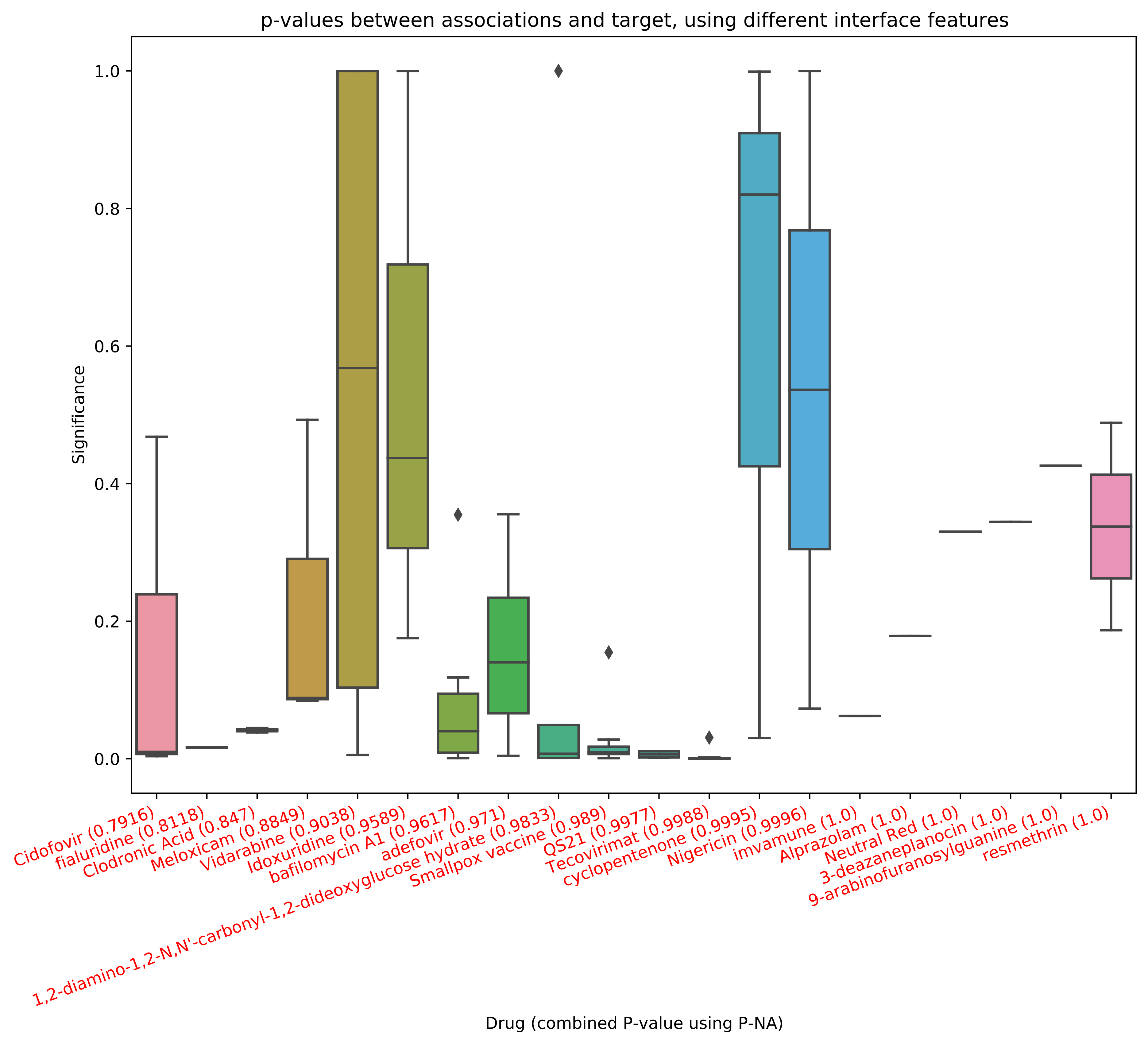

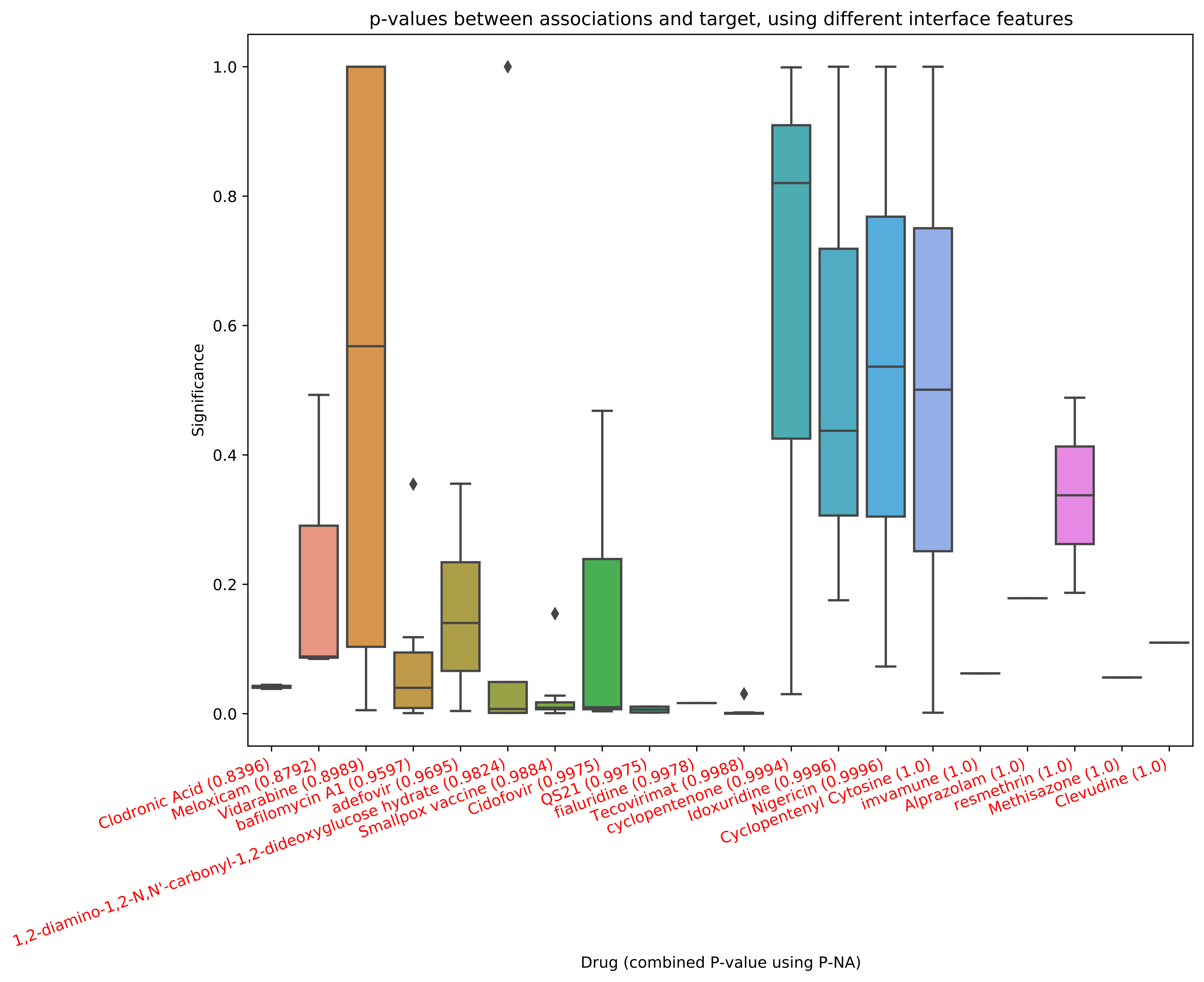
